# Dissecting causal relationships between cortical morphology and neuropsychiatric disorders: a bidirectional Mendelian randomization study

**DOI:** 10.1101/2024.09.05.24313146

**Authors:** Bochao Danae Lin, Yunzhi Li, Anastasia A. Goula, Xiao Chang, Katrina L. Grasby, Sarah Medland, Ole A. Andreassen, Bart P. F. Rutten, Sinan Guloksuz, Dennis van der Meer, Jurjen J. Luykx

**Affiliations:** Department of Psychiatry and Neuropsychology, School for Mental Health and Neuroscience, Maastricht University Medical Centre, Maastricht, The Netherlands; Department of Preventive Medicine, Institute of Biomedical Informatics, Bioinformatics Center, School of Basic Medical Sciences, Henan University, Kaifeng, China; Department of Psychiatry, UMC Utrecht Brain Center, University Medical Center Utrecht, Utrecht University, Utrecht, the Netherlands; Department of Radiology, Gansu Provincial Maternity and Child-care Hospital (Gansu Province Central Hospital), Lanzhou, Gansu, China; Institute of Science and Technology for Brain-Inspired Intelligence, Fudan University, Shanghai, 200433, China; Psychiatric Genetics, QIMR Berghofer Medical Research Institute, Brisbane, QLD, Australia; Norwegian Centre for Mental Disorders Research, KG Jebsen Centre for Psychosis Research, Division of Mental Health and Addiction, Oslo University Hospital, and Institute of Clinical Medicine, University of Oslo, Oslo, Norway; Department of Psychiatry, Yale University School of Medicine, New Haven, CT; Department of Psychiatry, Amsterdam University Medical Center, Amsterdam, the Netherlands; GGZ inGeest Mental Health Care, Amsterdam, The Netherlands

**Author notes:** Correspondence to: Jurjen J. Luykx Full address: Department of Psychiatry, Amsterdam University Medical Center, Amsterdam, the Netherlands. these authors contributed equally to this work (Bochao Danae Lin, Yunzhi Li, Anastasia A. Goula). these authors contributed equally to this work (Dennis van der Meer, Jurjen J. Luykx).

**Keywords:** Mendelian Randomization, schizophrenia, cognitive performance, cerebral cortex, cortical thinning, surface area

## Abstract

Brain cortical morphology, indexed by its surface area and thickness, is known to be highly heritable. Previous research has suggested a relationship of cortical morphology with several neuropsychiatric phenotypes. However, the multitude of potential confounders makes it difficult to establish causal relationships. Here, we employ Generalized Summary-data-based Mendelian Randomization and a series of sensitivity analyses to investigate causal links between 70 cortical morphology measures and 199 neuropsychiatric, behavioral, and metabolic phenotypes. We show that total brain cortical surface area (TSA) has significant positive causal effects on 18 phenotypes. The strongest effects include TSA positively influencing cognitive performance, while reverse analyses reveal small effects of cognitive performance on TSA. Global mean cortical thickness (MTH) exhibits significant causal effects on five phenotypes, including schizophrenia. MTH reduces schizophrenia risk and bidirectional causality is found between MTH and smoking initiation. Finally, in regional analyses we detect positive influences of the transverse temporal surface area on cognitive performance and negative influences of transverse temporal thickness on schizophrenia risk. Overall, our results highlight bidirectional associations between TSA, MTH, and neuropsychiatric traits. These insights offer potential avenues for intervention studies aimed at improving brain health.

## Introduction

Cortical thickness and surface area are highly heritable^1,2^ and are strongly associated with neuropsychiatric phenotypes, including substance use disorders.^3–7^ To improve our understanding of neurobiological determinants and neuropsychiatric disorders, it is critical to dissect the interplay between cortical morphology and neuropsychiatric phenotypes.

Establishing causality between brain measures and neuropsychiatric phenotypes is challenging due to numerous possible confounding factors, e.g., medication use^8^ and comorbid conditions.^9^ Mendelian Randomization (MR) may overcome several challenges pertinent to other lines of research, such as reverse causation, confounding and other types of bias.^10^ Of particular relevance for neuropsychiatric disorders is the possible impact of (psychotropic) medication on brain morphology, ^11,12^ which is circumvented in MR.

Recent MR studies have examined causal links between brain measures and psychiatric disorders, including substance use.^13,14^ One study identified brain measures with causal influences on the risks of schizophrenia, anorexia nervosa, and bipolar disorder in the UK Biobank (UKB).^11^ Another MR study detected negative associations between global cortical thickness and alcohol drinking behaviour.^12^ Considering the importance of cortical morphology for cognitive functioning and the previously detected associations of cortical morphology with neuropsychiatric traits,^15,16^ we set out to further elucidate its role and how it is related to mental health.^16–18^

Given the co-occurrence of inflammation, metabolite changes, and type 2 diabetes mellitus (T2D) with neuropsychiatric phenotypes,^9,19,20^ it is of interest to examine cortical morphology changes in the context of metabolic phenotypes. For example, inflammation may contribute to structural brain changes through the activation of microglia and/or astrocytic dysfunction in various neuropsychiatric disorders.^21^ Moreover, alterations in cortical thickness have been observed in middle-aged patients with T2D, which may be related to diabetes-related brain damage.^22^ Additionally, cortical thinning was observed in relation to body mass index (BMI) and visceral adipose tissue (VAT), likely involving a mechanism of adipose tissue-related low-grade inflammation.^23^ Other findings suggest that C-reactive protein (CRP) and vitamin D exert causal effects on region-specific cortical thickness and point to a negative causal relationship between serum CRP levels and the thickness of the lingual region.^24^ Therefore, examining causal relationships between a range of metabolic traits and cortical morphology could provide clues about the underlying mechanisms of various (neuropsychiatric) disorders.

Here, we hypothesized that specific cortical morphology measures have causal relationships with neuropsychiatric, behavioral, and metabolic phenotypes. By leveraging the availability of recently published large genome-wide data, we aimed to shed light on causal mechanisms using a thorough and stringent methodology, namely Generalized Summary-data-based Mendelian Randomization (GSMR). By incorporating the heterogeneity in dependent instruments (HEIDI) method,^25,26^ GSMR is powerful and conservative in identifying and removing pleiotropic instrumental variables.^27^ In addition, we used cortical morphology data from Enhancing Neuro-Imaging Genetics through Meta-Analysis (ENIGMA),^28^ which has several advantages over the use of genome-wide association studies (GWASs) based only on UK Biobank participants. For example, ENIGMA is a brain imaging-focused study that collected data from many sites across the globe, thus increasing the chances of generalizability.^29^ Finally, by including an unprecedented set of neuropsychiatric, behavioral, and metabolic phenotypes, we aimed to refine the understanding of directions of effect between cortical morphology and such phenotypes. We thus searched for any bidirectional causal relationships between 70 cortical morphology measures (both global and regional measures)^29^ and 199 phenotypes spanning neuropsychiatric, behavioral and metabolic phenotypes. We highlight several new unidirectional and bidirectional relationships that extend our understanding of the relationships between neuropsychiatric disorders and cortical morphology. The findings provide actionable insights that could inform the development of targeted lifestyle interventions for neuropsychiatric disorders.

## Results

### Overview: analysis set-up & genetic instruments

We performed forward GSMR analyses of total surface area (TSA) and mean cortical thickness (MTH, *N*=∼0.5 million subjects) on the 199 phenotypes explained in the methods section (*N*=∼13.5 million subjects; Figure 1; also see STable 1-2, Supplementary tables ST1-ST5. Of those phenotypes, 172 fulfilled the criterion of two or more instrument variables (IVs), allowing us to also perform reverse GSMR analyses on cortical morphology measures (STable 2, Supplementary tables ST3, ST5), along with the respective sensitivity analyses for these 172 phenotypes (Supplementary tables ST6, ST7). As the instrument strength was strong for all phenotypes tested (F-statistics in forward and reverse MR analyses ranging from 28.11 to 98.89), we did not find any evidence of weak instrument bias. For all 199 GSMR tests of MTH effects, the number of genetic instruments was ≥10, except for 9 tests for which the numbers of instruments ranged from 7-9 (Supplementary tables ST4). Out of these four tests, two were significant: the protective effects of MTH on SCZ (using 9 instruments) and on number drinks per week (using 8 instruments). Finally, in the regional analyses, we observed that the global-corrected method yields less potential causal relationships than its non-global-corrected counterpart.

**Figure 1.**
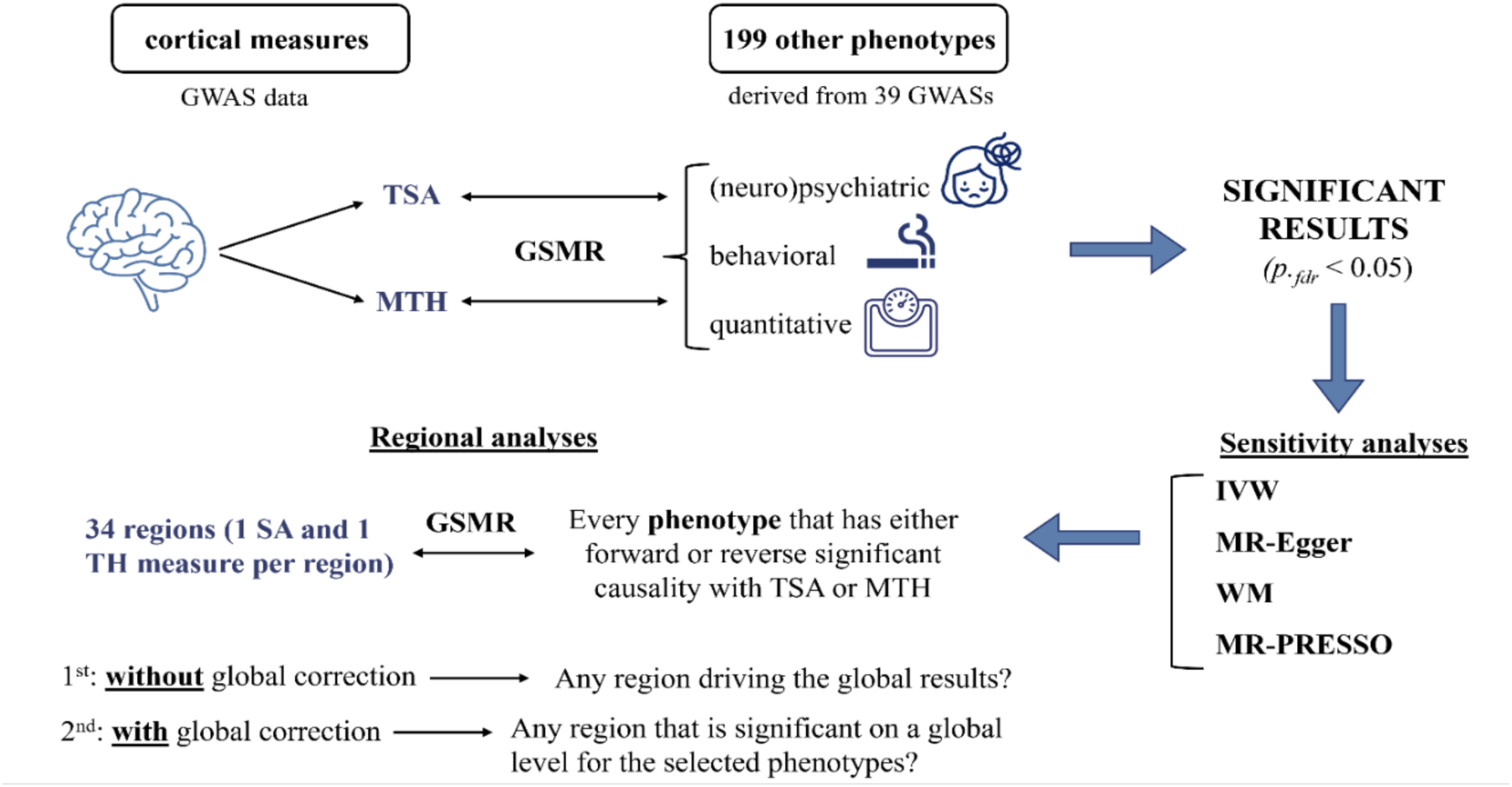
Schematic overview of our study design. TSA: total surface area, MTH: mean cortical thickness, GSMR: Generalized Summary-data-based Mendelian Randomization, IVW: Inverse Variance Weighted fixed-effects method, MR-Egger method, WM: Weighted Median MR method, MR-PRESSO: MR-pleiotropy residual sum and outlier method.

### Relationships between surface area and other phenotypes

#### Forward analyses

We identified significant causal effects of TSA on 18 phenotypes (Figure 2A, Supplementary Figure 1A). Further details on beta values, standard errors and p-values can be found in the Supplementary tables ST2, where the significant results are marked in bold. TSA had significant causal effects with a positive effect direction on eight outcomes, namely cognitive performance (CP),^30^ educational attainment (EA),^31^ height,^32^ cross disorders group (CDG),^33^ age of smoking initiation 2022 (AgeSmk#2),^34^ high-density lipoprotein cholesterol (HDL),^35^ bipolar disorder (BIP),^36^ and age of smoking initiation 2019 (AgeSmk).^37^ Furthermore, TSA had significant negative causal effects on 10 outcomes, namely smoking initiation 2022 (SmkInit#2),^34^ log-transformed triglycerides (logTG),^35^ smoking initiation 2019 (SmkInit),^39^ attention-deficit/hyperactivity disorder (ADHD2019),^38^ smoking cessation 2022 (SmkCes#2),^34^ total cholesterol (TC),^35^ non-high-density lipoprotein cholesterol (nonHDL),^35^ Alzheimer’s disease (ALZ),^39^ type II diabetes 2017 (T2D2017), and type II diabetes 2020 (T2D2020).^40^ The three most significant effects were found for the following phenotypes: CP (*β*=0.56, *se*=0.04, *p._fdr_*=8.2 x 10^-41)^, EA (*β*=0.40, *se*=0.03, *p._fdr_*=9.1 x 10^-32^), and height (*β*=0.35, *se*=0.04, *p._fdr_*=2.7 x 10^-15^). A forest plot summarizes the above-mentioned findings using beta values in Figure 2A and odds ratios (ORs) in SFigure 1A.

**Figure 2.**
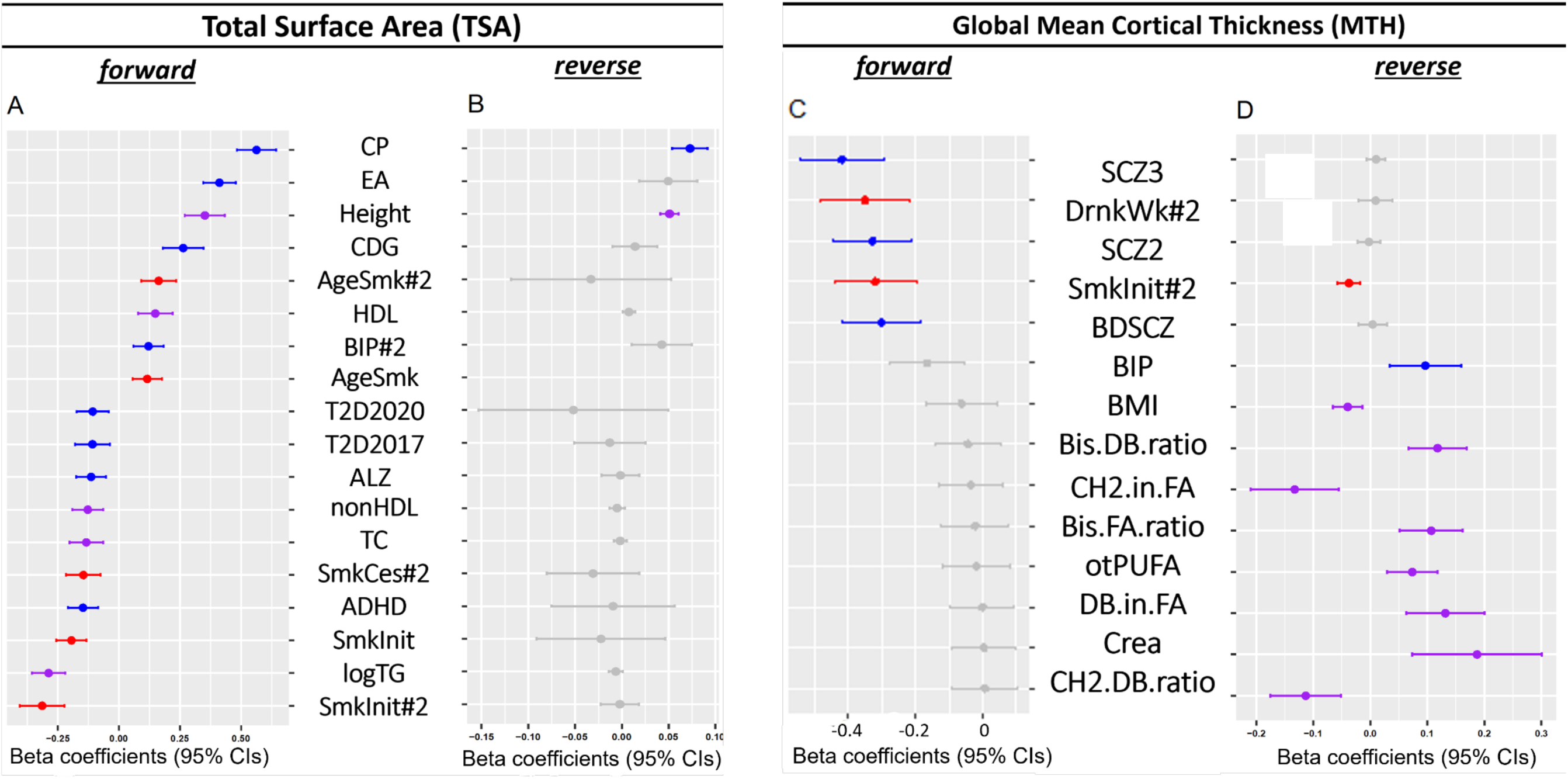
Forest plots of GSMR models with significant results (*p._fdr_* < 0.05) for ΤSA and ΜTH. (**A**) Forward MR results of ΤSA on outcomes. (**B**) Reverse MR results of outcomes on ΤSA. (**C**) Forward MR results of ΜTH on outcomes. (**D**) Reverse MR results of outcomes on ΜTH. Significant results (p.fdr < 0.05) are colored by category: red= behavior, purple=quantitative phenotype and blue=(neuro)psychiatric phenotype (except for T2D2017, TSD2020). The non-significant results (p.fdr > 0.05) are colored in grey. In Fig.2B, there were not enough significantly associated SNPs for age of smoking initiation 2019 (AgeSmk) to be extracted as instruments and hence the result is unavailable. (**A & B**) CP: cognitive performance, EA: educational attainment, Height, CDG: cross disorders group, AgeSmk#2: age of smoking initiation (2022), HDL: high-density lipoprotein cholesterol, BIP#2: bipolar disorder excluding participants from the UK biobank (2021), AgeSmk: age of smoking initiation (2019), T2D2020: type II diabetes (2020), T2D2017: type II diabetes (2017), ALZ: Alzheimer’s disease, nonHDL: non-high-density lipoprotein cholesterol, TC: total cholesterol, SmkCes#2: smoking cessation (2022), ADHD: attention deficit hyperactivity disorder, SmkInit: ever smoked regularly (2019), logTG: log-transformed triglycerides, SmkInit#2: ever smoked regularly (2022) (**C & D**) SCZ3: schizophrenia freeze 3, DrnkWk#2: drinks per week (2022), SCZ2: schizophrenia freeze 2, SmkInit#2: ever smoked regularly (2022), BDSCZ: meta-analysis bipolar disorder and schizophrenia, BIP: bipolar disorder (2019), BMI: body mass index, Bis.DB.ratio: ratio of bisLallylic bonds to double bonds in lipids, CH2.in.FA: CH2 groups in fatty acids, Bis.FA.ratio: ratio of bisLallylic bonds to total fatty acids in lipids, otPUFA: other polyunsaturated fatty acids than 18:2, DB.in.FA: double bonds in fatty acids, Crea: creatinine, CH2.DB.ratio: CH2 groups to double bonds ratio.

#### Reverse analyses

We identified two phenotypes - height and cognitive performance - with significant causal effects on TSA (Figure 2B for beta values and SFigure 1B for ORs). We thus detected positive bidirectional causal relationships between TSA-CP (forward: *β*=0.56, 95%CI=0.48-0.64, *se*=0.04, *p._fdr_*=8.2 x 10^-41^, reverse: *β*=0.07, 95%CI=0.05-0.09, *se*=0.01, *p._fdr_*=3.53 x 10^-12^) and TSA-height (forward: *β*=0.35, 95%CI=0.27-0.34, *se*=0.04, *p._fdr_*=2.7 x 10^-15^, reverse: *β*=0.05, 95%CI=0.04-0.06, *se*=0.004, *p._fdr_*=1.7 x 10^-23^). Statistical details can be found in Supplementary tables ST3.

### Relationships between cortical thickness and other phenotypes

#### Forward analyses

MTH had significant negative causal effects on five outcomes (see Figure 2C for beta values and SFigure 1C for ORs), namely schizophrenia (SCZ freeze 3 (SCZ3)^41^ and SCZ freeze 2 (SCZ2)^42^), alcoholic drinks consumed per week 2022 (DrnkWk#2),^34^ smoking initiation 2022 (SmkInit#2)^34^ and bipolar disorder and SCZ considered together as one phenotype (BDSCZ).^43^ The most significant result was the causal effect of MTH on SCZ3 (*β*=-0.42, 95%CI=-0.54 - -0.29, *se*=0.06, *p._fdr_*=1.02 x 10^-8^; statistical details in Supplementary tables ST4).

#### Reverse analyses

We identified 10 significant phenotypes with causal effects on MTH (Figure 2D for beta values, SFigure 1D for ORs). We detected positive causal effects of blood metabolites (Bis.DB.ratio, Bis.FA.ratio, DB.in.FA, otPUFA, Crea)^44^ and bipolar disorder (BIP)^36^ on MTH, along with negative causal effects of smoking initiation 2022 (SmkInit#2)^34^, CH2.DB.ratio,^44^ CH2.in.FA,^44^ and BMI^45^ on TH. The most significant result was found for the ratio of bisLallylic bonds to double bonds in lipids (Bis.DB.ratio, *β*=0.12, 95%CI=0.07-0.17, *se*=0.026, *p._fdr_*=0.001). The full names of all abbreviated metabolic traits are in STable 3. Based on the forward and reverse analyses, significant negative bidirectional causal effects between MTH and smoking initiation were established (forward: *β*=-0.32, 95%CI=-0.44- -0.22, *se*=0.06, *p._fdr_*= 1.5 x 10^-5^, reverse: *β*=- 0.04, 95%CI=-0.06- -0.02, *se*=0.01, *p._fdr_*=0.01; statistical details in Supplementary tables ST5). Furthermore, to detect whether the causal effects of smoking initiation and bipolar disorder still existed when accounting for BMI and the significantly associated blood metabolites mentioned in this paragraph, two multivariable MR models were conducted. Smoking initiation showed a robust relationship with SmkInit#2 (*β*=-0.03, 95%CI=-0.08 - -0.004, se=0.01, *P*= 0.007) in the MVMR-IVW model, whereas bipolar disorder causal effects on MTH disappeared (P=0.084) when jointly analyzed with the BMI and candidate blood metabolites (Supplementary tables ST 7B).

Most of the sensitivity MR analyses confirmed the directions of effects detected using GSMR (Figure 3). We first discuss the results of the additional MR models that we ran as sensitivity analyses. The statistics pertinent to those results -including ORs, confidence intervals, p-values, F-statistics, I^2^ statistics and residual heterogeneity by Cochran’s Q-test can be found in Supplementary tables ST6 and ST7 (any non-significant results are highlighted in grey). As these were confirmatory analyses, we checked for consistency in the directions of effects rather than significance, so we did not correct for multiple testing. When using Inverse-Variance Weighted (IVW) and Pleiotropy RESidual Sum and Outlier (PRESSO) MR models, we obtained similarly significant causal effects (same direction of effect and nominal p values < 0.05) for all significant results from GSMR. The weighted median method, even though it yielded some non-significant results, was also consistent in all directions of effects. When using the MR Egger model, however, which is the most conservative among the models used here,^46^ we observed non-significant results and several cases where the direction of effect deviated from the consensus (i.e., the direction determined by the rest of the models, see ST6, ST7A, where non-consistent MR Egger results are written in red). All those cases were nominally non-significant (*p >* 0.05) and therefore did not affect the reliability of our results or our conclusions (STable 4). Additionally, with regards to the *I^2^* statistics of MR-Egger, there was high variability, ranging from 0 to 81.6 for TSA- and 0 to 96.7 for MTH-related causal associations. In general, for MTH we observed either very high heterogeneity (*I^2^* > 90%) or no heterogeneity at all (*I^2^*=0), whereas for TSA the I^2^ statistics were relatively moderate, typically ranging around 65%. High variability in *I^2^* statistics suggests that the studies used for those associations have diverse results or effect sizes, which might be attributed to differing study designs or populations. Thus, overall, except for some divergent, non-significant results from the MR-Egger and some convergent, non-significant results from the weighted median, the sensitivity MR methods we employed mainly yielded similar significant results as the GSMR model.

**Figure 3.**
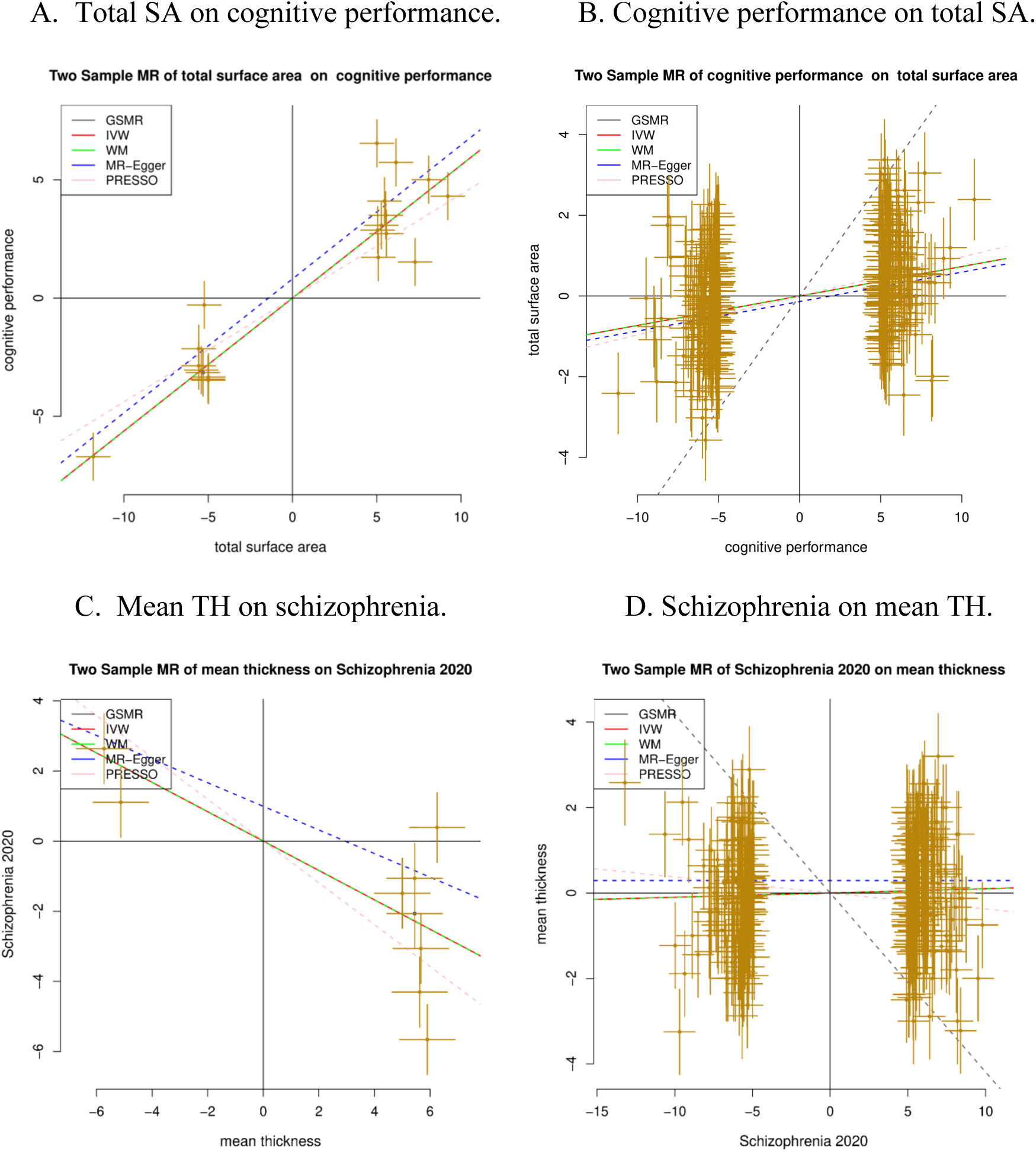
Scatter plots of bidirectional MR analyses using several models to examine causal relationships between total surface area (TSA) and cognitive performance (CP)^30^ (Fig. 3A & 3B), as well as global mean cortical thickness (MTH) and schizophrenia (freeze3, SCZ3)^41^ (Fig. 3C & 3D). (**A**) Forward MR results of TSA on CP. (**B**) Reverse MR results of CP on TSA. Based on Fig.3A and 3B, TSA and CP show a bidirectional causal relationship. (**C**): Forward MR results of MTH on SCZ3. (**D**): Reverse MR results of SCZ3 on MTH. Based on Fig. 3C and 3D, there is only a forward causal relationship between MTH and SCZ3. Note: The five models applied are all denoted: Black: GSMR, red: fixed-effect IVW, green: weighted median, blue: MR Egger, pink: MR PRESSO. The first method served as our main analysis, while the rest of the methods were used for sensitivity analyses. Before using the instruments, we detected and removed outliers with the HEIDI test. SA: total cortical surface area, TH: global mean cortical thickness, CP: cognitive performance,^30^ SCZ3: schizophrenia PGC GWAS freeze3.^41^

Next, when applying the P-value threshold of P<10e-7 for inclusion of genetic instruments to all of the significant findings in GSMR analyses, for the TSA forward analyses, 3 out of 18 causal effects had the same direction of effect but were no longer significant: CDG, nonHDL, and TC (Supplementary tables ST2). All other effects had the same direction of effect and remained significant after correction for multiple testing. In TSA reverse analyses, both positive significant causal effects of height and cognitive performance remained significant (Supplementary tables ST3). Additionally, all 5 causal effects of MTH were confirmed with the new threshold, while 9 out of 10 reverse causal effects of MTH remained similarly significant (Supplementary tables ST3&4), except for the reverse causal effects of MTH on bipolar disorder (p=0.10). As for sub-regional results, the majority of the results were the same when using the new threshold (Supplementary tables ST8-10). However, some results, particularly those related to MTH subregional corrected global thickness (Supplementary tables ST11), were no longer significant due to limited statistical power in the underlying GWASs.

Furthermore, the Cochran’s Q test in the fixed-effect IVW model and the MR-Egger model suggested that in some cases there was significant (*p* < 0.05) heterogeneity in the instrumental variables, which may be due to violation of instrumental variable assumptions. However, such findings also depend on the number of the extracted genetic instruments, so significant results in the Cochran’s Q test may also be attributed to high statistical power.

Finally, leave-one-out analyses (SFigure 4 and SFigure 5) showed that no SNPs altered the pooled IVW beta coefficient, confirming the stability of our results.

### Relationships between regional brain measures and neuropsychiatric phenotypes

#### Forward analyses for SA regions

For non-global-corrected SA (Figure 4, Supplementary tables ST8), 27 regional SA phenotypes had positive causal influences on CP,^30^ with insula as the most significant result (β=0.48, 95%CI= 0.41-0.55, se=0.04, *p._fdr_*= 2.11 x 10^-34^). After global correction (Figure 3, Supplementary tables ST9), only two regions were causally associated with a higher CP, namely insula (*β*=0.28, 95%CI= 0.19-0.38, *se*=0.05, *p._fdr_*=5.3 x 10^-5^) and transverse temporal surface area (*β*=0.24, 95%CI= 0.14-0.34, *se*=0.05, *p._fdr_*=0.002).

**Figure 4.**
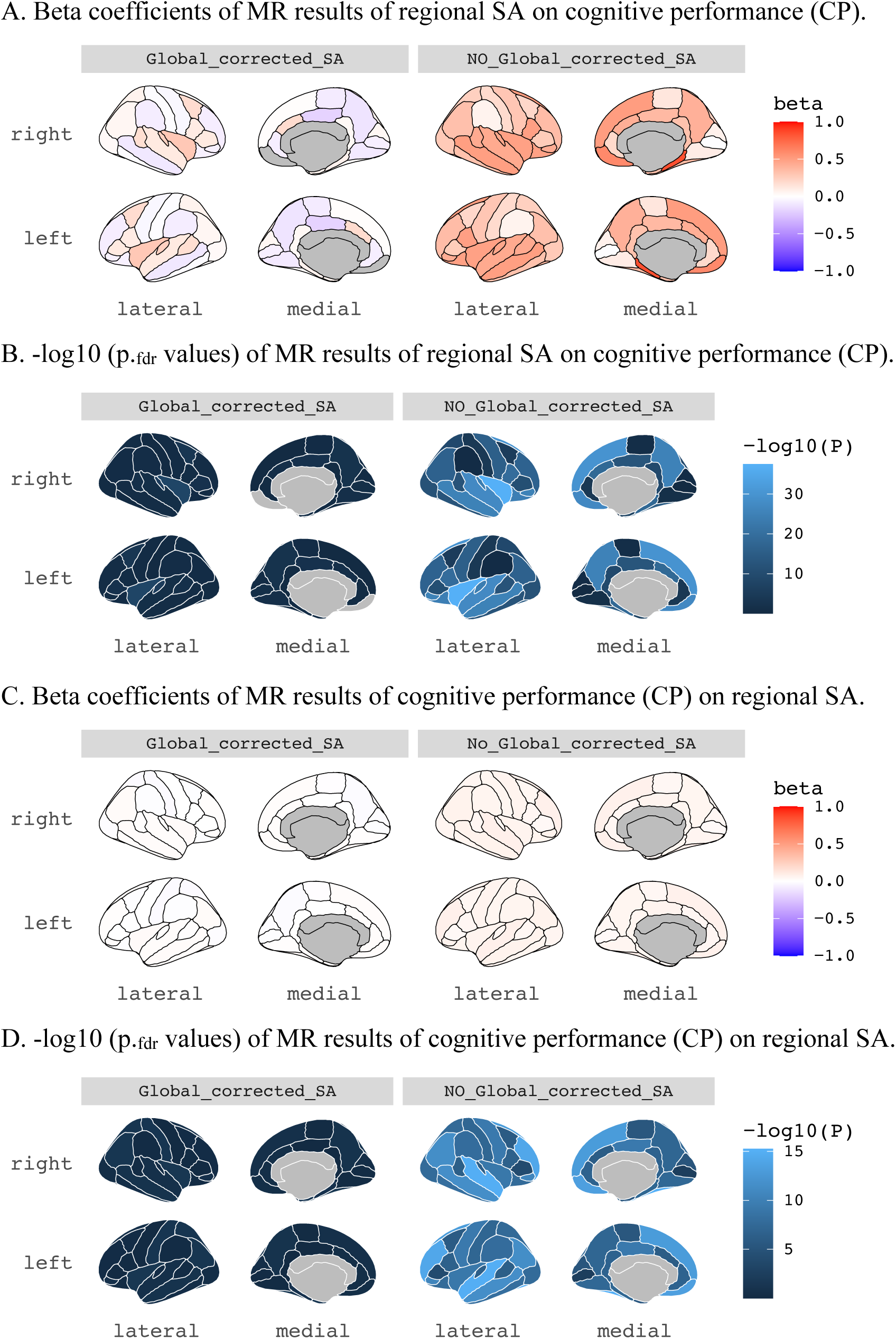
Regional plots for SA with cognitive performance (CP). Visualization in a plot of the Desikan-Killiany atlas, right hemisphere (upper) and left hemisphere (lower). Global-corrected (left) and non-global-corrected (right) MR results between SA regional measures and cognitive performance (CP). (**A**) Beta coefficients of MR results of regional SA on cognitive performance (CP). (**B**) -log10 (p._fdr_ values) of MR results of regional SA on cognitive performance (CP). (**C**) Beta coefficients of MR results of cognitive performance (CP) on regional SA. (**D**) -log10 (p._fdr_ values) of MR results of cognitive performance (CP) on regional SA. (**A & C**): the color intensity represents the strength of the causal association via beta coefficients (red: strong positive, blue: strong negative). (**B & D**): -log10 p-value after FDR-correction: the lighter the color blue, the more statistically significant the result.

Out of the 18 previously identified outcomes that are causally influenced by total SA (Figure 1A, SFigure 1A, Supplementary tables ST2), nine did not show any significant results in the respective regional analyses after global correction (Supplementary tables ST9), namely height,^32^ ADHD,^38^ Alzheimer’s disease (ALZ),^47^ age of smoking initiation 2019^37^ and 2022^34^ (SmkInit and SmkInit#2), non-high-density-lipoprotein (nonHDL),^35^ total cholesterol (TC),^35^ type II diabetes 2017^48^ and 2020^40^ (T2D2017, T2D2020). For the remaining nine outcomes, we detected some region-specific causal influences, as demonstrated in Table 1. The full results can be found in Supplementary tables ST10.

**Table 1.** Top ten significant results of the global-corrected regional MR analyses between SA regional measures and the selected phenotypes.

| *exposure* | *outcome* | *nsnps* | $\beta$ | *95% CI* | *se* | *p<sub>fdr</sub>* |
| --- | --- | --- | --- | --- | --- | --- |
| insula | CP | 14 | 0.28 | 0.19 0.38 | 0.05 | 5.30 x 10<sup>-5</sup> |
| precuneus | HDL | 22 | -0.21 | -0.28 -0.13 | 0.04 | 5.41 x 10<sup>-5</sup> |
| parahippocampal | SmkInit | 8 | 0.37 | 0.24 0.50 | 0.07 | 5.41 x 10<sup>-5</sup> |
| lingual | logTG | 17 | -0.22 | -0.30 -0.14 | 0.04 | 10.02 x 10<sup>-5</sup> |
| precuneus | EA | 21 | -0.18 | -0.26 -0.11 | 0.07 | 1.83 x 10<sup>-3</sup> |
| transverse temporal | CP | 14 | 0.24 | 0.14 0.34 | 0.05 | 1.90 x 10<sup>-3</sup> |
| lateralorbitofrontal | SmkInit | 19 | -0.18 | -0.26 -0.10 | 0.04 | 4.48 x 10<sup>-3</sup> |
| parahippocampal | logTG | 6 | 0.34 | 0.19 0.50 | 0.08 | 6.01 x 10<sup>-3</sup> |
| rostralmiddlefrontal | SmkInit#2 | 16 | -0.19 | 0.01 0.03 | 0.05 | 0.03 |
| temporal pole | CDG | 1 | -0.90 | -0.28 -0.10 | 0.23 | 0.03 |

#### Reverse analyses for SA regions

In the reverse, non-global-corrected analyses, we detected CP causal effects on 33 SA regions (Supplementary tables ST8). However, after global surface area correction (Supplementary tables ST9), all these effects were absent and the only significant reverse causal associations were height^32^ on supramarginal SA (*β*=0.02, 95%CI=0.01-0.03, *se*=0.005, *p._fdr_*=0.022) and educatioxnal attainment EA^31^ on inferior parietal SA (*β*=0.06, 95%CI=0.04-0.09, *se*=0.015, *p._fdr_*=0.037), as marked in bold in ST9. Neither of these associations are particularly strong (very small beta values). The full results can be found in Supplementary tables ST10.

#### Forward analyses for TH regions

Before global correction (Supplementary tables ST10), three regional thickness phenotypes (namely caudal middle frontal, middle temporal and inferior parietal) increased the risk of schizophrenia freeze3 (SCZ3),^41^ whereas three regional thickness phenotypes (namely caudal anterior cingulate, inferior temporal and fusiform) decreased such risk (Figure 5). As for schizophrenia freeze2 (SCZ2),^42^ it was causally influenced by only one regional measure, namely caudal middle frontal (SFigure 6). Comparing the results of those two data freezes demonstrates that the more data are available, the more causal associations can be identified, as expected (Supplementary tables ST10). However, after global thickness correction (Supplementary tables ST11), only transverse temporal thickness was associated with a lower risk of schizophrenia (SCZ3) (*β*=-1.96, 95%CI=-2.30--0.92, *se*=0.44, *p._fdr_*= 0.006). Furthermore, we detected negative causal effects of superior frontal TH and inferior temporal TH on alcoholic drinks per week 2022 (DrnkWk#2),^34^ along with negative causal effects of cuneus TH on BDSCZ.^43^ We did not detect any specific regional effects on smoking initiation 2022 (SmkInit#2)^34^ (SFigure 7). The full results can be found in Supplementary tables ST11.

**Figure 5.**
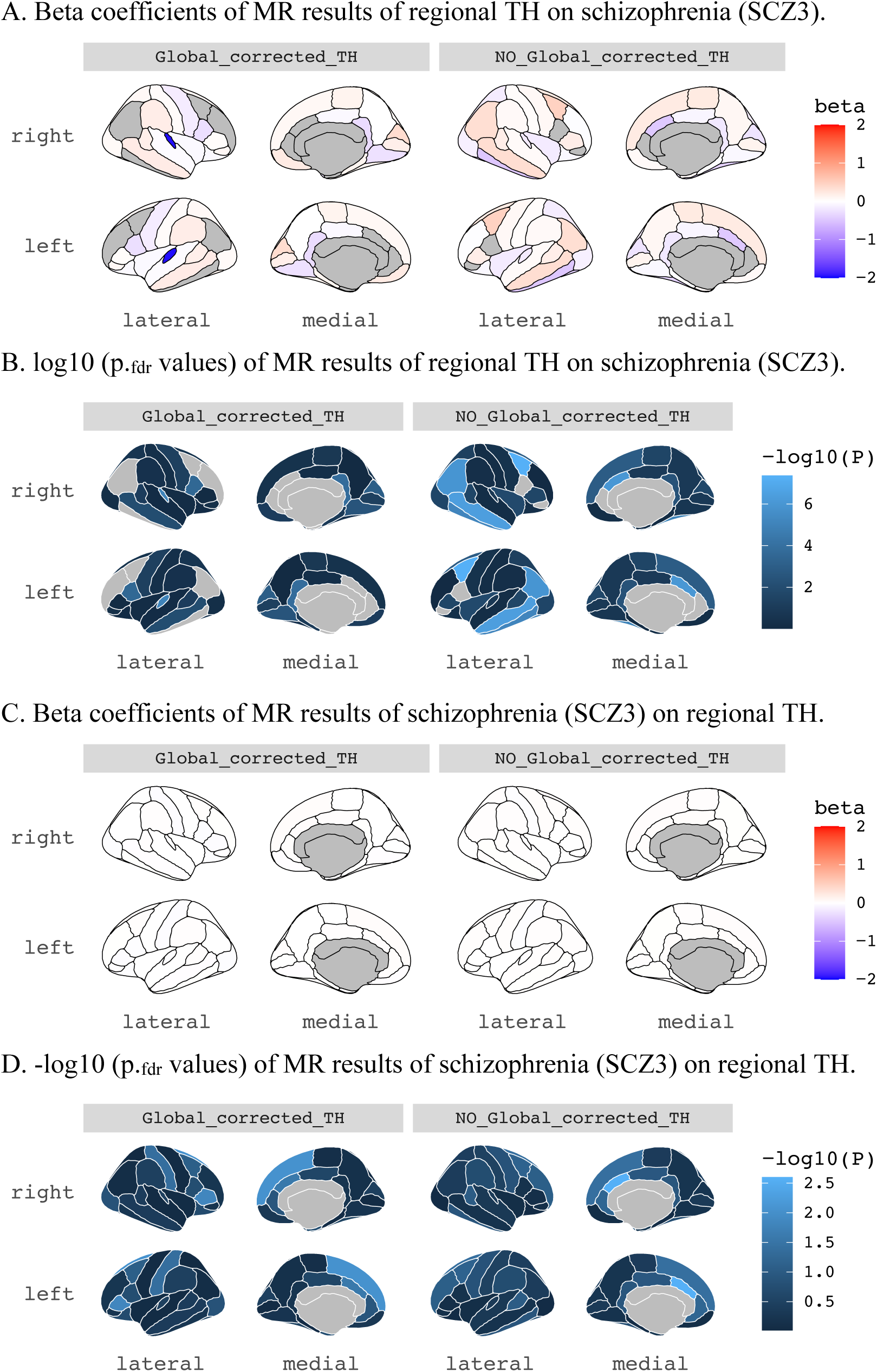
Regional plots for TH with schizophrenia freeze 3 (SCZ3). Visualization in a plot of the Desikan-Killiany atlas, right hemisphere (upper) and left hemisphere (lower). Global-corrected (left) and non-global-corrected (right) MR results between TH regional measures and SCZ3. (**A**) Beta coefficients of MR results of regional TH on schizophrenia freeze 3 (SCZ3). (**B**) -log10 (p._fdr_ values) of MR results of regional TH on schizophrenia freeze 3 (SCZ3). (**C**) Beta coefficients of MR results of schizophrenia freeze 3 (SCZ3) on regional TH. (**D**) -log10 (p._fdr_ values) of MR results of schizophrenia freeze 3 (SCZ3) on regional TH. (**A & C**): the color intensity represents the strength of the causal association via beta coefficients (red: strong positive, blue: strong negative). (**B & D**): -log10 p-value after FDR-correction: the lighter the color blue, the more statistically significant the result.

#### Reverse analyses for TH regions

We did not detect any phenotypic causal effects on any specific regional TH. The full results can be found in Supplementary tables ST11.

## Discussion

Here, by using data from over 13.5 million participants, we provide evidence that cortical morphology has causal relationships with a range of neuropsychiatric phenotypes and related biological measures. Our findings hint that variation in cortical morphology is largely causal of and not consequential to neuropsychiatric phenotypes. In addition, TSA exhibited a more widespread impact on phenotypes (*N*=18, of which 5 neuropsychiatric) than MTH (*N*=5, of which 3 neuropsychiatric). Finally, in the reverse analyses we found that fewer phenotypes (*N*=2, of which 1 neuropsychiatric) influenced TSA than MTH (*N*=10, of which 1 neuropsychiatric). Of all the detected causal relationships, several noteworthy findings include bidirectional associations between TSA, MTH and neuropsychiatric traits, as well as protective effects of TSA on cognitive performance and of MTH on schizophrenia.

The novelty of our approach and findings may be summarized as follows. First, we leveraged an unprecedented set of 199 phenotypes spanning neuropsychiatric, behavioral, and metabolic domains. This comprehensive approach allowed us to explore bidirectional causal relationships between 70 cortical morphology measures and a wide range of phenotypes, extending our understanding beyond the scope of previous MR studies. Second, we applied several statistical models to examine whether the findings align. Our use of GSMR, incorporating the heterogeneity in dependent instruments (HEIDI) method, in addition to the four sensitivity analyses, provided a stringent and powerful approach. Third, the use of cortical morphology data from the ENIGMA consortium provided significant advantages over the use of UKB data. Fourth and finally, we highlight several novel findings. We uncovered several new unidirectional and bidirectional relationships between cortical morphology and various neuropsychiatric, behavioral, and metabolic phenotypes. These findings provide new insights into the potential causal mechanisms underlying these relationships and highlight novel areas for further research.

Besides the influence of bipolar disorder (BIP) on MTH, no other psychiatric disorders seemed to affect cortical morphology. Conversely, several psychiatric traits were affected by cortical morphology, thus confirming some observational associations^5^, to which we add evidence about causality and directions of effect. Notably, we detected no causal associations for major depression disorder (MDD) and anxiety-related disorders. As those disorders show the lowest heritability among the most common psychiatric problems,^49^ possibly, insufficient GWAS hits have hitherto been detected to be extracted as instruments and help identify causal relationships.

The more widespread impact of TSA than of MTH on cortical morphology may be explained by the greater number of associated loci that have been identified for TSA than for MTH.^28,50^ These differences are likely to have been driven by the divergent evolutionary paths of the two metrics, as also captured by the radial unit hypothesis:^15,51^ TSA has expanded several orders of magnitude in humans compared to other mammals, while MTH has remained roughly the same across these species. The scale of the measures may further contribute to the difference in genetic discoverability: MTH is ∼3-4 mm, while TSA is dozens or hundreds of cubic mm per region.^52–54^ TSA is therefore more likely to have a higher signal-to-noise ratio, thus increasing chances of capturing biological processes. Notably, however, the reverse analyses showed that MTH was causally influenced by more phenotypes than TSA, suggesting that the discovered causal associations reflect true underlying mechanisms and do not result from measurement bias.

We highlight some divergent effects of TSA and MTH on a variety of neuropsychiatric disorders. For example, TSA had a positive causal effect on bipolar disorder (BIP#2) and a negative causal effect on attention-deficit/hyperactivity disorder (ADHD), and Alzheimer’s disease (ALZ). On the other hand, MTH had negative causal effects on schizophrenia (SCZ2 and SCZ3), as well as on the joint schizophrenia and bipolar disorder phenotype (BDSCZ), which likely reflects a SCZ-driven association given the high proportion of SCZ cases. With regards to BIP, our results build on largely conflicting literature. For instance, one brain imaging study ^5^ found BIP to be associated with less MTH, but not with TSA, while another study reported decreased thickness in the left superior temporal gyrus.^55,56^ Furthermore, even though the studies of BIP and BIP#2 are non-independent (BIP#2 includes all BIP samples), the former only causally influences MTH, while the latter is only causally influenced by TSA, curtailing the robustness of the BIP findings. One reason for the heterogeneity in cortical morphology findings for BIP in MR studies may relate to the two clinically and genetically different subtypes of bipolar disorder (types 1 and 2), proportions of which may vary across GWASs,^57^ as well as varying durations of illness.^58,59^ Importantly, the studies discussed here for BIP were not MR studies and therefore the associations reported may be subject to disease state influences. As for SCZ, previous research has found genetic associations of SCZ with both SA and TH, with the former association pertaining to the global level and the latter mostly to the regional level.^60^ In our study, both MTH and regional TH measures were causally associated with SCZ. In fact, all three SCZ-related phenotypes involved in this study (SCZ2, SCZ3, BDSCZ) were causally influenced by MTH in the same direction. This observation further adds to the already established associations between MTH and SCZ,^61–63^ and we now contribute evidence for negative causation. After global thickness correction, only the transverse temporal thickness showed negative causal effects on schizophrenia (SCZ3), while cuneus thickness (TH) showed negative causal effects on BDSCZ. The transverse temporal gyrus (Heschl’s gyrus) is a cortical structure that is implicated in processing incoming auditory information, with TH decreases reported to be associated with the occurrence of auditory verbal hallucinations,^64–66^ a common clinical feature of schizophrenia. Another function of the cuneus is facial emotion recognition and visual memory,^67^ both of which are often impaired in schizophrenia. For the ADHD findings that we report, most of the non-global corrected 17 regions we found to influence risk were consistent with a previous study that found smaller surface area in the frontal, temporal, and cingulate regions.^68^ However, after applying global correction, no regions remained significant. Lastly, our study revealed a causal effect of TSA on Alzheimer’s disease (ALZ). However, it is worth noting that a previous MR study did not find a causal relationship between cortical structure and ALZ.^69^ Discrepancies between the results may have originated from differences in statistical methodology and power. In general, excessive cortical thinning is a very prominent characteristic of ALZ, but it mostly impacts specific brain regions.^70^ Therefore, when conducting the broad MR analyses using the global mean of cortical thickness, it is possible that those regional changes in TH faded out in the broader measurement. However, when performing the regional analyses, ALZ was still not causally influenced by any of the regional TH measures. Conversely, several SA regions were found to have causal effects on ALZ, but only when using the non-global-corrected method. Speculatively, these findings could indicate that changes in TSA are what actually triggers the onset of ALZ, even though cortical thinning eventually takes over and shows very high associations in later stages of the disease. This does not mean, however, that cortical thinning is causally implicated in the pathogenesis of ALZ. Interestingly, a previous MR study by Wu et al.^69^ also highlights the causality of TSA on ALZ but identified different regional TSA measures as causal (precentral and isthmus cingulate). This may be because of the different ALZ GWAS summary statistics that were used, with us leveraging the results of a more recent and powerful GWAS.^39,47^

Concerning behavioral traits, TSA had positive causal effects on the age of smoking initiation and negative causal effects on ever smoking regularly. Furthermore, genetically predicted TSA increased the chances of quitting smoking. With regards to MTH, we observed a negative causal effect on the number of alcoholic drinks per week and a negative bidirectional causal effect on ever smoking regularly. Based on these findings and considering that cortical volume is mainly influenced by TSA and MTH,^71^ one may surmise that the larger the cortical volume, the healthier lifestyle choices are made. For regional MR analyses, after global correction for SA, three regions (parahippocampal, lateral orbitofrontal, and rostral middle frontal) showed significant effects on ever smoking regularly. The positive association between the surface area of the parahippocampal gyrus and regular smoking may reflect this region’s role in contextual memory and emotional processing, which may influence smoking habits. Additionally, after global correction, we observed negative causal effects of superior frontal thickness and inferior temporal thickness on weekly alcohol consumption. This aligns with previous research linking frontal lobe dysfunctions, such as impaired working memory and pattern recognition, to alcohol use. Binge drinkers often show deficits in frontal inhibitory control and executive functions.^72,73^

Through reverse MR analyses, we identified several phenotypes with causal effects on TSA and MTH, thus providing insights into modifiable factors of relevance for brain health and for clinical practice, where physicians over recent years have developed a growing awareness about lifestyle factors impacting the course of illness of many conditions.^74^ For MTH, reverse analyses yielded several actionable targets, i.e., smoking, blood lipid levels, and BMI. In the adult population, cortical thinning is a normal process of ageing, which has also been associated with numerous neuropsychiatric disorders.^75,76^ Even if cortical thinning is unavoidable, some associations indicated that the extent to and speed at which it happens can potentially be modified by external factors, e.g., physical activity, education, or diet.^77–79^ We found that regular smoking has negative bidirectional relationships with MTH, a novel finding that adds to the existing literature, which so far to our knowledge has identified only associations and no causal relationships.^80,81^ Moreover, our findings indicate a negative causal effect of BMI on MTH. Indeed, overweight individuals have reduced MTH, due to obesity-triggered neuroinflammation, which may lead to neuronal loss.^82^ A study involving children aged 4 to 18 years did not detect any significant associations between TH and BMI,^83^ suggesting that the link between BMI and cortical thinning may develop after adolescence. Additionally, we observed that several key fatty acid metabolic traits have causal effects on MTH. Polyunsaturated fatty acids (PUFAs), for instance, were found to positively affect MTH. This could potentially be because of PUFAs’ crucial role in maintaining the fluidity and permeability of neuronal membranes.^84^ In sum, our findings hint that avoiding smoking, reducing BMI, and intake of fatty acids may yield benefits for brain health.

Our study has several strengths, including a large number of phenotypes and a wide range of powerful statistical methods. These include our main analysis method (GSMR), a stringent approach,^85^ along with various extra steps to ensure reliability, e.g., a range of sensitivity analyses, the use of several GWASs for single phenotypes, and the HEIDI test. Considering that quality assessment tools have not been adequately validated and established yet,^86^ we leveraged the aforementioned strategies along with the estimation of several parameters like the F-statistics. We did, however, fill out the Strobe-MR checklist (Supplement), to show how we complied with standards in MR research. Nevertheless, the following limitations should be borne in mind when interpreting our findings. First, a general limitation of MR is that MR assumptions cannot be explicitly proven. We increased chances that such assumptions are met by using powerful and stringent statistical methods, as well as sensitivity analyses. In addition, limited GWAS sample sizes may reduce statistical power in MR research; this may have applied to some of the phenotypes under study. Thus, although by applying multiple testing correction we diminished the type I error likelihood, it is possible that for some phenotypes relevant associations were missed. Furthermore, underrepresentation of various ethnic groups in GWASs hampers the global generalizability of our results. Finally, the limited representation of diverse ethnic groups in GWAS studies, which is a general challenge in psychiatric genetics, affects the broader applicability of our findings. Genetic variants may not have the same impact across populations. Two-sample Mendelian randomization (MR) requires population matching primarily to ensure the validity and accuracy of causal inferences [for more details please see our reply to reviewer’s 2 point 1]. To minimize the risk of bias arising from population stratification, we focused on European populations. Future studies should incorporate more diverse populations to enhance the generalizability of genetic research, while novel methods may address population stratification to better understand the interplay between genetics, environment, and various phenotypes across populations. In addition, longitudinal studies are needed to confirm temporal relationships and causality between cortical morphology and related phenotypes. Moreover, investigating interactions between genetic predispositions and environmental factors such as diet, lifestyle, and socioeconomic status may provide a more comprehensive understanding of these relationships. Finally, intervention studies may examine a range of brain morphology and behavioral outcome measures of lifestyle sessions given to targeted populations, e.g., those who smoke or those with a specific blood lipids profile.

In conclusion, we show that genetically predicted variation in cortical morphology plays causal roles in neuropsychiatric disorders, particularly schizophrenia. We additionally provide new insights into effects of genetically predicted phenotypes on cortical morphology, many of which are actionable.

## Acknowledgements

We would like to acknowledge the authors of the original GWAS studies who shared (either publicly or privately) the GWAS summary statistics, encouraging data accessibility and thus scientific collaboration.

## Funding

D.M. is funded by the Research Council of Norway (#324252).

Y.L. is funded by the Natural Science Foundation of Gansu Province, China (#22JR5RA728). No funding was provided to carry out this work.

## Supplementary material

Supplementary methods, tables (STable 1-3) and figures (SFigures 1-7) can be found in SupplementaryMaterial.docx. Supplementary tables (ST1-ST11) can be found in SupplementaryData.xlsx. The STROBE-MR checklist we filled out can be found as a separate supplement.

## Data availability

All results data generated and analyzed during this study are included in the supplementary materials accompanying this manuscript. These supplementary materials provide the complete dataset necessary to interpret, verify, and extend the research presented in the article.

For any additional information or access to specific datasets beyond what is provided in the supplementary materials, reasonable requests can be made to the corresponding author.

## Code availability

The code used for data analysis is available on GitHub (https://github.com/Bochao1/Brian_NMH).

## Competing interests Statement

The authors report no competing interests.

## Figure Legends/Captions for main text figures

Of note: The dots (centre for the error bars) represent beta coefficients in MR study. The error bars represent 95% Confidence Intervals of beta coefficients. In multivariable (MVMR) analyses, the result for BIP had the same direction of effect but was no longer significant after FDR-correction for multiple testing (STable 7B). Furthermore, 2 tests had <10 instruments, namely SCZ3 (number of instruments = 9) and DrnkWk#2 (number of instruments = 8). Statistical tests were two-sided, and p-values were FDR-corrected to adjust for multiple testing.

Regarding the sample size of GWASs, ΤSA, MTH= 51,665; CP=257,828; EA= 3 million, Height=253,288, CDG= 438,997; HDL, nonHDL, TC, logTG =1,320,016; BIP#2= 413,466; AgeSmk, SmkInit = 1.2 million; T2D2020=23,326; T2D2017=659,316; ALZ=200,853; SmkCes#2, SmkInit#2, DrnkWk#2= 3.4 million; ADHD= 53,293; SCZ3=161,405; SCZ2=105,308; BDSCZ=41,653; BIP=31,710; BMI=322,154; Bis.DB.ratio, CH2.in.FA, Bis.FA.ratio, otPUFA, DB.in.FA, Crea, CH2.DB.ratio= 24,925.

**Figure 3. Scatter plots of bidirectional MR analyses using several models to examine causal relationships between total surface area (TSA) and cognitive performance (CP)**^30^ **(Fig. 3A & 3B), as well as global mean cortical thickness (MTH) and schizophrenia (freeze3, SCZ3)**^41^ **(Fig. 3C & 3D).**

A. Total SA on cognitive performance. B. Cognitive performance on total SA.

C.Note: The error bars represent 95% Confidence Intervals of beta coefficients. The five models applied are all denoted: Black: GSMR, red: fixed-effect IVW, green: weighted median, blue: MR Egger, pink: MR PRESSO. The first method served as our main analysis, while the rest of the methods were used for sensitivity analyses. Before using the instruments, we detected and removed outliers with the HEIDI test. SA: total cortical surface area, TH: global mean cortical thickness, CP: cognitive performance,^30^ SCZ3: schizophrenia PGC GWAS freeze3.^41^ Regarding the sample size of GWASs, ΤSA, MTH= 51,665; CP=257,828; SCZ3=161,405.

Statistical tests were two-sided, and p-values were FDR-corrected to adjust for multiple testing. Regarding the sample size of GWASs, SA= 51,665; CP=257,828.

Statistical tests were two-sided, and p-values were FDR-corrected to adjust for multiple testing. Regarding the sample size of GWASs, TH= 51,665; SCZ3=161,405.

## Notes

### Competing Interest Statement

The authors have declared no competing interest.

### Author Declarations

Existing public datasets were (or will be) used.

