## Supplementary material for "Dissecting causal relationships between cortical morphology and neuropsychiatric disorders: a bidirectional Mendelian randomization study"

### SUPPLEMENTARY METHODS

#### Linkage Disequilibrium Score Regression (LDSC)

LDSC was performed as an extra verification step to highlight the quality of our datasets. As mentioned in the main text, most cases (25/27) had an intercept that crosses 0, indicating no sample overlap. There were two exceptions to this observation (see ST1), namely educational attainment (EA, lower CI: 0.002, upper CI: 0.027) and cognitive performance (CP, lower CI: 0.015, upper CI: 0.044). Taking into account the large power of both studies, especially of EA (i.e., their very large sample sizes: > 3million participants for EA freeze 4 and >250,000 for CP), it is likely that some participants have shared ancestry backgrounds with those of the ENIGMA study. The more participants included in the studies, the higher the risk of potential overlap between them. This is an important factor to consider, however, judging from the lower and upper confidence intervals (CIs) as well as the observed results, even if there is a tiny overlap between studies, this has very little impact on the results and their reliability.

Finally, to further confirm our observations, we also tested the intercept of SA with itself (last row of ST1), which, as expected, was very close to 1 (lower CI: 0.93, upper CI: 0.96).

#### Multivariable MR (MVMR)

We conducted multivariable MR (MVMR) analyses using the MendelianRandomization R package to examine which phenotypes remained significant when taking into account pleiotropic effects, similarly to previously outlined methods.[1] MVRM estimates the effects of each exposure on an outcome adjusting for genetic associations between multiple phenotypes and the exposure. MVMR is an extension of MR that may be useful and reliable when three or more exposures are involved. In this study two MVMR IVW models were conducted for smoking initiation and bipolar disorder, adjusting for the exposures of BMI and 7 blood metabolic phenotypes as BMI and such metabolic traits may be biologically very closely related. For these MVMR analyses, we constructed instruments using SNPs from univariable MR tests of BMI and from the 7 blood metabolic phenotypes highlighted as significant in the main text (Figure 2D), along with smoking initiation (model 1), and bipolar disorder (model 2), as these were all the phenotypes showing associations with MTH, in addition to the metabolic quantitative traits listed as significant in this figure 2D. Then, we selected independent SNPs (n=296 for model 1; n=132 for model 2) as instruments for MVMR-IVW tests. The residual heterogeneity was detected by Cochran's Q test (and the p-value of the Q statistics).

**STable1. Cortical morphology measures as derived from the ENIGMA consortium.**
These include 2 global cortical measures (total surface area (SA) and average thickness (TH)), as well as SA and TH measures for 34 brain regions (so 68 regional measures in total).

| | SA | TH |
|---|---|---|
| **0.** | Total SA (global, TSA) | Average TH (global, MTH) |
| **1.** | Frontal Pole | |
| **2.** | Medial Orbitofrontal | |
| **3.** | Lateral Orbitofrontal | |
| **4.** | Rostral Anterior Cingulate | |
| **5.** | Caudal Anterior Cingulate | |
| **6.** | Superior Frontal | |
| **7.** | Rostral Middle Frontal | |
| **8.** | Pars Orbitalis | |
| **9.** | Pars Triangularis | |
| **10.** | Pars Opercularis | |
| **11.** | Causal Middle Frontal | |
| **12.** | Paracentral | |
| **13.** | Precentral | |
| **14.** | Postcentral | |
| **15.** | Precuneus | |
| **16.** | Superior Parietal | |
| **17.** | Supramarginal | |
| **18.** | Inferior Parietal | |
| **19.** | Posterior Cingulate | |
| **20.** | Isthmus Cingulate | |
| **21.** | Insula | |
| **22.** | Entorhinal | |
| **23.** | Parahippocampal | |
| **24.** | Fusiform | |
| **25.** | Temporal Pole | |
| **26.** | Inferior Temporal | |
| **27.** | Middle Temporal | |
| **28.** | Superior Temporal | |
| **29.** | Banks of the Superior Temporal Sulcus | |
| **30.** | Transverse Temporal | |
| **31.** | Lingual | |
| **32.** | Pericalcarine | |
| **33.** | Cuneus | |
| **34.** | Lateral Occipital | |

**STable2. Phenotypes of interest used in our study (with abbreviations), along with the data source and sample size information.**

| | Trait | Reference | Cases | Controls | Sample | Cohorts |
|---|---|---|---|---|---|---|
| **Neuropsychiatric phenotypes** (+Type II diabetes) | Alzheimer's disease (ALZ) | Wightman et al., 2021[3] | 79,360 | 121,493 | 200,853 | PGCALZ2, European ancestry, excluding23andMe & UKB |
| | Attention-deficit / hyperactivity disorder (ADHD) | Demontis et al., 2018[4] | 19,099 | 34,194 | 53,293 | "A meta-analysis restricted to European-ancestry individuals" |
| | Attention-deficit / hyperactivity disorder symptom scores (ADHD_NTR) | Middeldorp et al., 2016[5] | | | 17,666 | 11 cohorts of European descent (collected from Europe, Australia, and the United States) from EAGLE consortium. |
| | Anxiety (AN) | Otowa et al., 2016[6] | 11,600 | 33,970 | 45,570 | 7 cohorts from ANGST consortium (European ancestry) |
| | Anxiety disorders (AD) | Meier et al., 2019[7] | 19,681 | 33,970 | 53,651 | iPSYCH project (Danish sample) |
| | Agreeableness_GPC1<br>Conscientiousness_GPC1<br>Extraversion_GPC1<br>Neuroticism_GPC1<br>Openness_GPC1 | De Moor et al., 2012[8] | | | 17,375 | All participants were of European ancestry. |
| | Aggression (Agression_NTR) | Pappa et al., 2016[9] | | | 18,988 | All children were of North European ancestry. |
| | Amyotrophic lateral sclerosis (ALS) | Nicolas et al., 2018[10] | 20,806 | 59,804 | 80,610 | US+Italian+UK+French+ Belgian |

| | | | | | |
|---|---|---|---|---|---|
| Autism spectrum disorder (ASD) | Grove et al., 2019[11] | 18,382 | 27,969 | 46,351 | part of the iPSYCH project (Danish) |
| Bipolar disorder (BIP) | Stahl et al., 2019[12] | 20,352 | 31,358 | 31,710 | European descent (32 cohorts from 14 countries in Europe, North America and Australia) |
| Bipolar disorder exluding participants from the UK Biobank (BIP#2) | Mullins et al., 2021[13] | 41,917 | 371,549 | 413,466 | European descent (57 BD cohorts collected in Europe, North America and Australia) |
| Meta-analysis bipolar disorder and schizophrenia (BDSCZ) | Bipolar Disorder and Schizophrenia Working Group of the Psychiatric Genomics Consortium et al., 2018[14] | 20,129 | 21,524 | 41,653 | 'European descent' |
| Cognitive performance (CP) | Lee et al., 2018[15] | | | 257,828 | 'All cohort-level analyses were restricted to individuals of European ancestry' |
| Cross disorders Group (CDG) | Cross-Disorder Group of the Psychiatric Genomics Consortium. Electronic address and Cross-Disorder Group of the Psychiatric Genomics 2019[16] | 162,151 | 276,846 | 438,997 | "European samples" |

| | | | | | |
|---|---|---|---|---|---|
| Educational attainment (EA) | Okbay et al., 2022[17] | | | 3 million | European Ancestry excluding UKB |
| Anorexia nervosa (ED) | Watson et al., 2019[18] | 16,992 | 55,525 | 72,517 | European ancestry from 17 countries |
| Extraversion_GPC2<br>Neuroticism_GPC2 | Van den Berg et al., 2014[19] | | | 63,661 | All samples of European origin |
| Internalizing Problems (INT_NTR) | Benke et al., 2014[20] | | | 4,596 | 3 European ancestry cohorts: NTR+ Raine+generationR |
| Major depressive disorder (MDD) | Howard et al., 2019[21] | 170,756 | 329,443 | 500,199 | 33 European ancestry cohorts of the PGC exluding 23andme&UKB |
| Obsessive-compulsive disorder (OCD) | International Obsessive Compulsive Disorder Foundation Genetics Collaborative & O. C. D. Collaborative Genetics Association Studies, 2017[22] | 2,688 | 7,037 | 9,725 | Only individuals of European ancestry |
| Post-traumatic stress disorder (PTSD) | Nievergelt et al., 2019[23] | 23,212 | 151,447 | 174,659 | Only individuals of European ancestry |
| Schizophrenia freeze 2 (SCZ2) | Pardiñas et al., 2018[24] | 40,675 | 64,643 | 105,308 | 49 European cohorts |

| | | | | | | |
|---|---|---|---|---|---|---|
| | Schizophrenia freeze 3 (SCZ3) | Trubetskoy et al., 2022[25] | | | 161,405 | Meta-analysis of European ancestry cohorts |
| | Alcohol dependence (SUD_alcdep2018) | Walters et al., 2018[26] | 11,569 | 34,999 | 46,568 | European ancestry discovery GWAS |
| | Cannabis (SUD_cannabis) | Pasman et al., 2018 | 14,080 | 343,736 | 357,816 | European-ancestry cohorts, only including unrelated individuals |
| | Opioid dependence (SUD_Op_case)<br>Opioid exposure (SUD_Op_experience2018) | Polimanti et al., 2020[27] | 4,503<br>4,173 | 32,500 | 41,176 | European-ancestry cohorts* |
| | All epilepsy<br>Generalized epilepsy<br>Focal epilepsy | The International League Against Epilepsy Consortium on Complex Epilepsies[28] | 15,212 | 29,677 | 44,889 | Caucasian-only analyses* |
| | Tourette syndrome (TS) | Yu et al., 2019[29] | 4,819 | 9,488 | 14,307 | Four cohorts of European ancestry |
| | Type 2 diabetes 2020 (T2D2020) | Cai et al., 2020[30] | 9,978 | 13,348 | 23,326 | A total of 26 research centers located in eight different European countries (France, Italy, Spain, UK, the Netherlands, Germany, Sweden, and Denmark) |
| | Type 2 diabetes 2017 (T2D2017) | Xue et al., 2018[31] | 62,892 | 596,424 | 659,316 | European ancestry |
| **Behavioral phenotypes** | Age smoking initiation (AgeSmk)<br>Drinks per week (DrnkWk) | Liu et al., 2019[32] | | | 1.2million | European ancestry* |

| | | | | |
|---|---|---|---|---|
| | Ever smoked regularly (SmkInit)<br>Cigarettes per day (CigDay)<br>Smoking cessation (SmkCes) | | | |
| | Age smoking initiation (AgeSmk#2)<br>Drinks per week (DrnkWk#2)<br>Ever smoked regularly (SmkInit#2)<br>Cigarettes per day (CigDay#2)<br>Smoking cessation (SmkCes#2) | Saunders et al., 2022[33] | 3.4 million | European ancestry* |
| | Executive function and processing speed (EF_A, EF_B, EF_C, EF_D) | Ibrahim-Verbaas et al., 2016[34] | 32, 070 | European ancestry aged 45 years or older, free of dementia and clinical stroke at the time of cognitive testing from 20 cohorts |
| **Quantitative phenotypes** | C-reactive protein (CRP) | Ligthart et al., 2018[35] | 204,402 | European individuals |
| | 123 blood metabolites | Kettunen et al., 2016[36] | 24,925 | 14 cohorts from Europe |
| | DSer, Dser_Pla, Lala, LDSer, logD_Ala, logDAla_Pla, logDPro, logGly, logGly_Pla, logLAla_Pla, logLDAla_Pla, logLDPro, logLPro, LSer, LSer_Pla | Luykx et al., 2015[37] | 414 | Every participant had four grandparents born in The Netherlands or other North-Western European countries (Belgium, Germany, UK, France and Denmark) |
| | Low-density lipoprotein cholesterol (LDL-C) | Graham et al., 2021[38] | 1,320,016 | European ancestry-specific* |

| | | | | |
|---|---|---|---|---|
| | High-density lipoprotein cholesterol (HDL-C) | | | |
| | Log-transformed triglycerides (logTG) | | | |
| | Total cholesterol (TC) | | | |
| | Non-high-density lipoprotein Cholesterol (nonHDL-C) | | | |
| | Height | Wood et al 2014[39] | 253,288 | 79 studies consisting of 253,288 individuals of European ancestry |
| | BMI | Lockel et al 2015[40] | 322,154 | individuals of European descent |

Note: Some neuropsychiatric phenotypes have no case-control data, since they are measured via continuous variables (e.g., symptom or personality scores). * The GWASs were conducted using multiple ancestry samples, and we selected the summary statistics of meta-analysis with only European ancestry samples.

**STable3. Full biochemical names for the metabolic traits under study.**

| Abbreviation | Full biochemical name |
|---|---|
| **AcAce** | Acetoacetate |
| **Ace** | Acetate |
| **Ala** | Alanine |
| **Alb** | Albumin |
| **ApoA1** | ApoA1 |
| **ApoB** | ApoB |
| **Bis.DB.ratio** | Ratio of bisLallylic bonds to double bonds in lipids |
| **Bis.FA.ratio** | Ratio of bisLallylic bonds to total fatty acids in lipids |
| **bOHBut** | 3Lhydroxybutyrate |
| **CH2.in.FA** | CH2 groups in fatty acids |
| **CH2.DB.ratio** | CH2 groups to double bonds ratio |
| **Cit** | Citrate |
| **Crea** | Creatinine |
| **CRP** | C-reactive protein |
| **DB.in.FA** | Double bonds in fatty acids |
| **DHA** | 22:6, docosahexaenoic acid (DHA) |
| **DSer** | D-Serine |
| **Dser_Pla** | D-Serine in plasma |
| **Est.C** | Esterified cholesterol |
| **FALen** | Fatty acid length |
| **FAw3** | OmegaL3 fatty acids |
| **FAw6** | OmegaL6 fatty acids |
| **FAw79S** | OmegaL7 and L9 and saturated fatty acids |
| **Free.C** | Free cholesterol |
| **Glc** | Glucose |
| **Gln** | Glutamine |
| **Glol** | Glycerol |
| **Gly** | Glycine |
| **Gp** | Glycoprotein acetyls, mainly a1Lacid glycoprotein |
| **HDL-C** | High-density lipoprotein cholesterol |
| **HDL.C** | Total cholesterol in HDL |
| **HDL.D** | HDL diameter |
| **His** | Histidine |
| **IDL.C** | Total cholesterol in IDL |
| **IDL.FC** | Free cholesterol in IDL |
| **IDL.L** | Total lipids in IDL |
| **IDL.P** | Concentration of IDL particles |
| **IDL.PL** | Phospholipids in IDL |
| **IDL.TG** | Triglycerides in IDL |
| **Ile** | Isoleucine |
| **L.HDL.C** | Total cholesterol in large HDL |
| **L.HDL.CE** | Cholesterol esters in large HDL |
| **L.HDL.FC** | Free cholesterol in large HDL |
| **L.HDL.L** | Total lipids in large HDL |
| **L.HDL.P** | Concentration of large HDL particles |
| **L.HDL.PL** | Phospholipids in large HDL |
| **L.LDL.C** | Total cholesterol in large LDL |
| **L.LDL.CE** | Cholesterol esters in large LDL |
| **L.LDL.FC** | Free cholesterol in large LDL |
| **L.LDL.L** | Total lipids in large LDL |
| **L.LDL.P** | Concentration of large LDL particles |

| | |
|---|---|
| **L.LDL.PL** | Phospholipids in large LDL |
| **L.VLDL.C** | Total cholesterol in large VLDL |
| **L.VLDL.CE** | Cholesterol esters in large VLDL |
| **L.VLDL.FC** | Free cholesterol in large VLDL |
| **L.VLDL.L** | Total lipids in large VLDL |
| **L.VLDL.P** | Concentration of large VLDL particles |
| **L.VLDL.PL** | Phospholipids in large VLDL |
| **L.VLDL.TG** | Triglycerides in large VLDL |
| **LA** | 18:2, linoleic acid (LA) |
| **Lac** | Lactate |
| **Lala** | L-alanine |
| **LDL-C** | Low-density lipoprotein cholesterol |
| **LDL.C** | Total cholesterol in LDL |
| **LDL.D** | LDL diameter |
| **LDSer** | L-D-Serine, the L-enantiomer of serine |
| **Leu** | Leucine |
| **logD_Ala** | Logarithm of the distribution coefficient of Alanine |
| **logD_Ala_Pla** | Logarithm of the distribution coefficient of Alanine in plasma |
| **logDPro** | Logarithm of the distribution coefficient of Proline |
| **logGly** | Logarithm of the distribution coefficient of Glycine |
| **logGly_Pla** | Logarithm of the distribution coefficient of glycine in plasma |
| **logLAla_Pla** | Logarithm of the distribution coefficient of L-Alanine in plasma |
| **logLDAla_Pla** | Logarithm of the distribution coefficient of L-D-Alanine in plasma |
| **logLDPro** | Logarithm of the distribution coefficient of L-D-Proline |
| **logLPro** | Logarithm of the distribution coefficient of L-Proline |
| **logTG** | Log-transformed triglycerides |
| **LSer** | L-Serine |
| **LSer_Pla** | L-Serine in plasma |
| **M.HDL.C** | Total cholesterol in medium HDL |
| **M.HDL.CE** | Cholesterol esters in medium HDL |
| **M.HDL.FC** | Free cholesterol in medium HDL |
| **M.HDL.L** | Total lipids in medium HDL |
| **M.HDL.P** | Concentration of medium HDL particles |
| **M.HDL.PL** | Phospholipids in medium HDL |
| **M.LDL.C** | Total cholesterol in medium LDL |
| **M.LDL.CE** | Cholesterol esters in medium LDL |
| **M.LDL.L** | Total lipids in medium LDL |
| **M.LDL.P** | Concentration of medium LDL particles |
| **M.LDL.PL** | Phospholipids in medium LDL |
| **M.VLDL.C** | Total cholesterol in medium VLDL |
| **M.VLDL.CE** | Cholesterol esters in medium VLDL |
| **M.VLDL.FC** | Free cholesterol in medium VLDL |
| **M.VLDL.L** | Total lipids in medium VLDL |
| **M.VLDL.P** | Concentration of medium VLDL particles |
| **M.VLDL.PL** | Phospholipids in medium VLDL |
| **M.VLDL.TG** | Triglycerides in medium VLDL |
| **MUFA** | MonoLunsaturated fatty acids |
| **nonHDL-C** | Non-high-density lipoprotein cholesterol |
| **otPUFA** | Other polyunsaturated fatty acids than 18:2 |
| **PC** | Phosphatidylcholine and other cholines |
| **Phe** | Phenylalanine |
| **Pyr** | Pyruvate |
| **S.HDL.L** | Total lipids in small HDL |
| **S.HDL.P** | Concentration of small HDL particles |
| **S.HDL.TG** | Triglycerides in small HDL |

| | |
|---|---|
| **S.LDL.C** | Total cholesterol in small LDL |
| **S.LDL.L** | Total lipids in small LDL |
| **S.LDL.P** | Concentration of small LDL particles |
| **S.VLDL.C** | Total cholesterol in small VLDL |
| **S.VLDL.FC** | Free cholesterol in small VLDL |
| **S.VLDL.L** | Total lipids in small VLDL |
| **S.VLDL.P** | Concentration of small VLDL particles |
| **S.VLDL.PL** | Phospholipids in small VLDL |
| **S.VLDL.TG** | Triglycerides in small VLDL |
| **Serum.C** | Serum total cholesterol |
| **Serum.TG** | Serum total triglycerides |
| **SM** | Sphingomyelins |
| **TC** | Total cholesterol |
| **TGs** | Triglycerides |
| **Tot.FA** | Total fatty acids |
| **TotPG** | Total phosphoglycerides |
| **Tyr** | Tyrosine |
| **Urea** | Urea |
| **Val** | Valine |
| **VLDL.D** | VLDL diameter |
| **XL.HDL.C** | Total cholesterol in very large HDL |
| **XL.HDL.CE** | Cholesterol esters in very large HDL |
| **XL.HDL.FC** | Free cholesterol in very large HDL |
| **XL.HDL.L** | Total lipids in very large HDL |
| **XL.HDL.P** | Concentration of very large HDL particles |
| **XL.HDL.PL** | Phospholipids in very large HDL |
| **XL.HDL.TG** | Triglycerides in very large HDL |
| **XL.VLDL.L** | Total lipids in very large VLDL |
| **XL.VLDL.P** | Concentration of very large VLDL particles |
| **XL.VLDL.PL** | Phospholipids in very large VLDL |
| **XL.VLDL.TG** | Triglycerides in very large VLDL |
| **XS.VLDL.L** | Total lipids in very small VLDL |
| **XS.VLDL.P** | Concentration of very small VLDL particles |
| **XS.VLDL.PL** | Phospholipids in very small VLDL |
| **XS.VLDL.TG** | Triglycerides in very small VLDL |
| **XXL.VLDL.L** | Total lipids in chylomicrons and extremely large VLDL |
| **XXL.VLDL.P** | Concentration of chylomicrons and extremely large VLDL particles |
| **XXL.VLDL.PL** | Phospholipids in chylomicrons and extremely large VLDL |
| **XXL.VLDL.TG** | Triglycerides in chylomicrons and extremely large VLDL |

### SUPPLEMENTARY RESULTS

#### GSMR test using $P<10^{-7}$ instruments

In the TSA forward analyses (Supplementary Data STable 2), we successfully validated 15 out of 18 causal effects using a new threshold, with exceptions noted for CDG, nonHDL, and TC, where directions of effect remained the same but the significance slightly decreased. Notably, significant causal effects observed for CP and height on TSA persisted in the TSA reverse analyses (Supplementary Data Stable 3). Additionally, in Supplementary Data Stable 2, all 5 causal effects of MTH were confirmed with the new threshold, while 9 out of 10 reverse causal effects of MTH were validated, except for the reverse causal effects of MTH on bipolar, where directions of effect remained the same but the significance slightly decreased. Regarding sub-regional results, the majority were validated with the new thresholds (Supplementary Data Stable 8-10). However, some results, particularly those related to MTH subregional correct global thickness (Supplementary Data Stable 11), could not be validated with more stringent thresholds due to limited statistical power in the underlying GWASs.

#### Multivariable MR results

In model 1, we used 296 independent SNPSs associated (p<10-6) with at least one exposure as instruments for MVMR-IVW. The results showed the causal effects of smoking initiation on MTH had the same direction of effect (beta=-0.032, P=0.07); so did the causal effects of BMI (beta =-0.041, p=0.031) and Bis.DB.ratio (beta=0.335, P=0.025). We detected significant heterogeneity (Qstat=340.298 and P Qstat=0.0167) in this model.

In model 2, we used 132 independent SNPSs associated (p<10-6) with at least one exposure as instruments for MVMR-IVW. The results showed the causal effects of bipolar disorders on MTH vanished, whereas the causal effects of BMI (beta =-0.042, p=0.01), CH2.in.FA(beta =-0.328, p=0.023) and Crea (beta =0.186, p=0.012) remained. We detected significant heterogeneity (Qstat=168.944 and P Qstat=0.004) in this model.

**STable4. All MR-Egger results with inconsistent directions of effect as compared to the consensus direction (as determined by the rest of the models). None of these results is statistically significant.**

| exposure | outcome | nsnps | beta | se | p |
|---|---|---|---|---|---|
| TSA | SmkInit#2 | 14 | 0.071 | 0.739 | 0.69 |
| TSA | AgeSmk#2 | 19 | -0.005 | 0.599 | 0.96 |
| TSA | BIP#2 | 27 | -0.036 | 0.567 | 0.65 |
| MTH | SmkInit | 10 | 0.286 | 0.707 | 0.70 |
| MTH | BDSCZ | 10 | 0.136 | 0.418 | 0.75 |
| BIP | MTH | 35 | -0.163 | 0.242 | 0.51 |

**exposure:** exposure of interest in the two-sample MR

**outcome:** outcome of interest in the two-sample MR

**direction:** the direction of the MR analysis (forward: regional SA measures on other phenotypes, reverse: other phenotypes on regional SA measures)

**nsnps:** number of the extracted genetic instruments, i.e., the independent ($r^2 < 0.01$) single-nucleotide polymorphisms (SNPs) with $p < 10^{-6}$

**beta:** beta value/coefficient (the degree of change in the outcome variable for every 1 unit of change in the exposure-predictor variable)

**se:** standard error of the beta value

**p:** p-value of the potential causal association. Statistical tests were two-sided. We consider the association significant if $p < 0.05$

**SFigure 1. Forest plots of GSMR models with significant results ($p_{.fdr} < 0.05$) for total SA and global mean TH.**

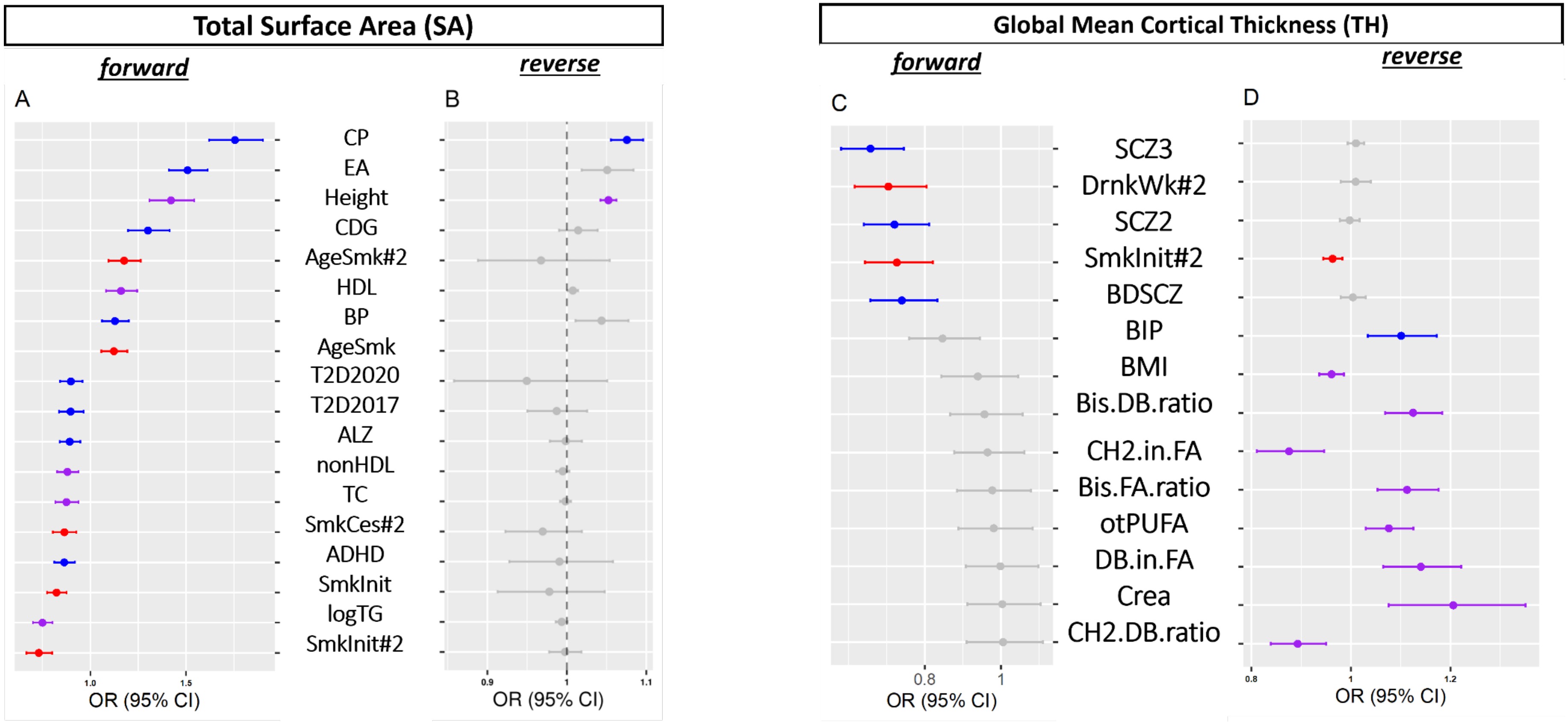

Figure 1A: Forward MR results of total SA on outcomes. Figure 1B: Reverse MR results of outcomes on total SA. Figure 1C: Forward MR results of global mean TH on outcomes. Figure 1D: Reverse MR results of outcomes on global mean TH. The significant results ($p_{.fdr}<0.05$) were colored by category: **red**= behavior, **purple**=quantitative phenotype, and **blue**=(neuro)psychiatric trait or disorder. The non-significant results ($p_{.fdr} > 0.05$) were colored in grey. In figure 1B, there were not enough significantly associated SNPs with the age of smoking initiation 2019 (AgeSmk) to be extracted as instruments, therefore the result is not available. Statistical tests were two-sided, and p-values were FDR-correction adjusted for multiple testing.

Panels A, B:

CP: cognitive performance, EA: educational attainment, Heigh), CDG: cross disorder group, AgeSmk#2: age of smoking initiation (2022), HDL: high-density lipoprotein cholesterol, BIP#2: bipolar disorder excluding participants from the UK biobank (2021), AgeSmk: age of smoking initiation (2019), T2D2020: type II diabetes (2020), T2D2017: type II diabetes (2017), ALZ: Alzheimer's disease, nonHDL: non-high-density lipoprotein cholesterol, TC: total cholesterol, SmkCes#2: smoking cessation (2022), ADHD: attention deficit hyperactivity disorder, SmkInit: ever smoked regularly (2019), logTG: log-transformed triglycerides, SmkInit#2: ever smoked regularly (2022)

Panels C, D:

SCZ3: schizophrenia freeze 3, DrnkWk#2: drinks per week (2022), SCZ2: schizophrenia freeze 2, SmkInit#2: ever smoked regularly (2022), BDSCZ: meta-analysis bipolar disorder and schizophrenia, BIP: bipolar disorder (2019), BMI: body mass index, Bis.DB.ratio: ratio of bisLallylic bonds to double bonds in lipids, CH2.in.FA: CH2 groups in fatty acids, Bis.FA.ratio: ratio of bisLallylic bonds to total fatty acids in lipids, otPUFA: other polyunsaturated fatty acids than 18:2, DB.in.FA: double bonds in fatty acids, Crea: creatinine, CH2.DB.ratio: CH2 groups to double bonds ratio

Note: The dots (centre for the error bars) represent beta coefficients in MR study. The error bars represent 95% Confidence Intervals of beta coefficients. Statistical tests were two-sided, and p-values were FDR-corrected to adjust for multiple testing.

Regarding the sample size of GWASs, TSA, MTH= 51,665; CP=257,828; EA= 3 million, Height=253,288, CDG= 438,997; HDL, nonHDL , TC, logTG =1,320,016; BIP#2= 413,466; AgeSmk, SmkInit = 1.2 million; T2D2020=23,326; T2D2017=659,316; ALZ=200,853; SmkCes#2, SmkInit#2, DrnkWk#2= 3.4 million; ADHD= 53,293; SCZ3=161,405; SCZ2=105,308; BDSCZ=41,653; BIP=31,710; BMI=322,154; Bis.DB.ratio, CH2.in.FA, Bis.FA.ratio, otPUFA, DB.in.FA, Crea, CH2.DB.ratio= 24,925.

#### SFigure 2. Scatter plots of bidirectional MR analyses between total SA and significantly associated phenotypes ($p_{fdr} < 0.05$).

Forward analyses of total SA on significant phenotypes (left) (ST2) and their respective reverse analyses (right) (ST3). Judging from the scatter plots, most causal effects are forward (i.e., the total SA causally influences the phenotypes) and there are no exclusively reverse causal associations. Finally, there are only two bidirectional relationships, namely total SA-cognitive performance (A) and total SA-Height (C).

Note: 1) The reverse analysis for the age of smoking initiation 2019 (AgeSmk) on total SA could not be conducted because there were not enough independent genome-wide significant loci (n>=2) to be extracted as instruments. 2) The five models applied are all denoted. Black: GSMR, red: fixed-effect IVW, green: weighted median, blue: MR Egger, pink: MR PRESSO. The first method served as our main analysis, while the rest of the methods were used for sensitivity analyses. Before using the instruments, we detected and removed outliers with the HEIDI test. The dots (centre for the error bars) represent beta coefficients in MR study. The error bars represent 95% Confidence Intervals of beta coefficients. Statistical tests were two-sided, and p-values were FDR-correction adjusted for multiple testing. Regarding the sample size of GWASs, TSA= 51,665; CP=257,828; EA= 3 million, Height=253,288, CDG= 438,997; HDL, nonHDL, TC, logTG =1,320,016; BIP#2= 413,466; AgeSmk, SmkInit = 1.2 million; T2D2020=23,326; T2D2017=659,316; ALZ=200,853; SmkCes#2, SmkInit#2, DrnkWk#2= 3.4 million; ADHD= 53,293.

##### A. Total SA on CP

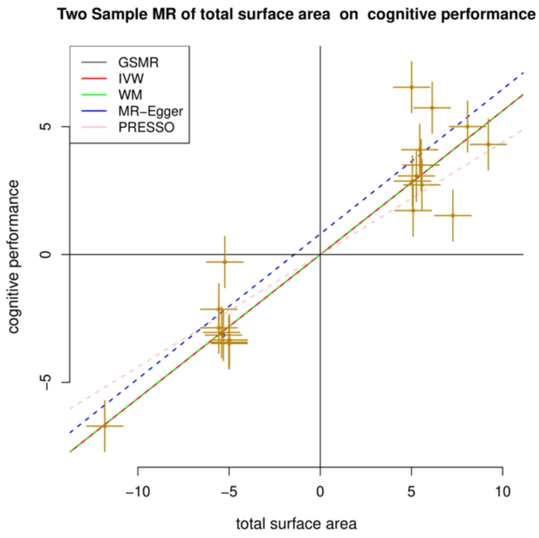

##### CP on total SA

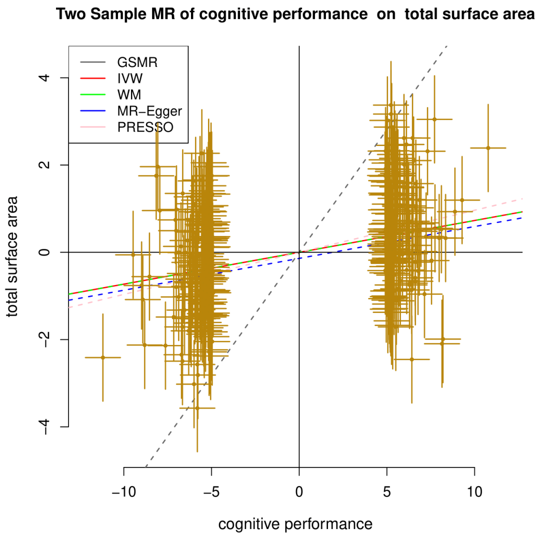

##### B. Total SA on EA

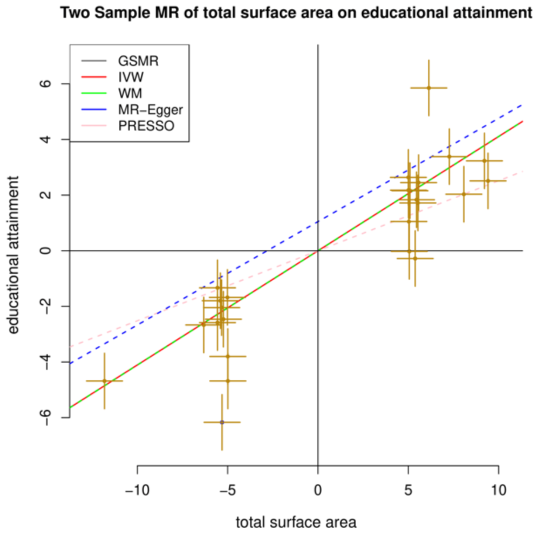

##### EA on total SA

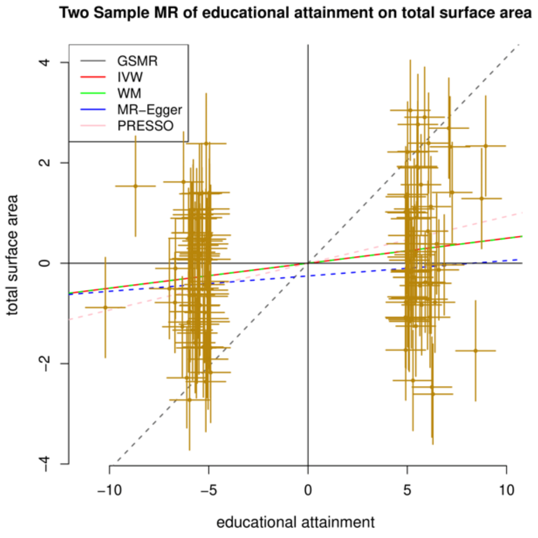

### C. Total SA on height

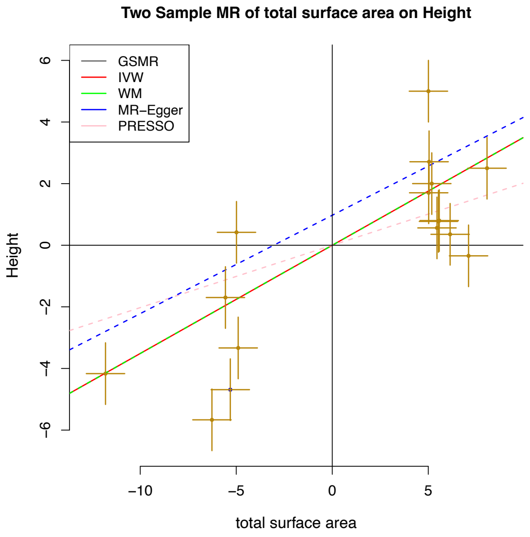

### height on total SA

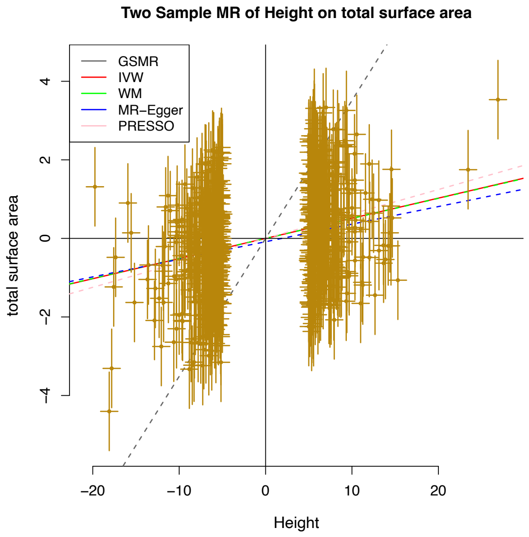

### D. Total SA on logTG

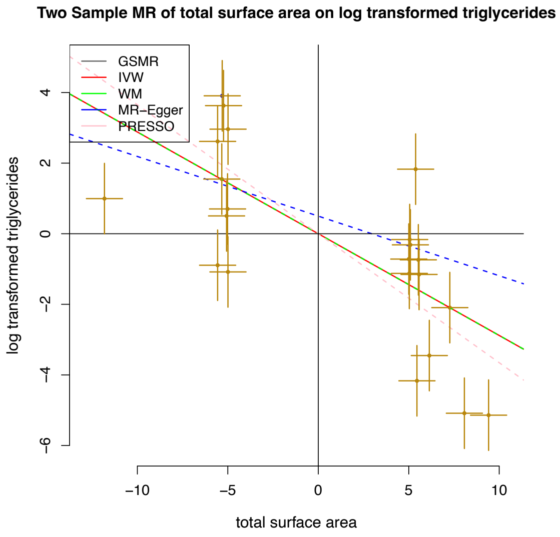

### logTG on total SA

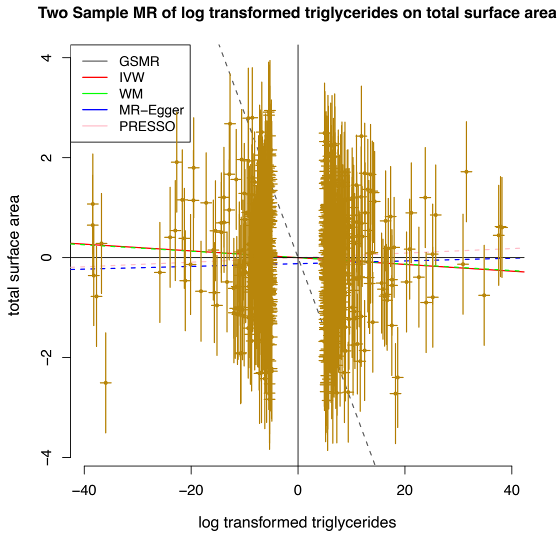

### E. Total SA on SmkInit#2

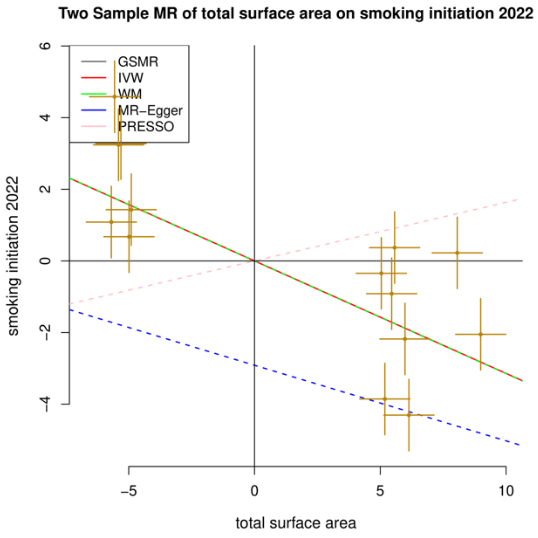

### SmkInit#2 on total SA

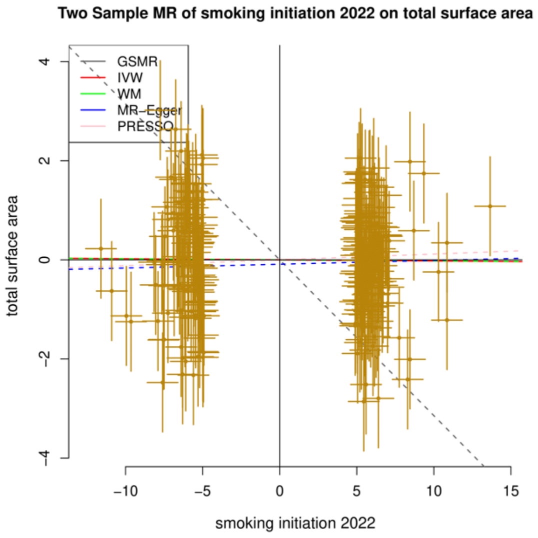

#### F. Total SA on SmkInit

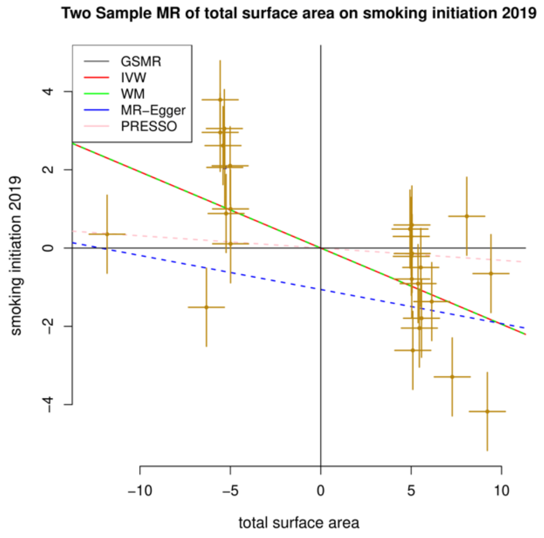

#### SmkInit on total SA

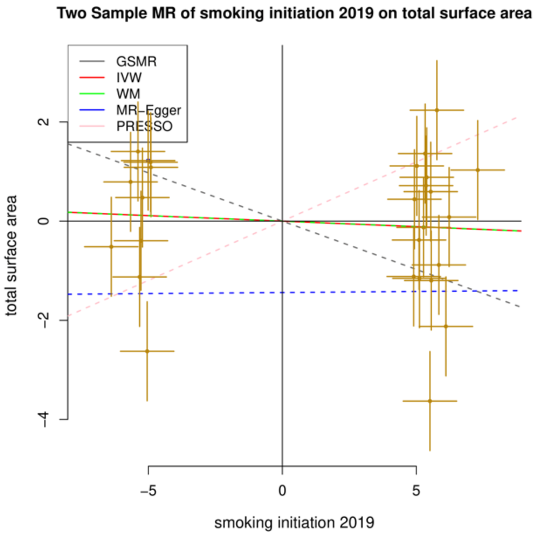

#### G. Total SA on CDG

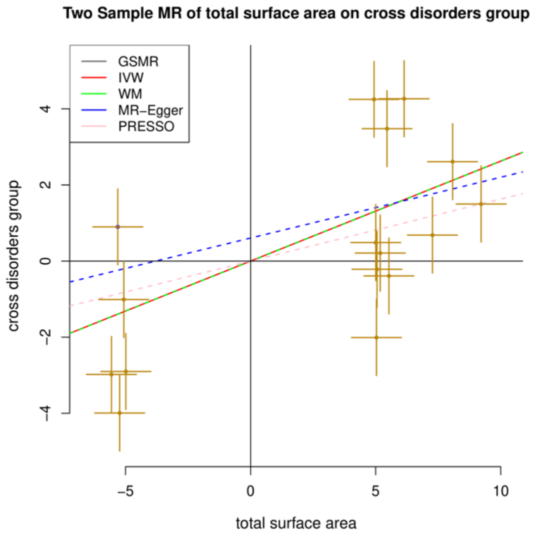

#### CDG on total SA

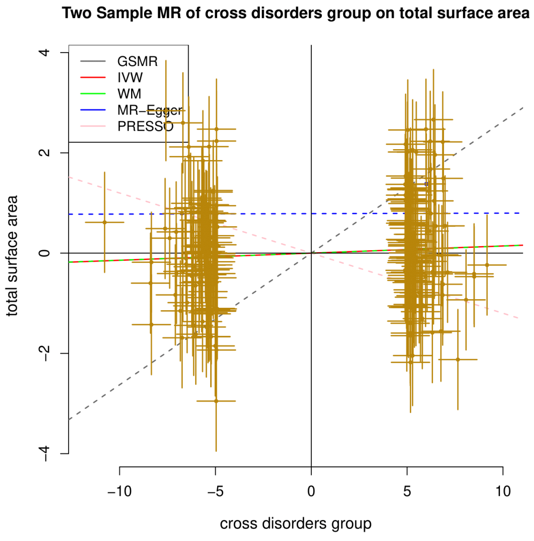

#### H. Total SA on ADHD

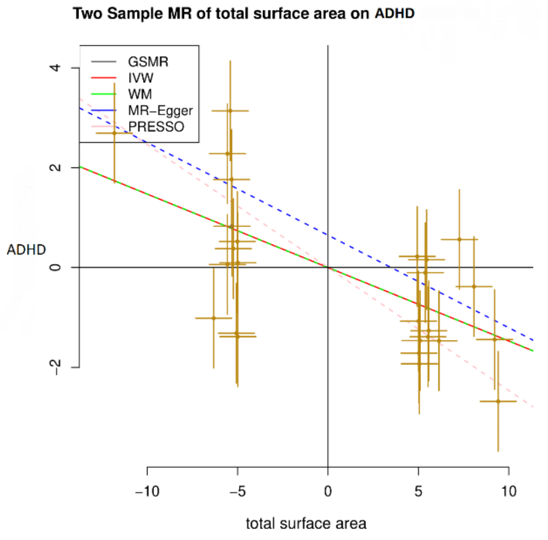

#### ADHD on total SA

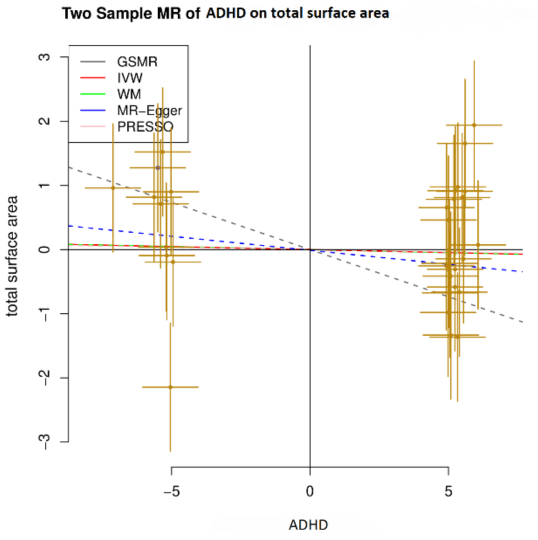

#### I. Total SA on AgeSmk#2

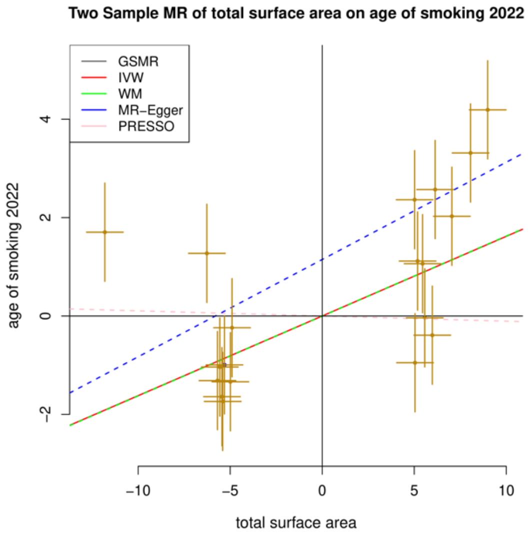

#### AgeSmk#2 on total SA

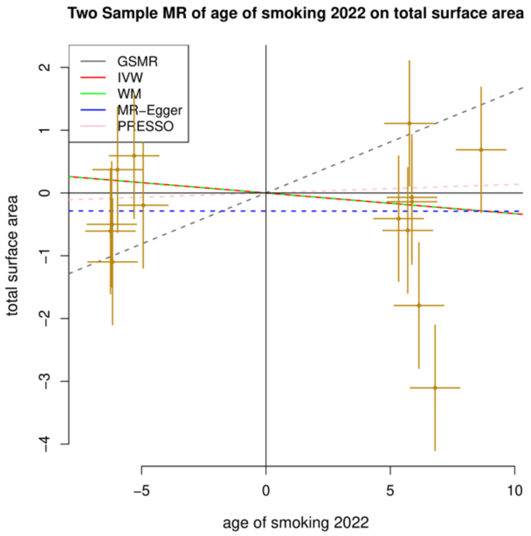

#### J. Total SA on HDL

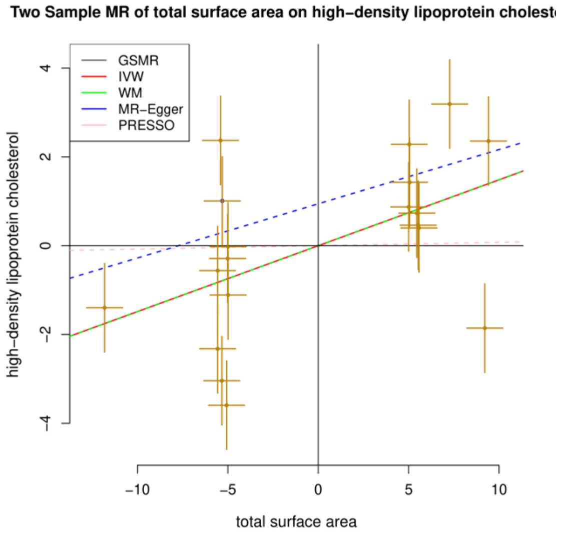

#### HDL on total SA

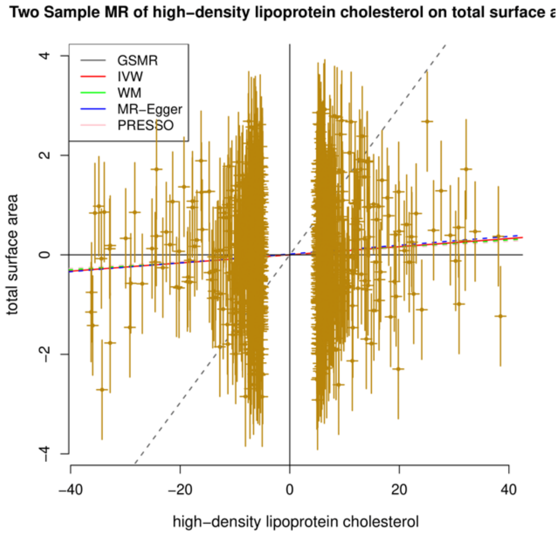

#### K. Total SA on SmkCes#2

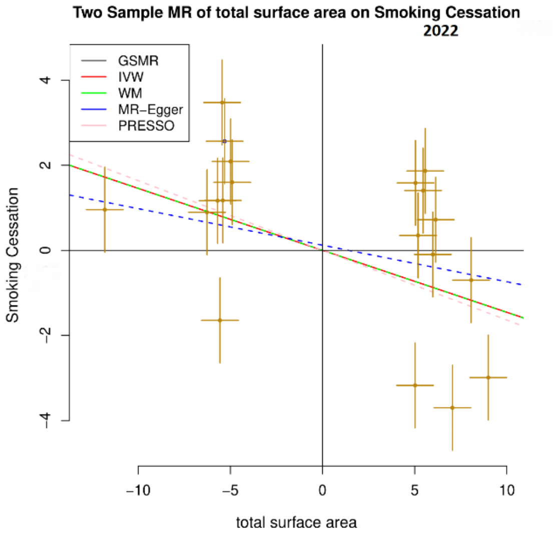

#### SmkCes#2 on total SA

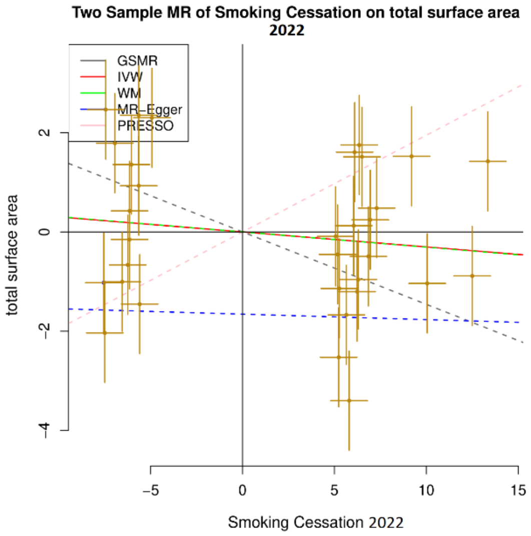

#### L. Total SA on non-HDL

#### non-HDL on total SA

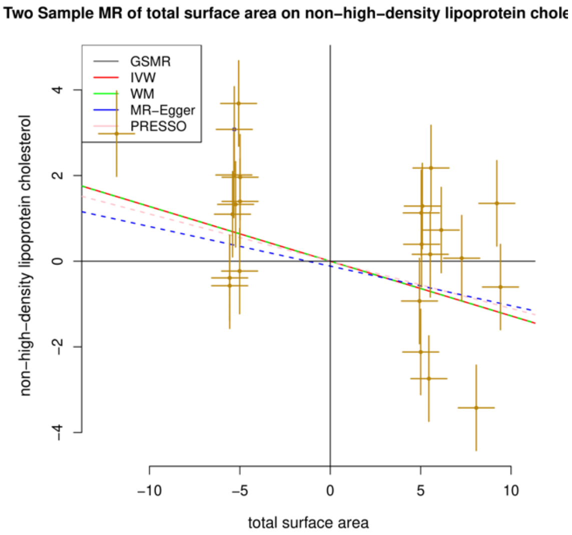

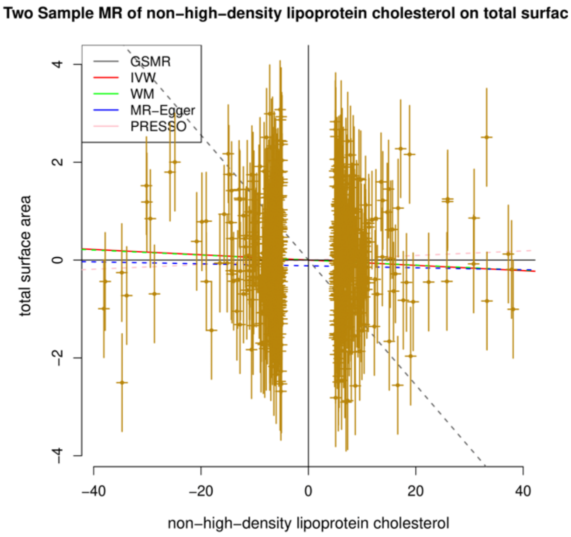

#### M. Total SA on BIP#2

#### BIP#2 on total SA

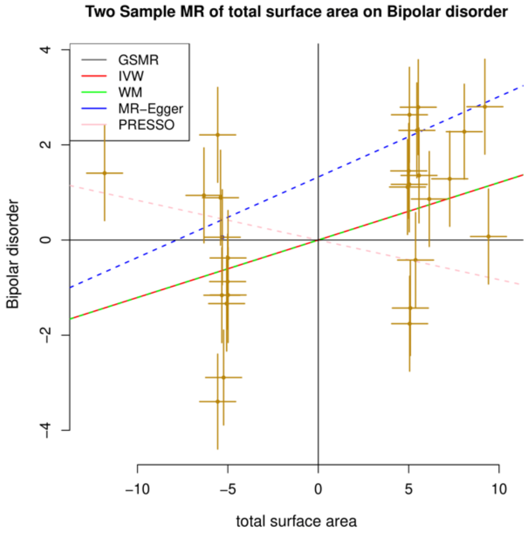

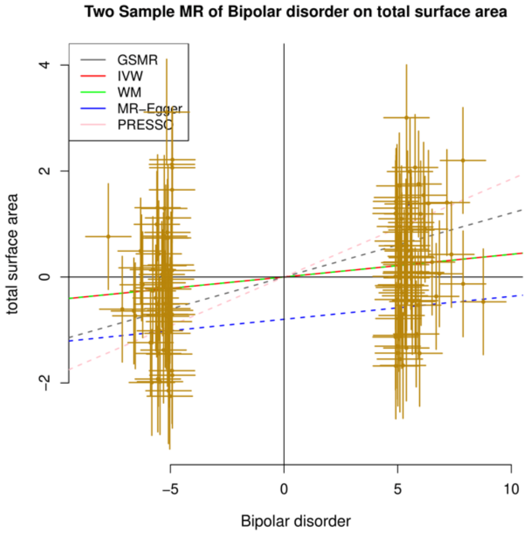

#### N. Total SA on TC

#### TC on total SA

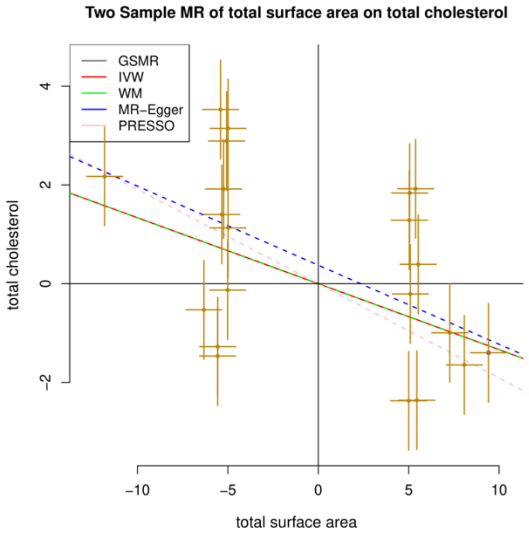

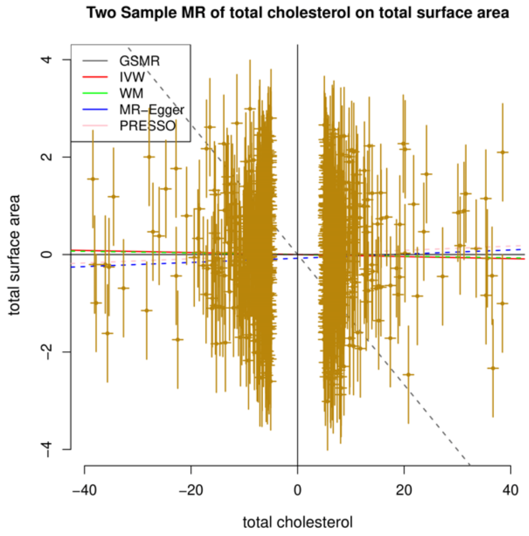

#### O. Total SA on AgeSmk

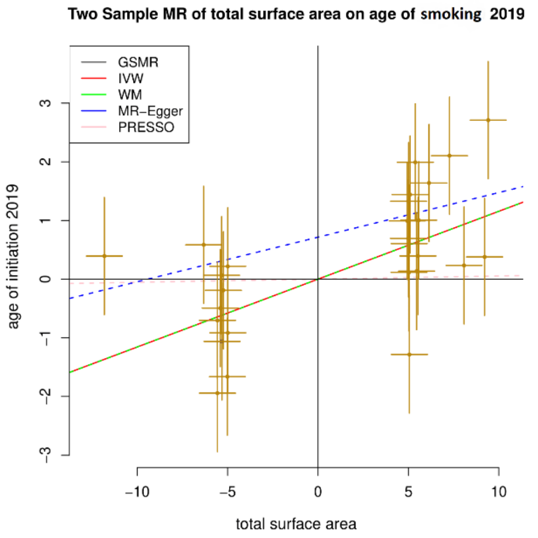

#### AgeSmk on Total SA

This phenotype did not have enough independent SNPs to be extracted as instruments and therefore we could not perform reverse MR analyses on total SA.

#### P. Total SA on ALZ

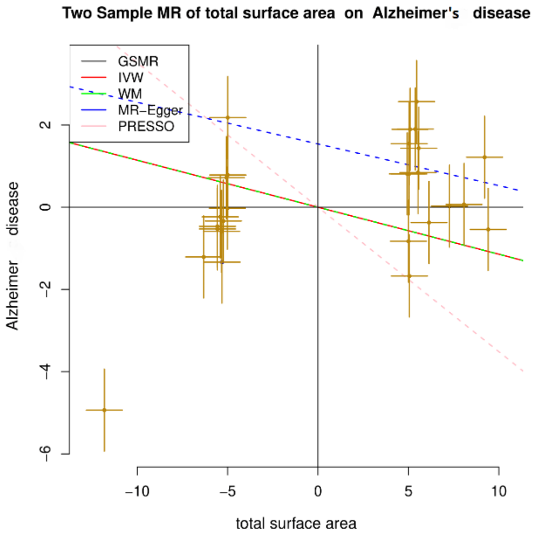

#### ALZ on total SA

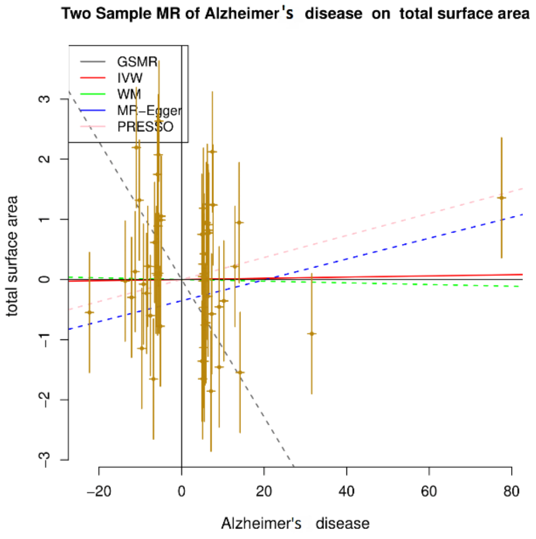

#### Q. Total SA on T2D2020

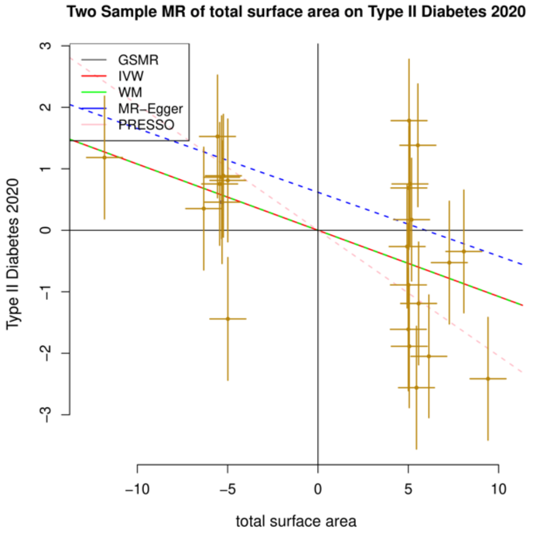

#### T2D2020 on total SA

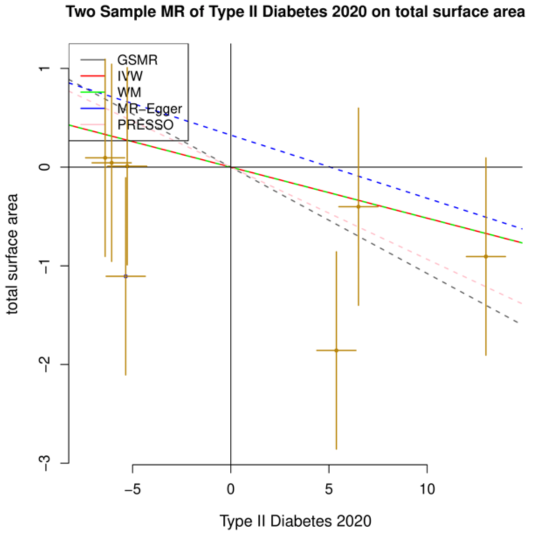

**R. Total SA on T2D2017**

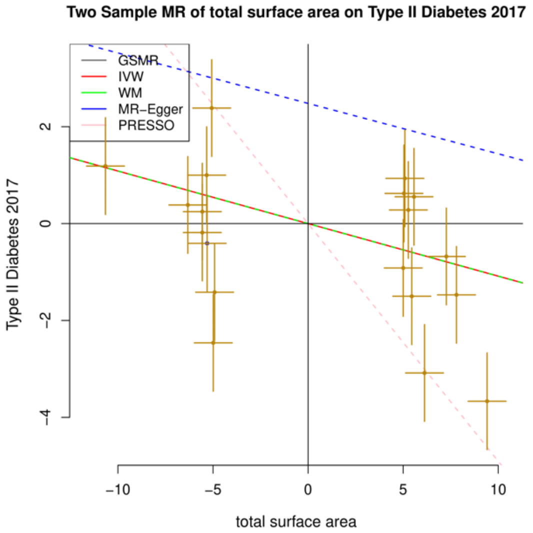

**T2D2017 on total SA**

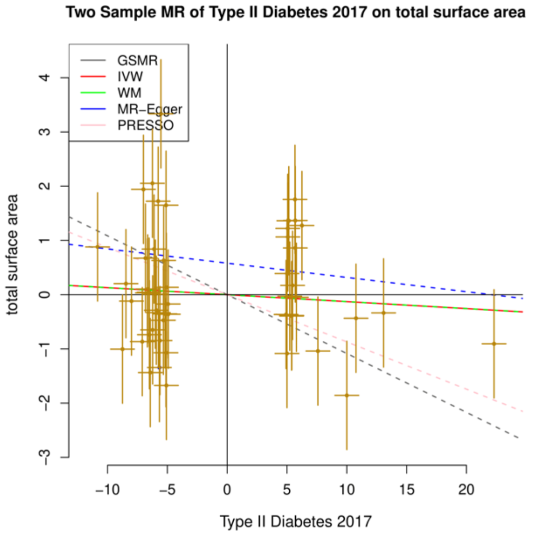

#### SFigure 3. Scatter plots of bidirectional MR analyses between global mean TH and significantly associated phenotypes ($p_{fdr} < 0.05$).

Forward analyses of global mean TH on phenotypes (left) (ST4) and the respective reverse analyses (right) (ST5). Judging from the scatter plots, four causal relationships are exclusively forward, nine are exclusively reverse and one is bidirectional: global mean TH has a negative causal relationship with smoking initiation (SmkInit#2) and vice versa (bidirectional relationship - Panel D).

Note: The five models applied are all denoted. Black: GSMR, red: fixed-effect IVW, green: weighted median, blue: MR Egger, pink: MR PRESSO. The first method served as our main analysis, while the rest of the methods were used for sensitivity analyses. Before using the instruments, we detected and removed outliers with the HEIDI test. The dots (centre for the error bars) represent beta coefficients in MR study. The error bars represent 95% Confidence Intervals of beta coefficients. Regarding the sample size of GWASs, MTH= 51,665; SmkCes#2, SmkInit#2, DrnkWk#2= 3.4 million; SCZ3=161,405; SCZ2=105,308; BDSCZ=41,653; BIP=31,710; BMI=322,154; Bis.DB.ratio, CH2.in.FA, Bis.FA.ratio, otPUFA, DB.in.FA, Crea, CH2.DB.ratio= 24,925.

##### A. Global mean TH on SCZ3

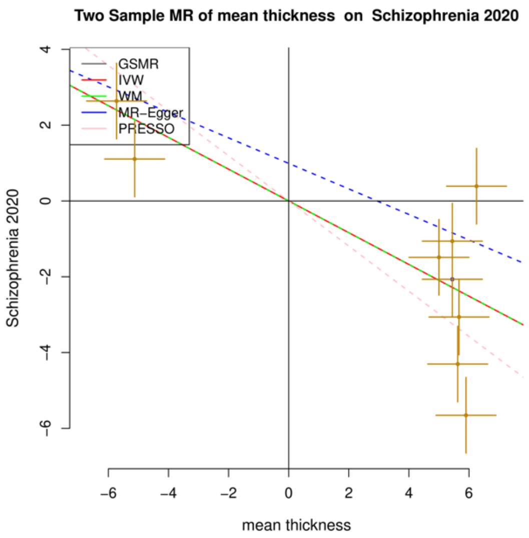

##### SCZ3 on global mean TH

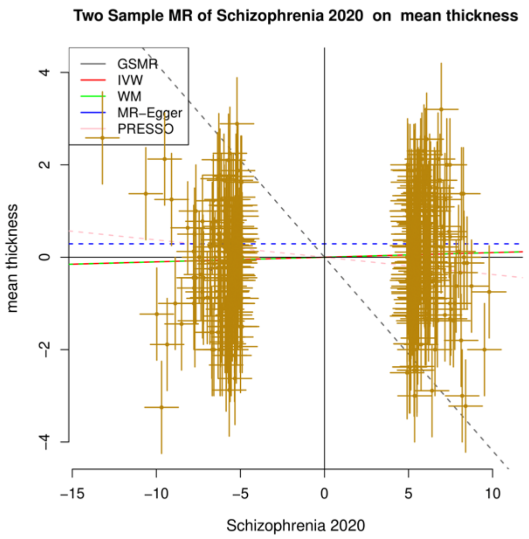

##### B. Global mean TH on SCZ2

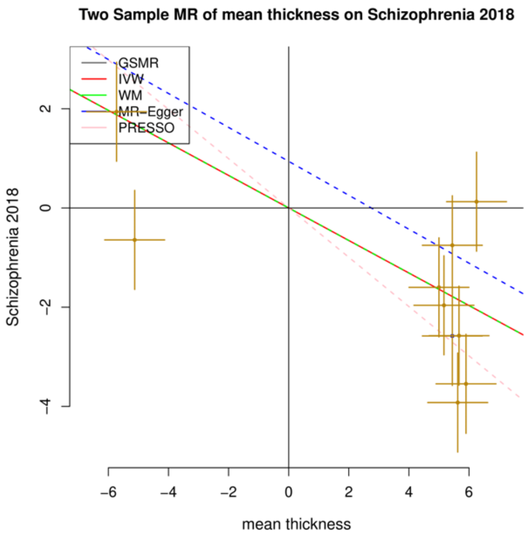

##### SCZ2 on global mean TH

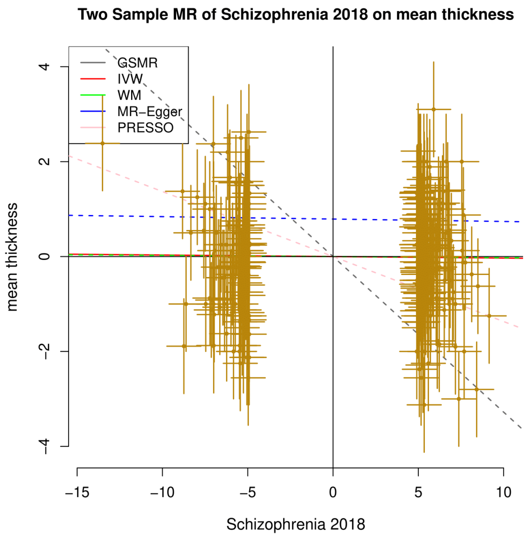

#### C. Global mean TH on DrnkWk#2

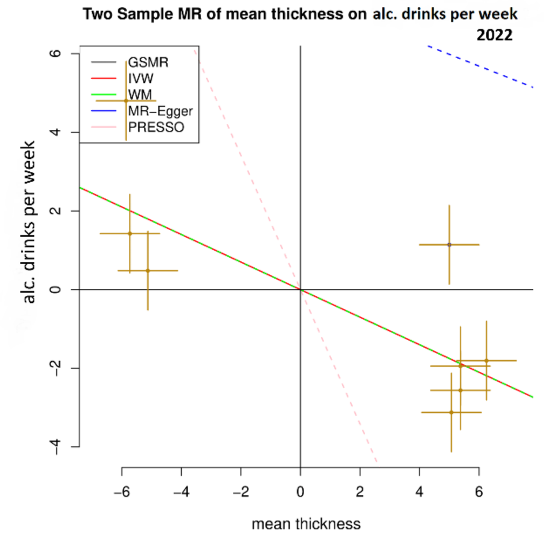

#### DrnkWk#2 on global mean TH

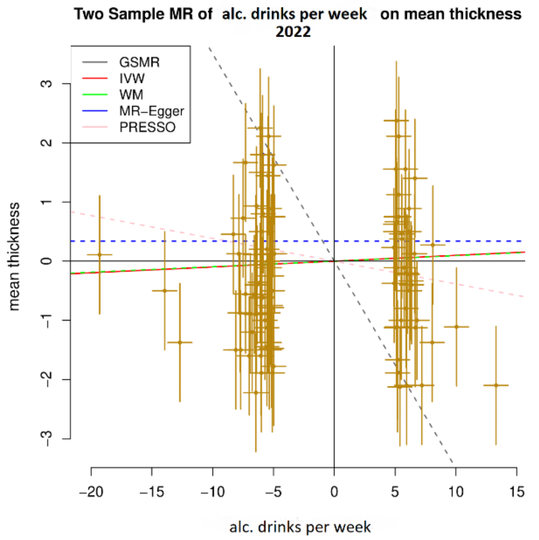

#### D. Global mean TH on SmkInit#2

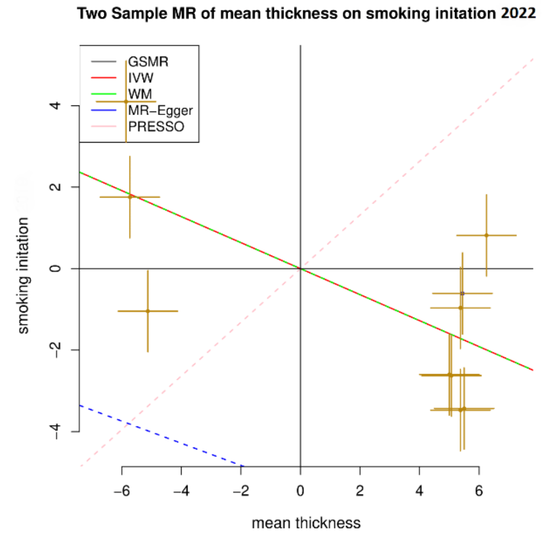

#### SmkInit#2 on global mean TH

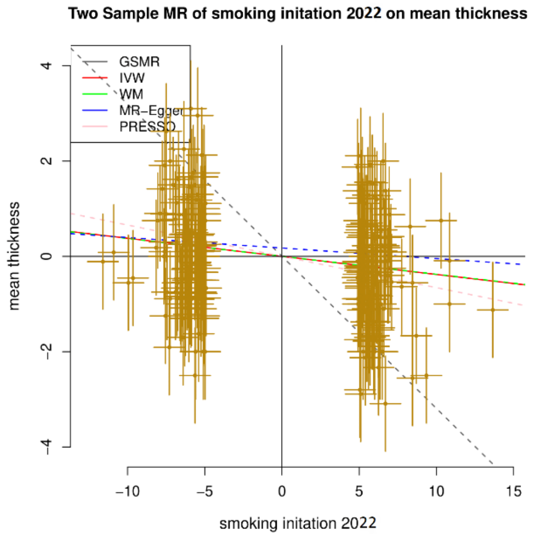

#### E. Global mean TH on BDSCZ

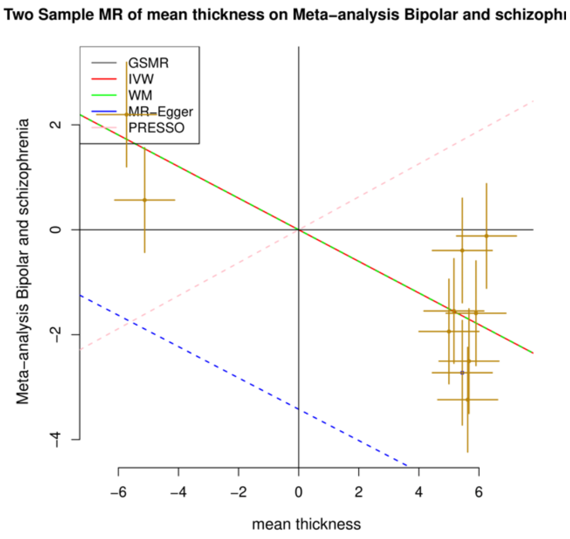

#### BDSCZ on global mean TH

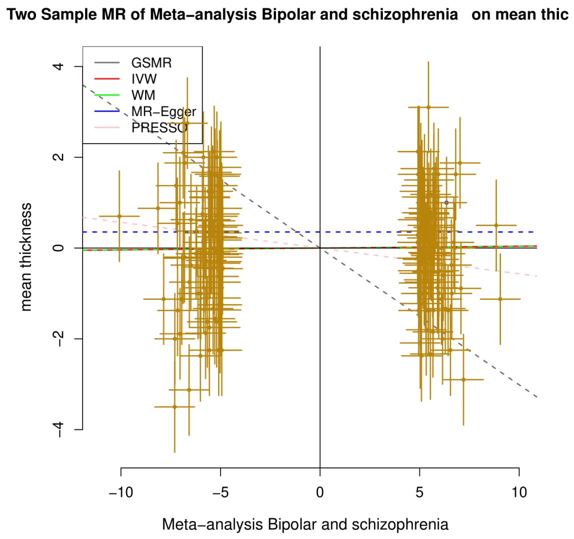

**F. Global mean TH on Bis.DB.ratio** **Bis.DB.ratio on global mean TH**

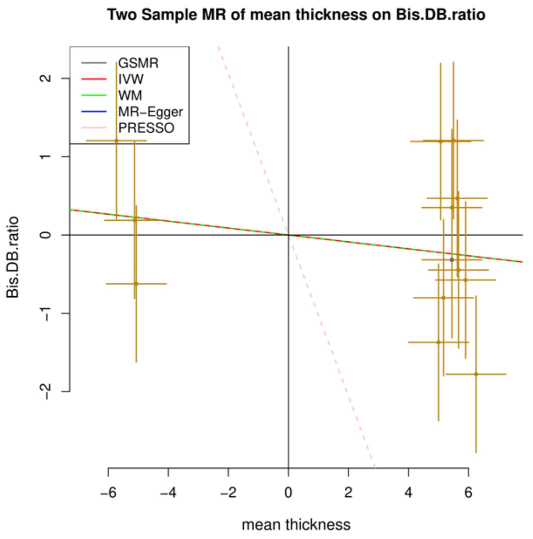

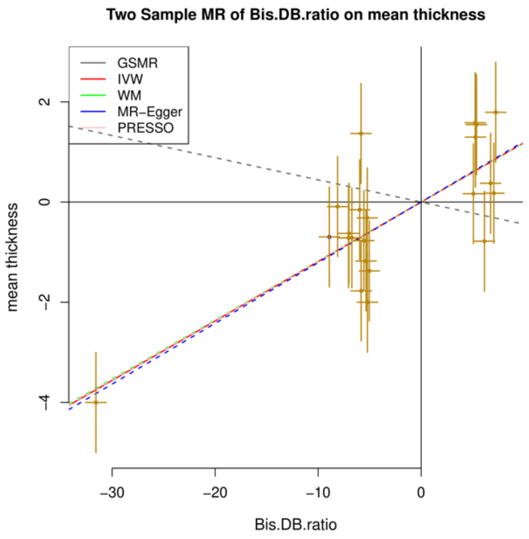

**G. Global mean TH on Bis.FA.ratio** **Bis.FA.ratio on global mean TH**

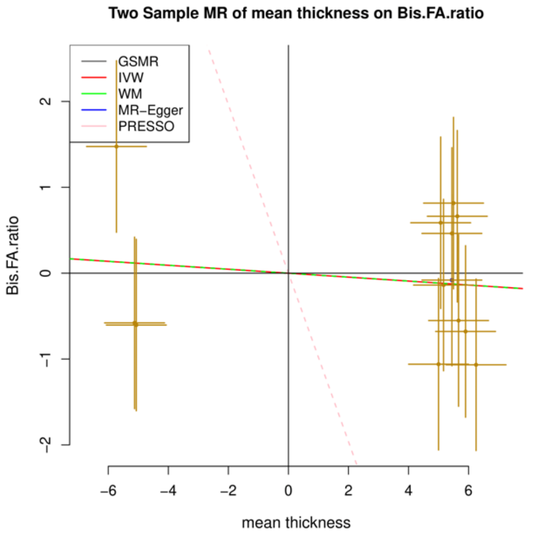

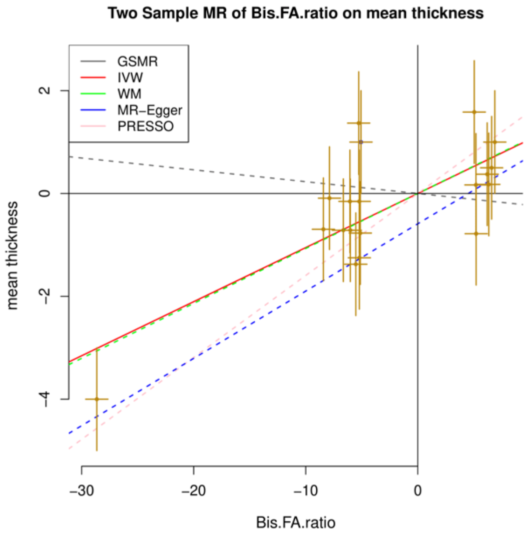

**H. Global mean TH on DB.in.FA** **DB.in.FA on global mean TH**

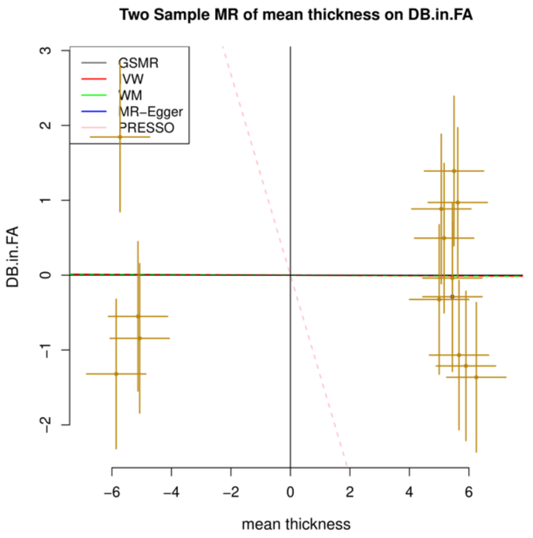

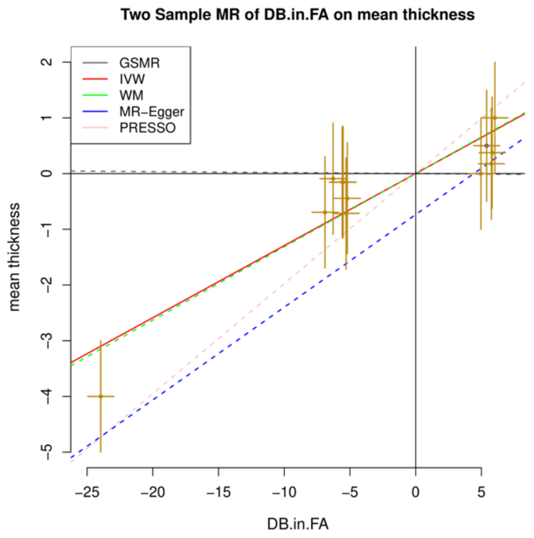

#### I. Global mean TH on CH2.DB.ratio

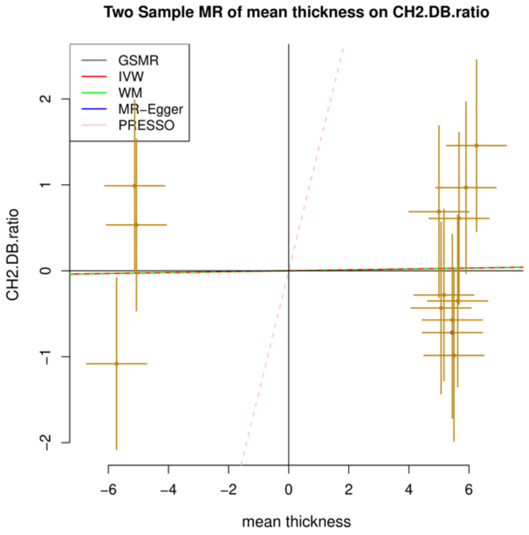

#### CH2.DB.ratio on global mean TH

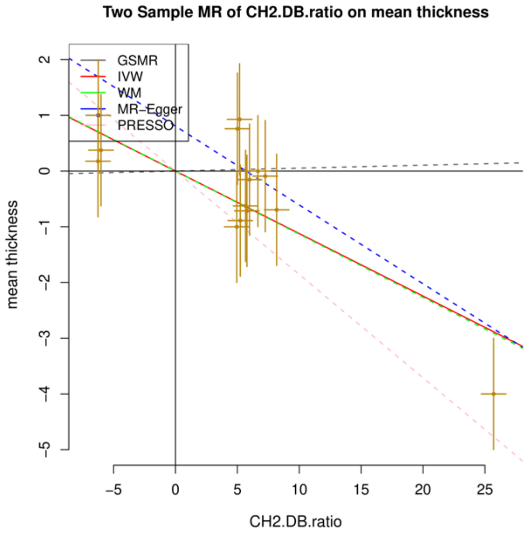

#### J. Global mean TH on CH2.in.FA

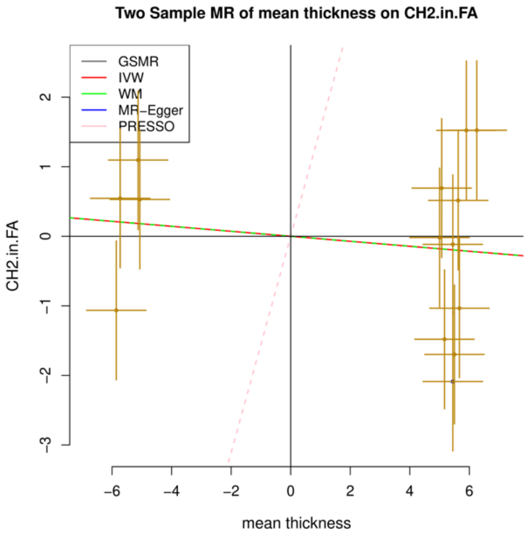

#### CH2.in.FA on mean TH

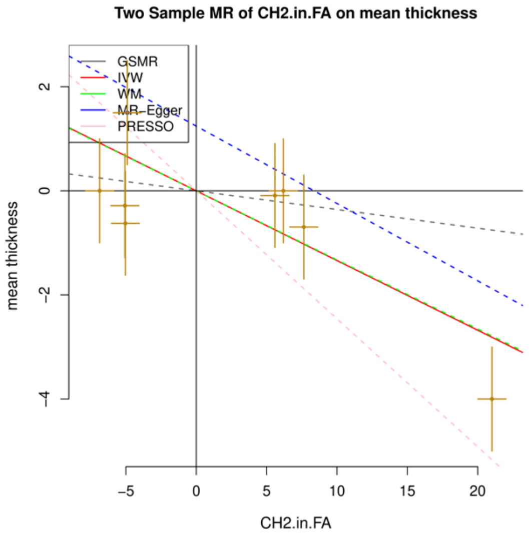

#### K. Global mean TH on Crea

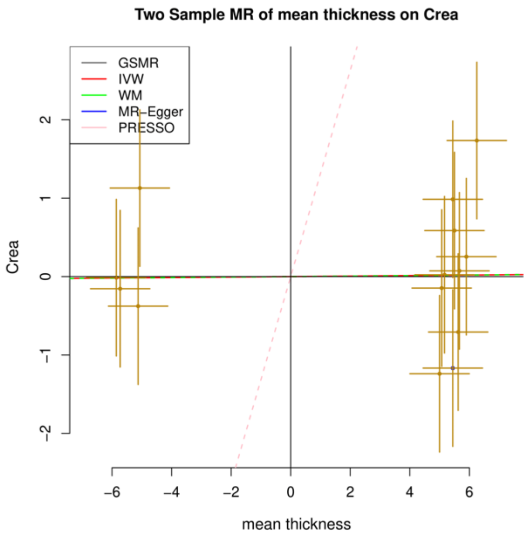

#### Crea on global mean TH

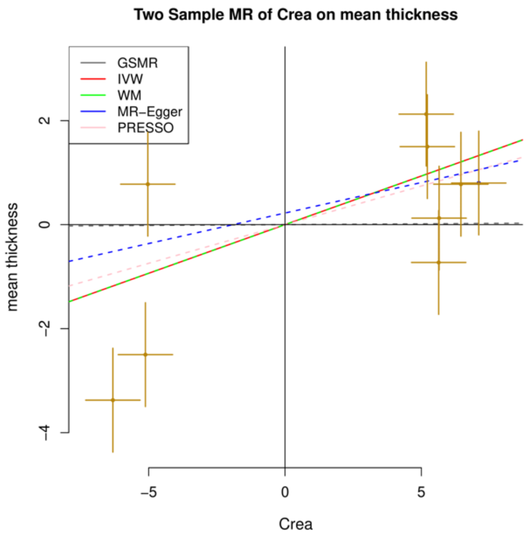

#### L. Global mean TH on otPUFA

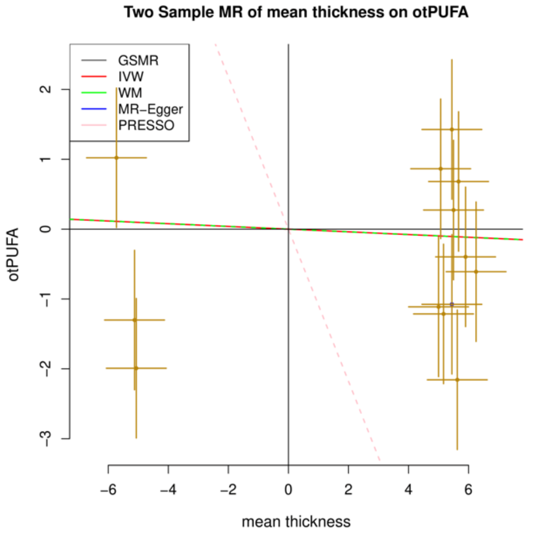

#### otPUFA on global mean TH

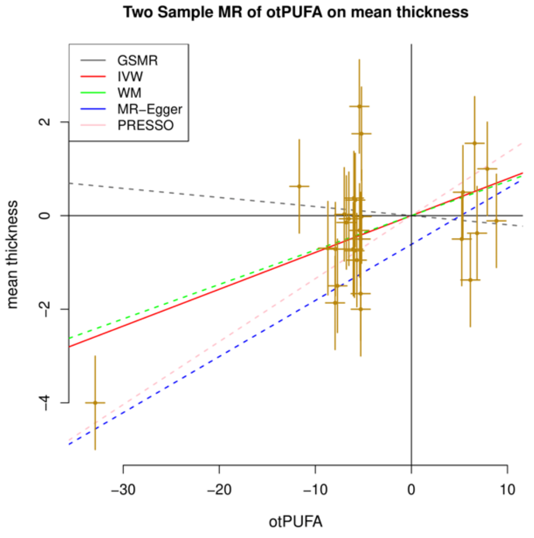

#### M. Global mean TH on BIP

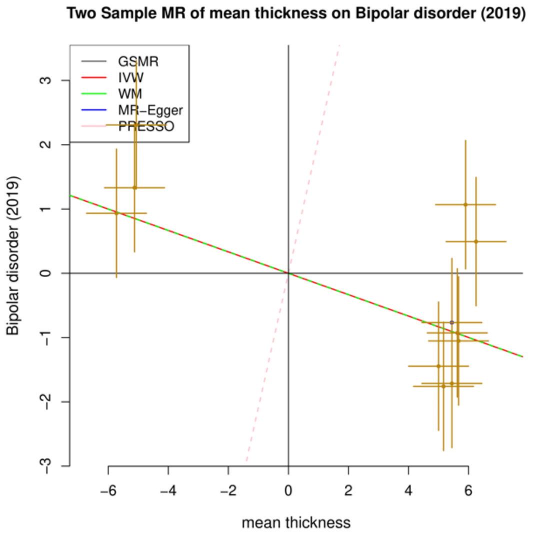

#### BIP on global mean TH

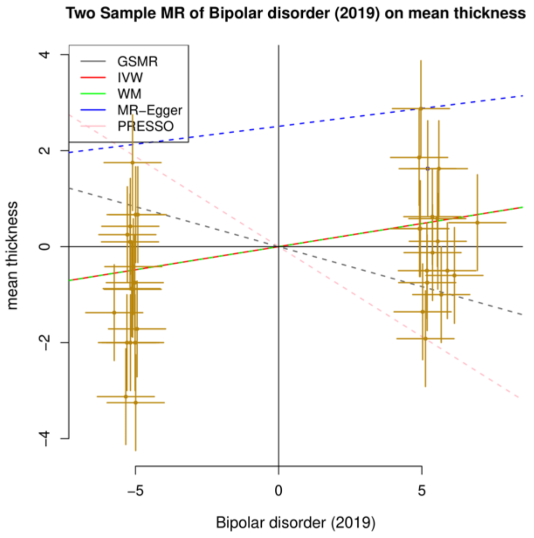

#### N. TH on BMI

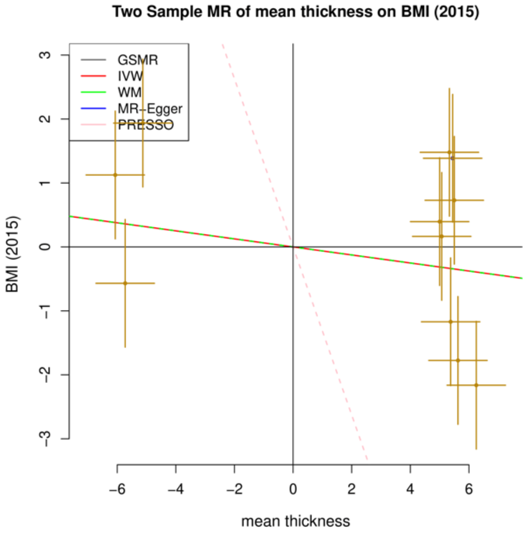

#### BMI on TH

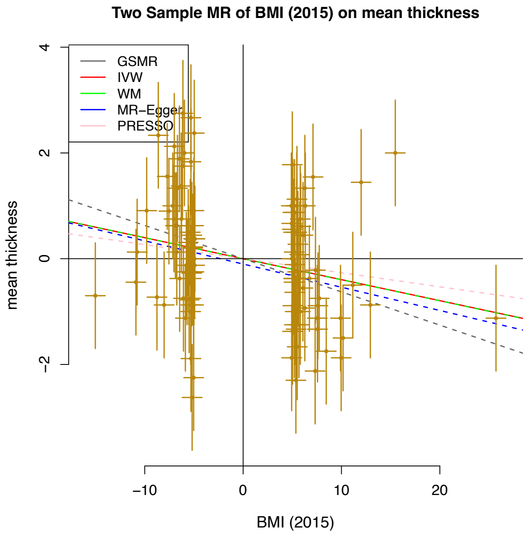

**SFigure 4. Sensitivity Analyses: leave-one-out plots (left) and funnel plots (right) for each significant ($p_{.fdr} < 0.05$) causal association regarding total SA.**

Note that we first show the plots for the forward analyses (total SA on phenotypes) and then the reverse results (phenotypes on total SA). Also note that, for reasons of simplicity and due to spatial constraints, the measure of "total SA" is abbreviated to just "SA" in the title of the leave-one-out plots. More details about the results of the sensitivity analyses can be found in ST6. Regarding the sample size of GWASs, TSA, MTH= 51,665; CP=257,828; EA= 3 million, Height=253,288, CDG= 438,997; HDL, nonHDL , TC, logTG =1,320,016; BIP#2= 413,466; AgeSmk, SmkInit = 1.2 million; T2D2020=23,326; T2D2017=659,316; ALZ=200,853; SmkCes#2, SmkInit#2, DrnkWk#2= 3.4 million; ADHD= 53,293. Statistical tests were two-sided, and p-values were FDR-correction adjusted for multiple testing.

-***LEAVE-ONE-OUT PLOTS*** (left) are a useful representation of sensitivity analyses. This approach involves systematically removing from the analysis one significant genetic variant at a time and re-estimating the causal effect at each iteration. In each leave-one-out plot, the x axis represents the estimates of the causal effect, while the y-axis depicts which variants are excluded each time. The red line at the bottom indicates the causal estimate (along with the confidence intervals) if all significant SNPs are included. Note: in some cases (e.g., Panel T), the y-axis is not clear because of the very large number of genetic variants (the number of genetic variants for each exposure-outcome pair can be found in ST6). Statistical tests were two-sided.
If, by removing a certain genetic variant, we observe a very large difference in the estimated causal effect, this suggests that the specific variant has a strong influence on the results. This is a warning sign for the stability and robustness of the results, since it can be attributed to various problems, mainly overreliance on a single variant, violation of instrumental variables' assumptions and heterogeneity among instruments.
Luckily, our analyses do not identify any particularly large discrepancies.

-***FUNNEL PLOTS*** (right) are also a useful tool to help identify any potential issues in the MR analyses (publication bias, heterogeneity etc.). In each plot and for each significant genetic variant that was used as an instrumental variable, the x-axis represents the estimated causal effect in the form of a beta value (β), as calculated by Instrumental Variable (IV) analyses (hence the $\beta_{IV}$ nomenclature). The y-axis depicts the measure of precision, in our case the inverse of the standard error as calculated by IV ($1/SE_{IV}$). In an ideal scenario, all the points on the plot would be scattered symmetrically around a symmetrically centered line, which represents the overall estimated causal effect. Note: asymmetry does not always mean bias or error. Also, please note that here we include two lines for the overall causal estimate, one for the Inverse-Variance Weighted Method (IVW) (light blue) and one for MR-Egger (darker blue). If the two lines are similar and overlap closely (e.g Panel S), it suggests that both methods yield consistent estimates of the causal effect, providing more confidence in the results.

#### ***Forward analyses***

##### A. Total SA on CP

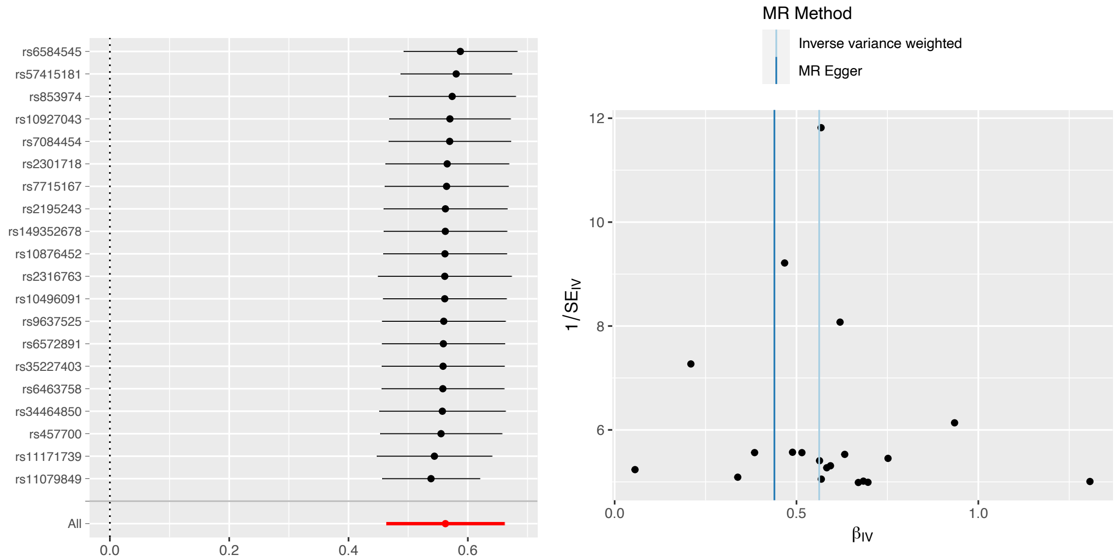

##### B. Total SA on EA

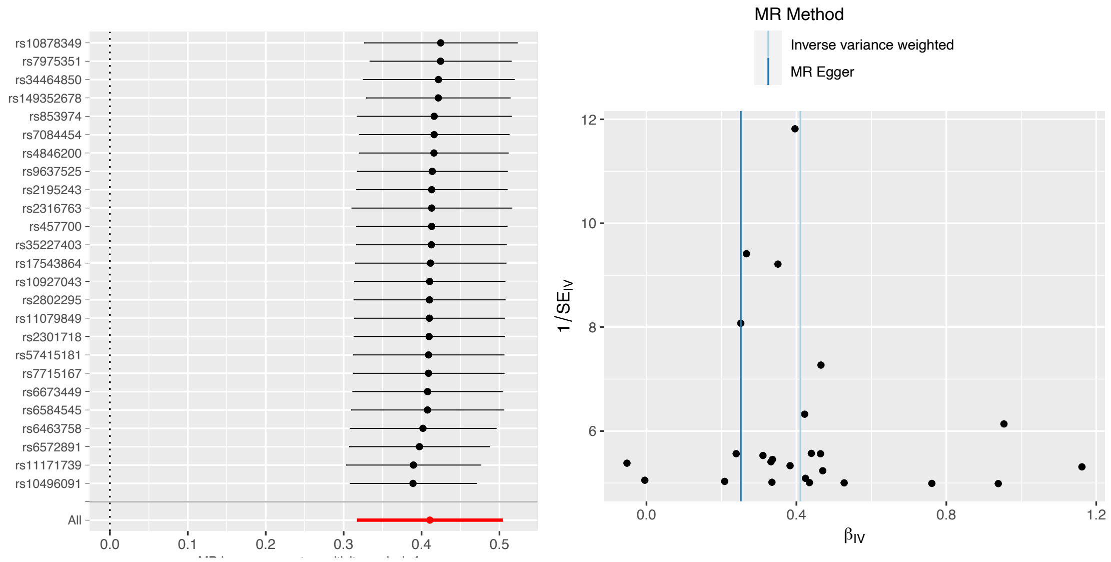

### C. Total SA on Height

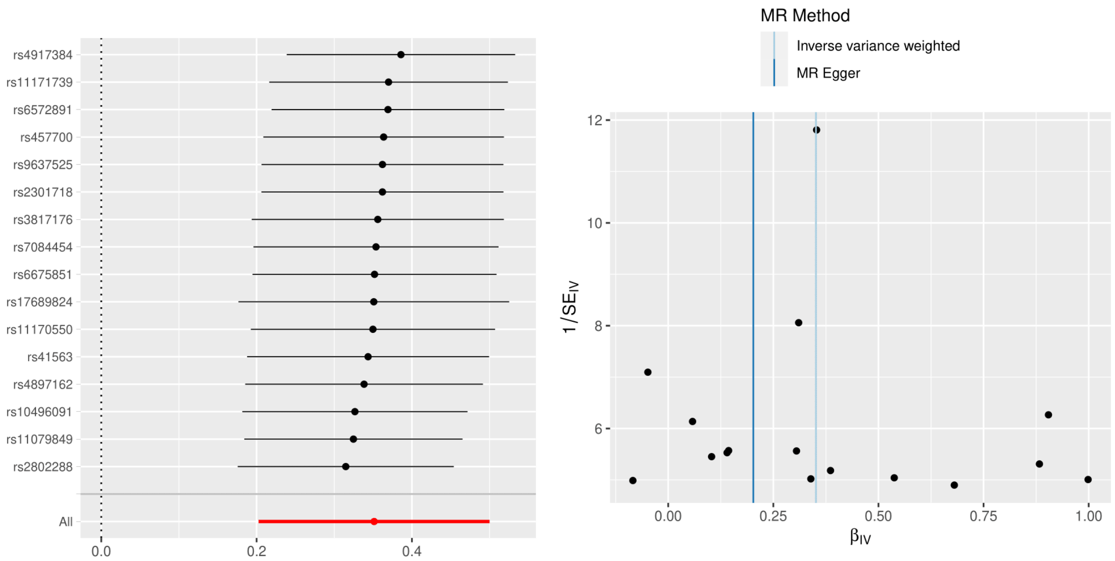

### D. Total SA on logTG

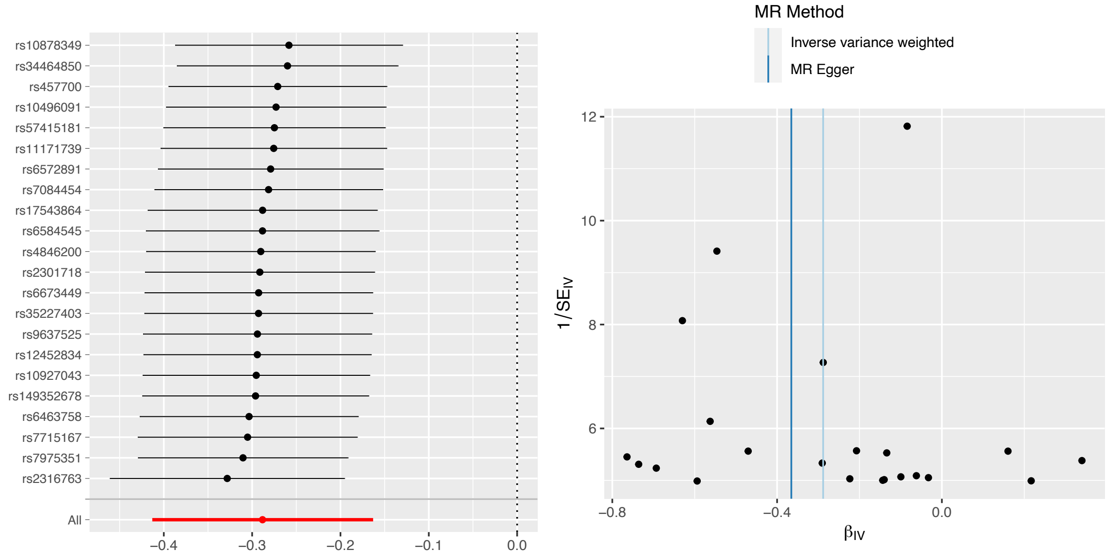

### E. Total SA on SmkInit#2

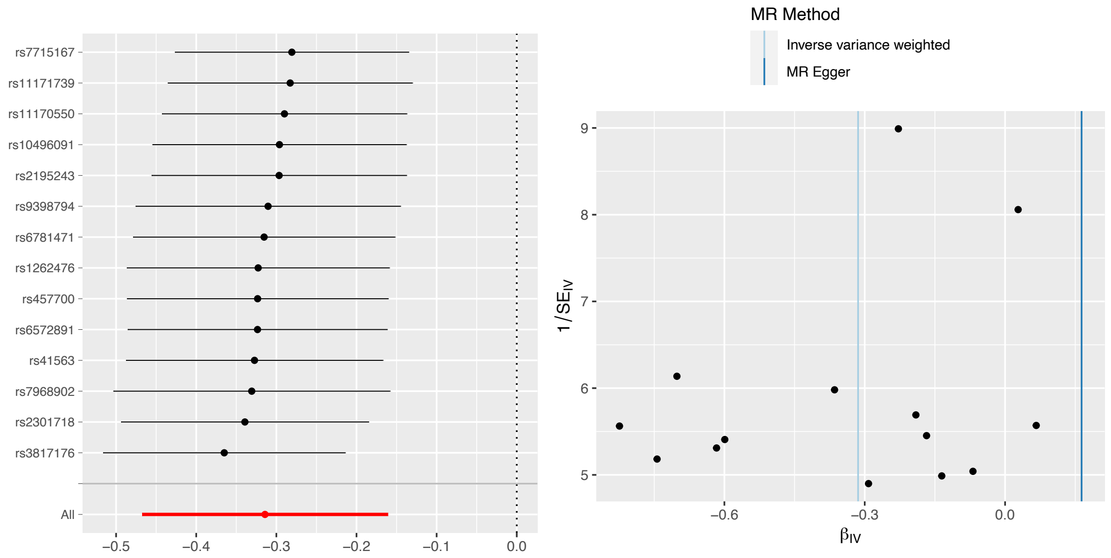

### F. Total SA on CDG

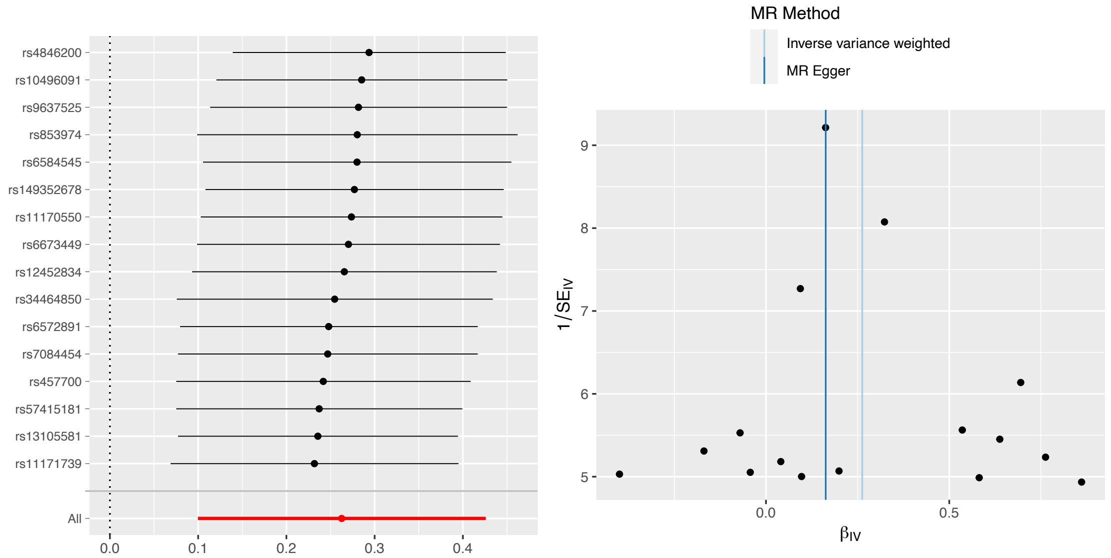

### G. Total SA on ADHD

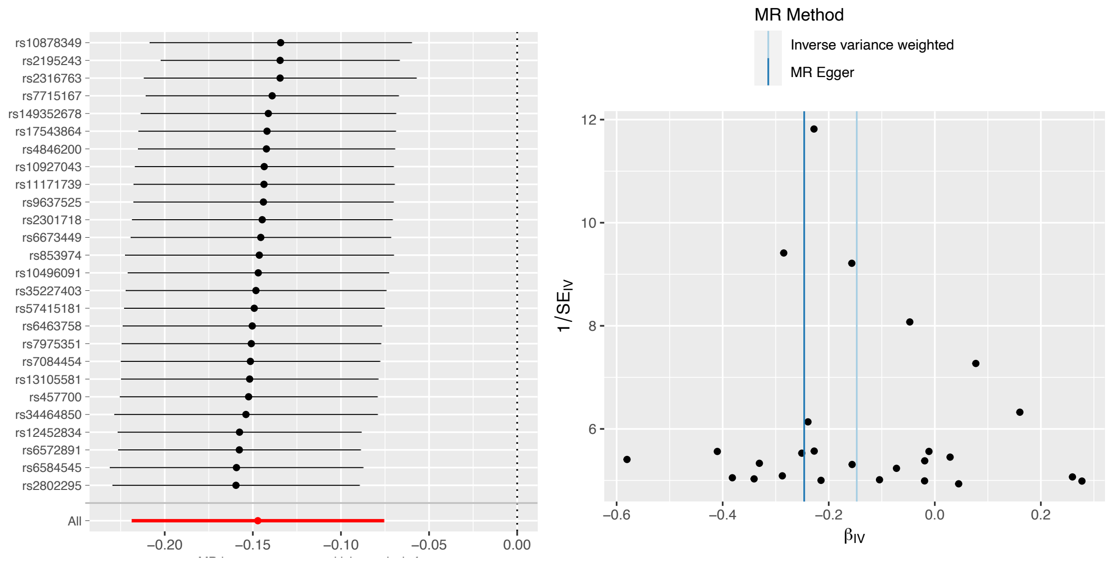

### H. Total SA on AgeSmk#2

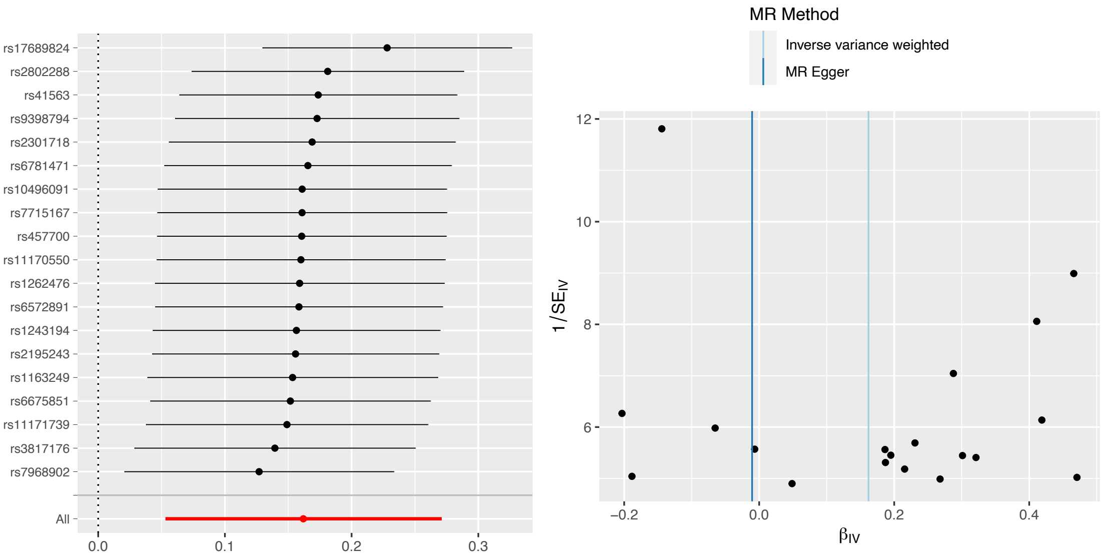

### I. Total SA on HDL

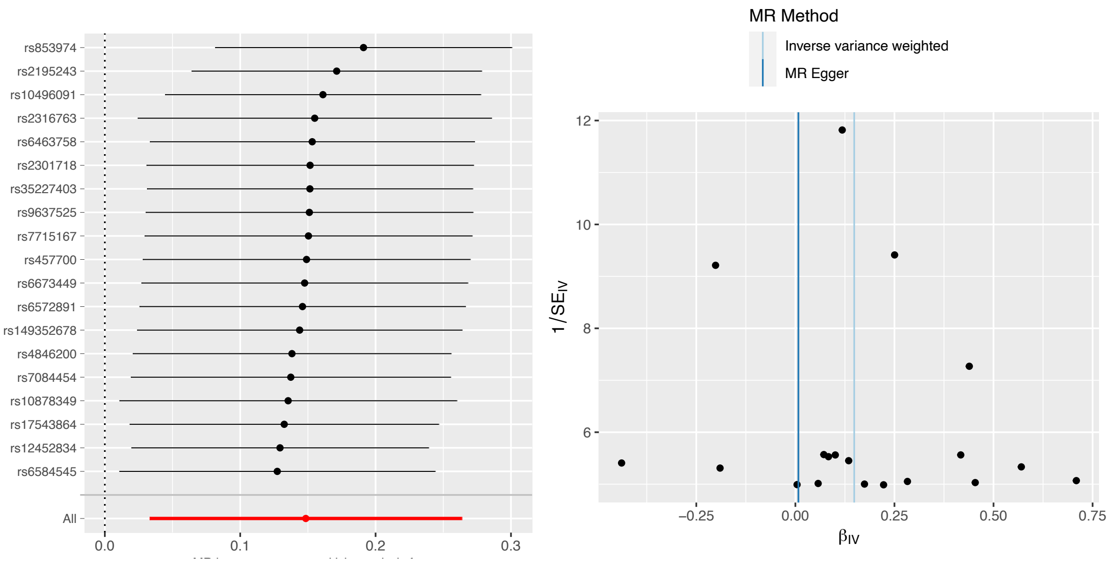

### J. Total SA on SmkCes#2

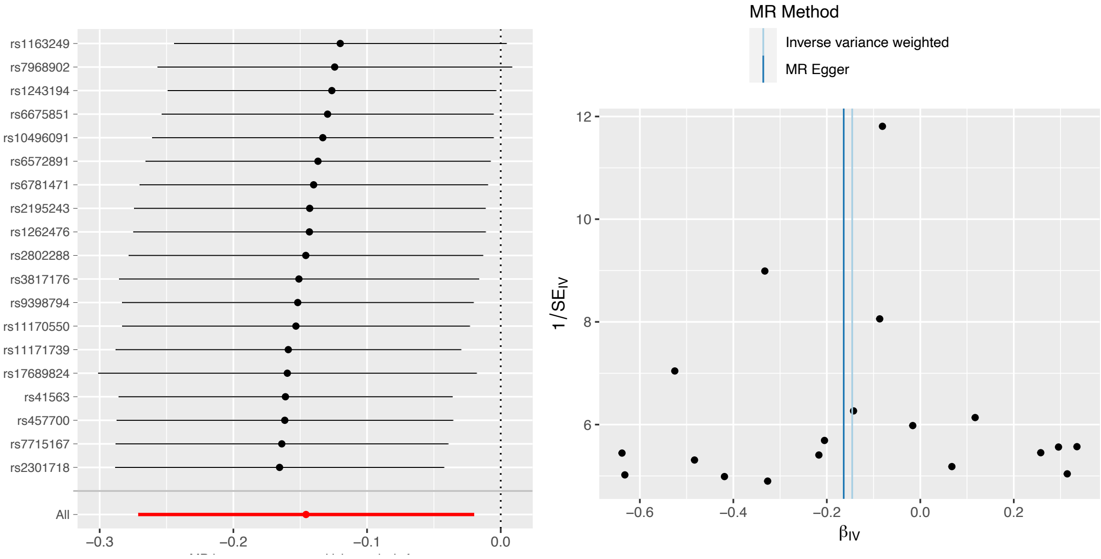

### K. Total SA on non-HDL

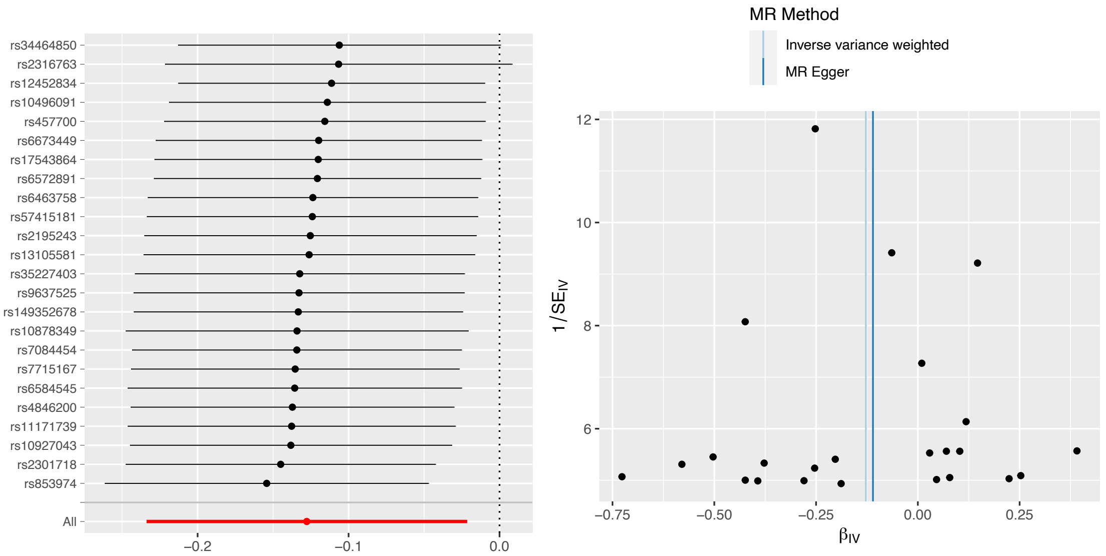

#### L. Total SA on BIP#2

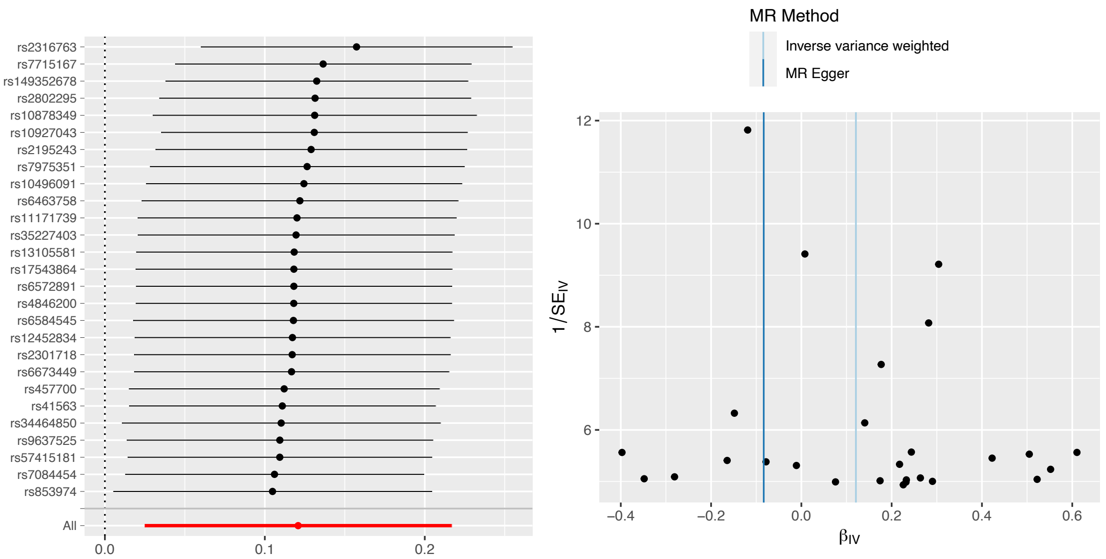

#### M. Total SA on TC

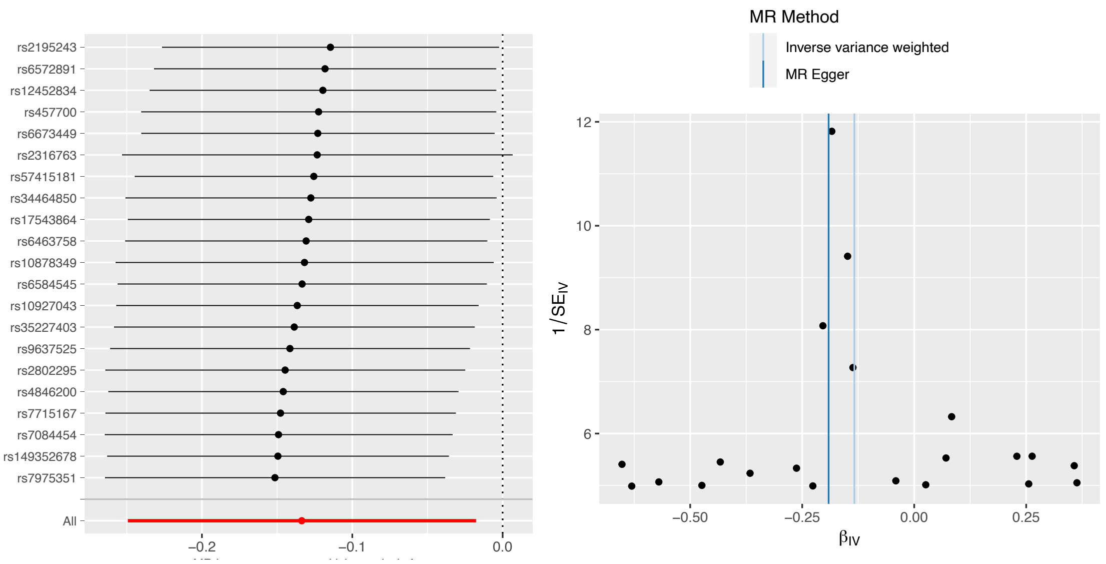

#### N. Total SA on AgeSmk

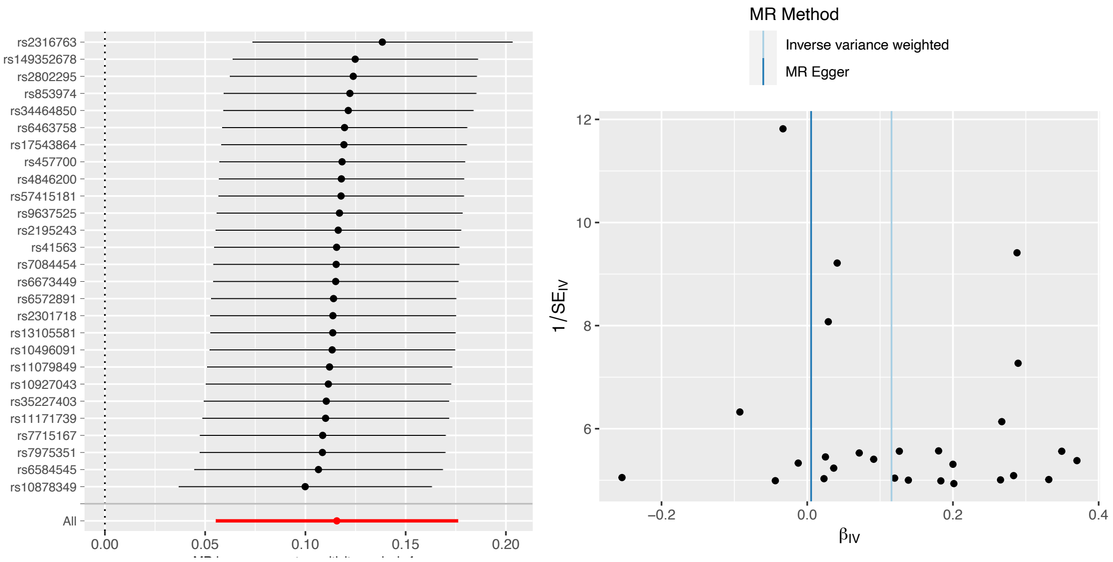

#### O. Total SA on ALZ

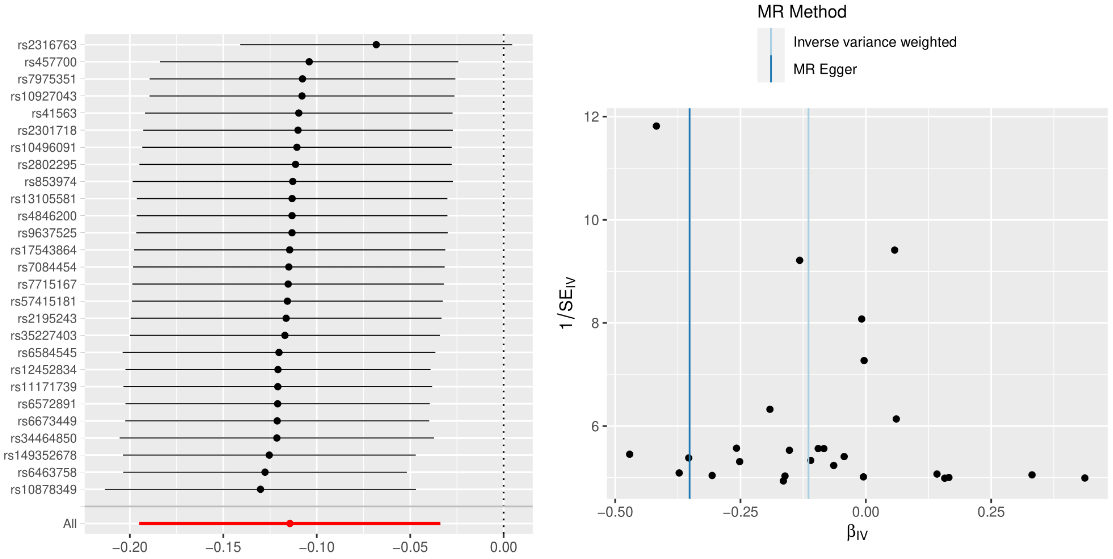

#### P. Total SA on T2D2020

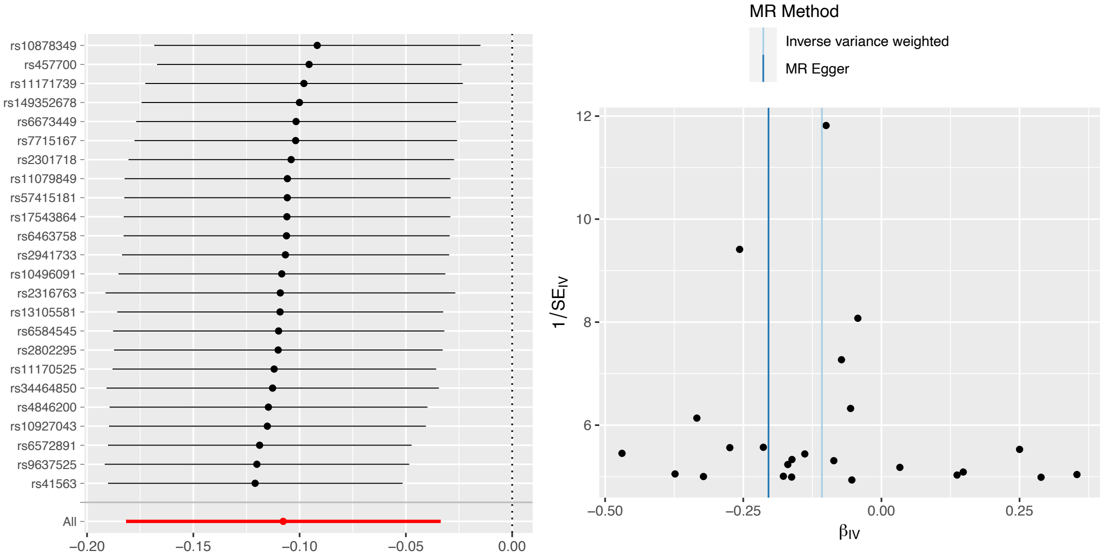

#### Q. Total SA on T2D 2017

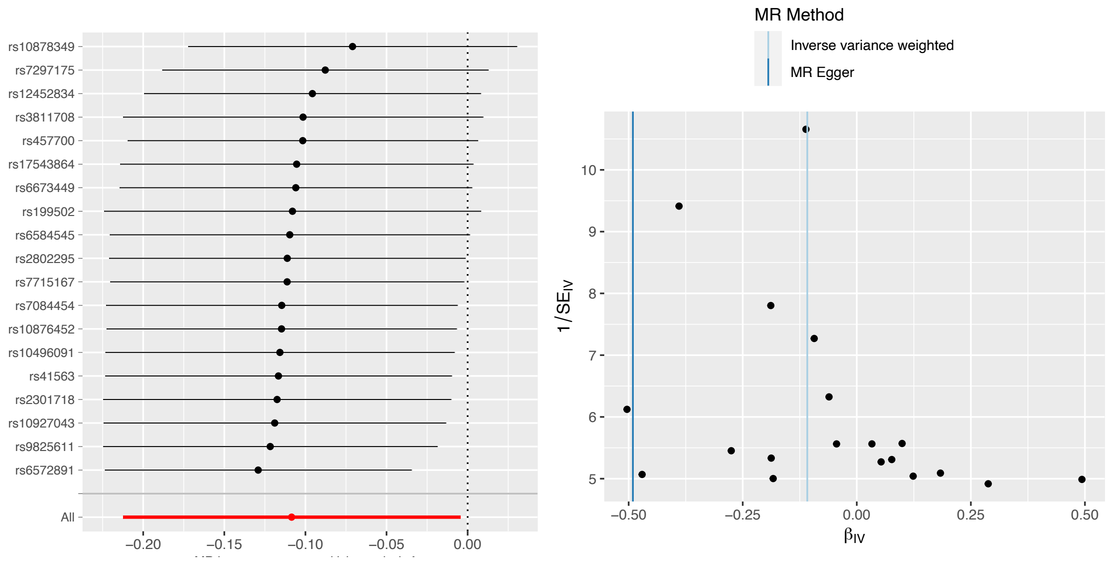

#### ***Reverse analyses***

#### R. Height on total SA

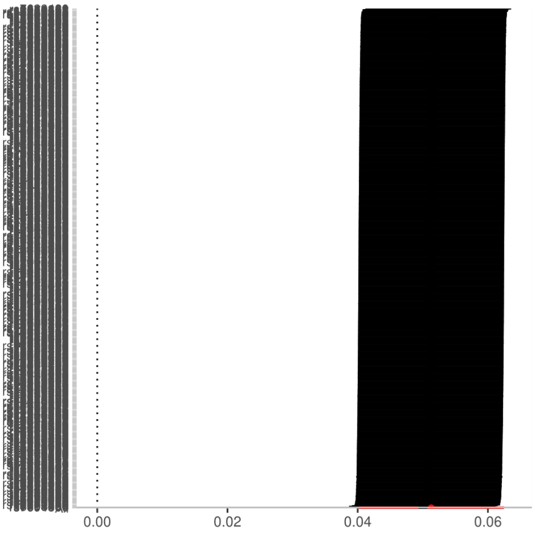

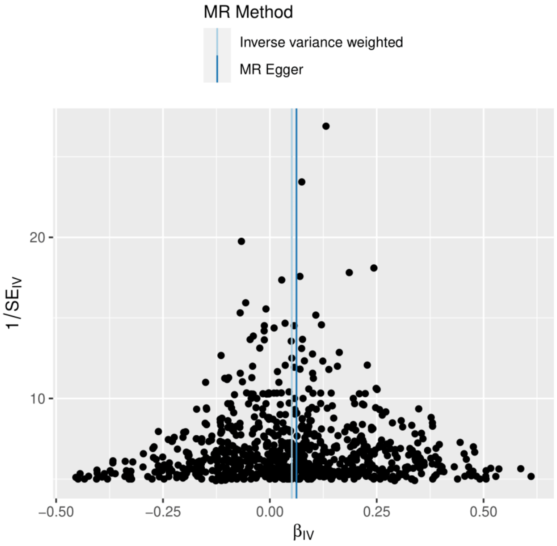

#### S. CP on total SA

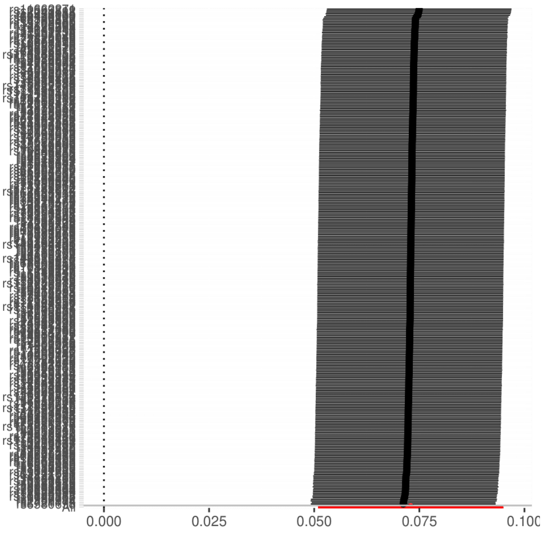

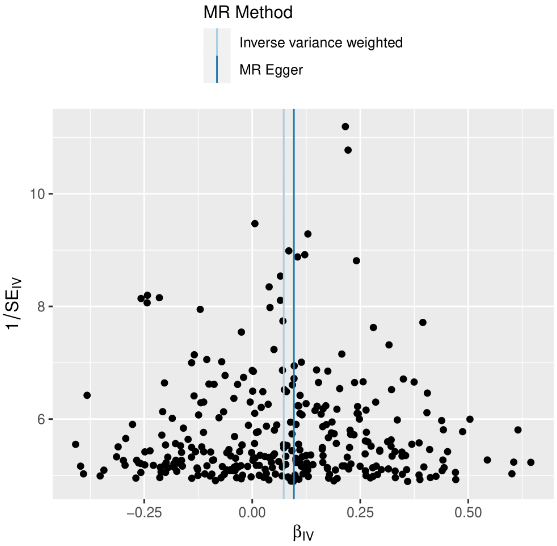

#### SFigure 5. Sensitivity Analyses: leave-one-out plots (left) and funnel plots (right) for each significant ($p_{fdr} < 0.05$) causal association regarding global mean TH.

Note that we first show the plots for the forward analyses (global mean TH on phenotypes) and then the reverse results (phenotypes on global mean TH). Also note that, for reasons of simplicity and due to spatial constraints, the measure of "global mean TH" is abbreviated to just "TH". More details about the results of the sensitivity analyses can be found in ST7. Regarding the sample size of GWASs, MTH= 51,665; AgeSmk, SmkInit = 1.2 million; SmkCes#2, SmkInit#2, DrnkWk#2= 3.4 million; SCZ3=161,405; SCZ2=105,308; BDSCZ=41,653; BIP=31,710; BMI=322,154; Bis.DB.ratio, CH2.in.FA, Bis.FA.ratio, otPUFA, DB.in.FA, Crea, CH2.DB.ratio= 24,925. Statistical tests were two-sided, and p-values were FDR-correction adjusted for multiple testing.

-***<u>LEAVE-ONE-OUT PLOTS</u>*** are a useful representation of sensitivity analyses. This approach involves systematically removing from the analysis one significant genetic variant at a time and re-estimating the causal effect at each iteration. In each leave-one-out plot, the x axis represents the estimates of the causal effect, while the y-axis depicts which variants are excluded each time. The dots (centre for the error bars) represent beta coefficients in MR study. The error bars represent 95% Confidence Intervals of beta coefficients. The red line at the bottom indicates the causal estimate (along with the confidence intervals) if all significant SNPs are included. Note: in some cases (e.g., Panel I), the y-axis is not clear because of the very large number of genetic variants (the number of genetic variants for each exposure-outcome pair can be found in ST7). If, by removing a certain genetic variant, we observe a very large difference in the estimated causal effect, this suggests that the specific variant has a strong influence on the results. This is a warning sign for the stability and robustness of the results, since it can be attributed to various problems, mainly overreliance on a single variant, violation of instrumental variables' assumptions and heterogeneity among instruments.
Luckily, our analyses do not identify any particularly large discrepancies. However, it is worth noting that the exclusion of a certain genetic variant (rs174546 in Panels G, H, J and K) seems to consistently lead to a divergent causal effect, even across several (related) phenotypes (Bis.FA.ratio, DB.in.FA, CH2.DB.ratio and CH2.in.FA). Fortunately, in all those cases the overall causal estimate remains stable. This indicates that the particular variant might have a strong influence on the estimated effect but is not essential for the overall inference. This can be attributed to various causes, namely redundancy across instruments, weak instrument or weak association, compensation by other instruments etc. Interestingly, rs174546 is an SNP found in the well-known *FADS1* gene and is highly associated with multiple plasma fatty acid concentrations.[41]
-***<u>FUNNEL PLOTS</u>*** (right) are also a useful tool to help identify any potential issues in the MR analyses (publication bias, heterogeneity etc.). In each plot and for each significant genetic variant that was used as an instrumental variable, the x-axis represents the estimated causal effect in the form of a beta value (β), as calculated by Instrumental Variable (IV) analyses (hence the $\beta_{IV}$ nomenclature). The y-axis depicts the measure of precision, in our case the inverse of the standard error as calculated by IV ($1/SE_{IV}$). In an ideal scenario, all the points on the plot would be scattered symmetrically around a symmetrically centered line, which represents the overall estimated causal effect. Note: asymmetry does not always mean bias or error. Also, please note that here we include two lines for the overall causal estimate, one for the Inverse-Variance Weighted Method (IVW) (light blue) and one for MR-Egger (darker blue). If the two lines are similar and overlap closely (e.g., Panel F), it suggests that both methods yield consistent estimates of the causal effect, providing more confidence in the results.

#### ***<u>Forward analyses</u>***

##### A. Global mean TH on SCZ3

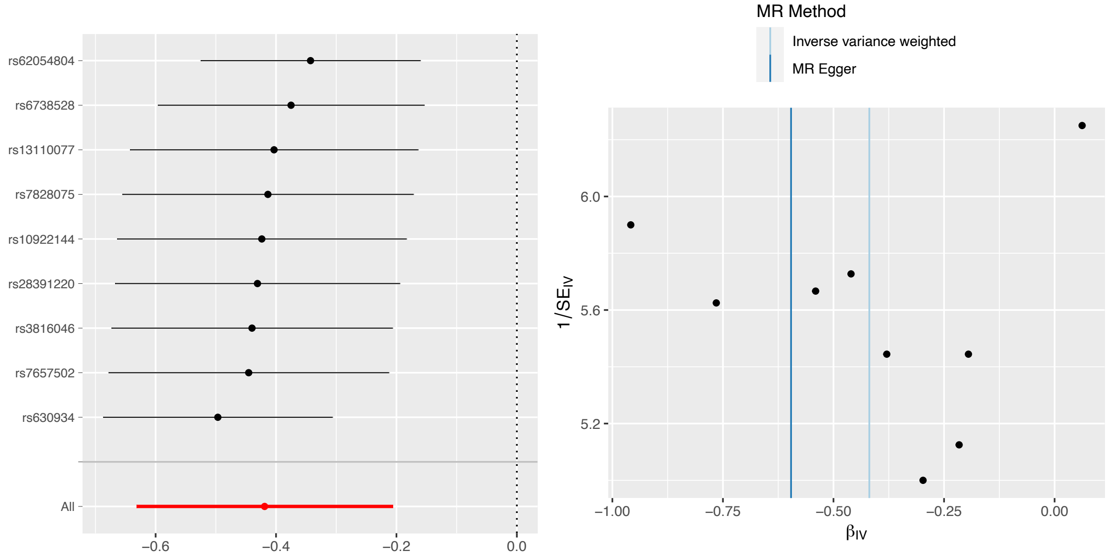

#### B. Global mean TH on SCZ2

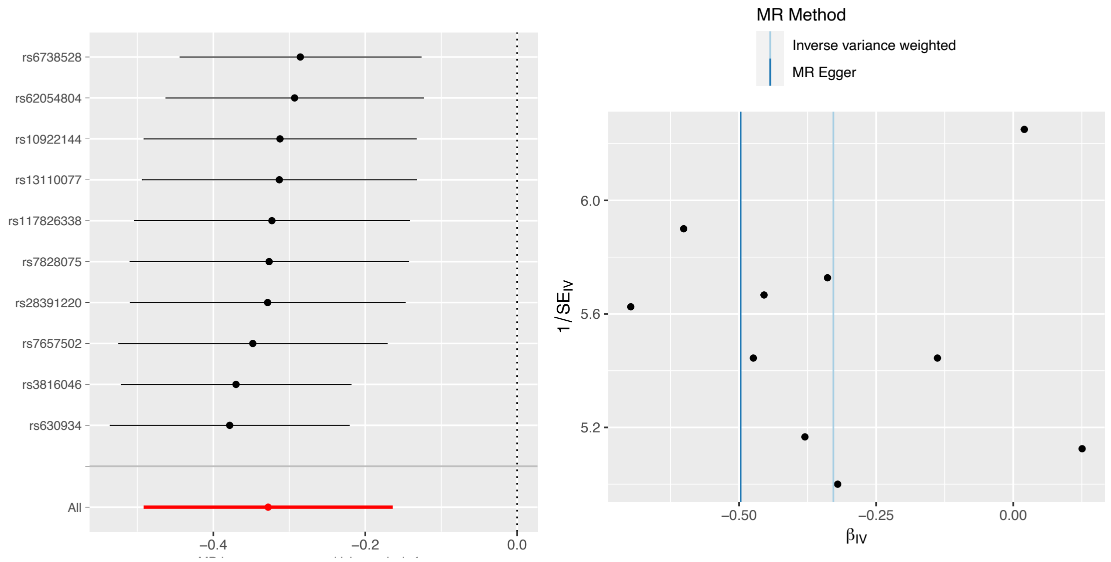

#### C. Global mean TH on DrnkWk#2

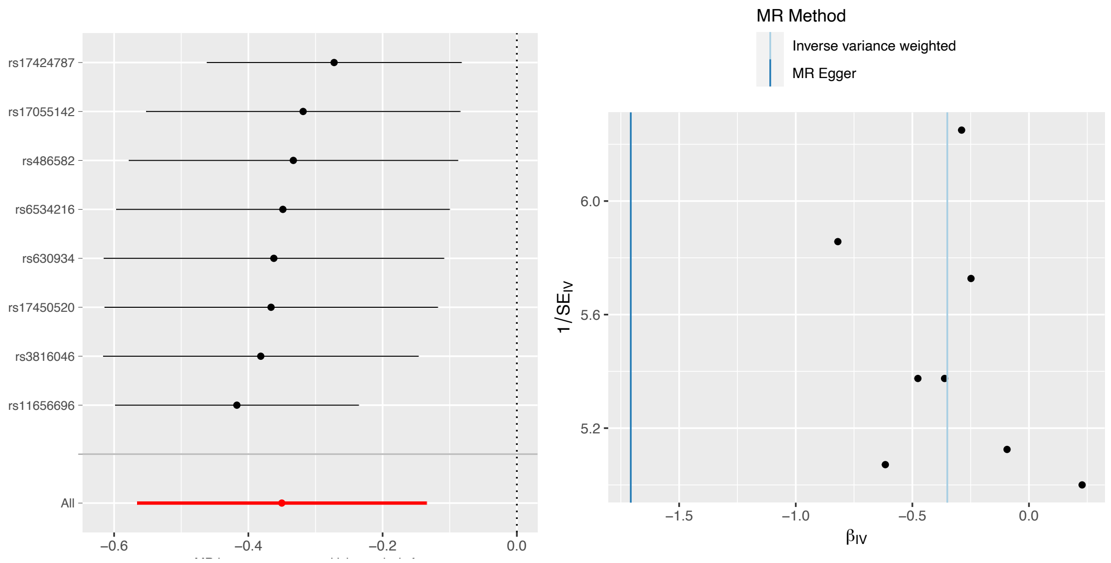

#### D. Global mean TH on SmkInit#2

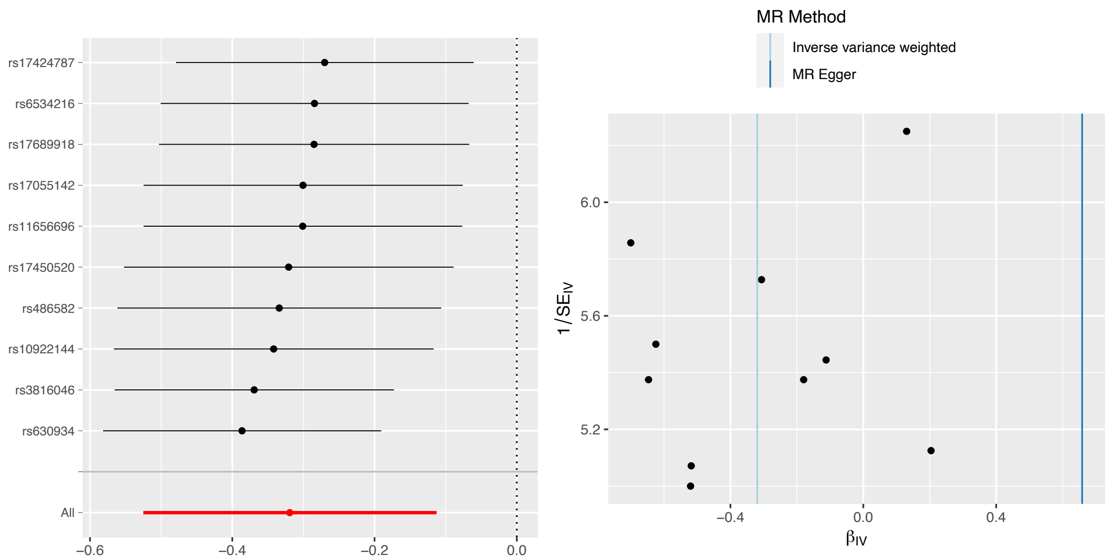

#### E. Global mean TH on BDSCZ

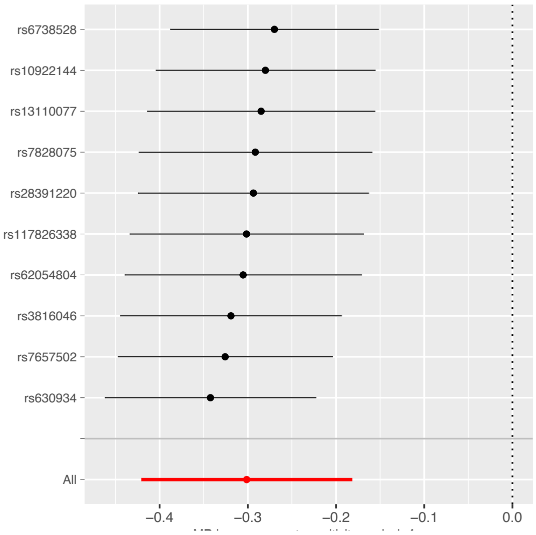

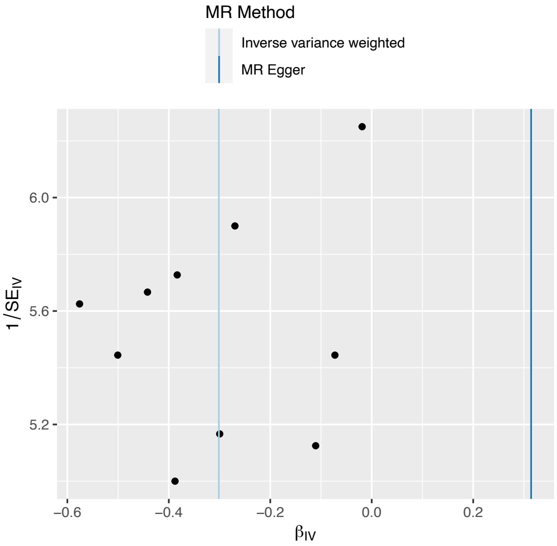

### *Reverse analyses*

#### F. Bis.DB.ratio on global mean TH

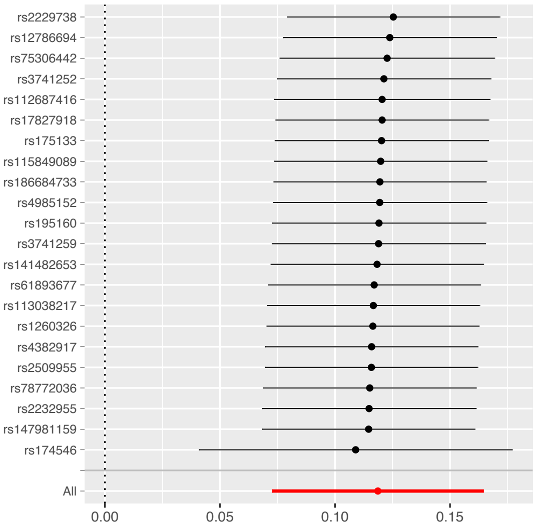

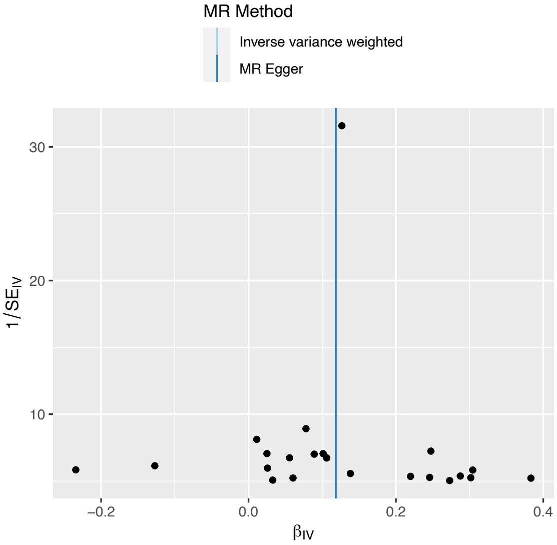

#### G. Bis.FA.ratio on global mean TH

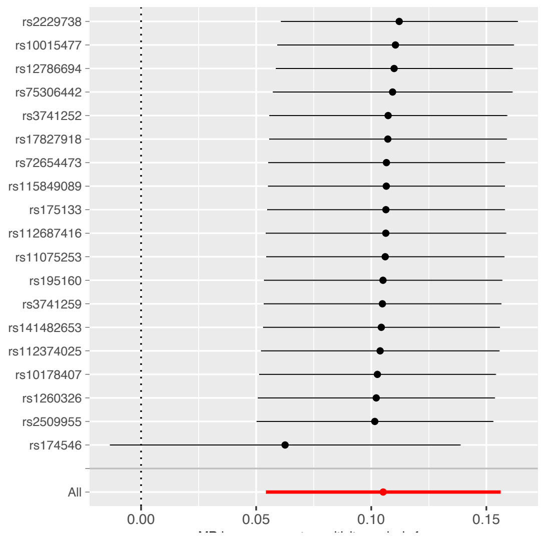

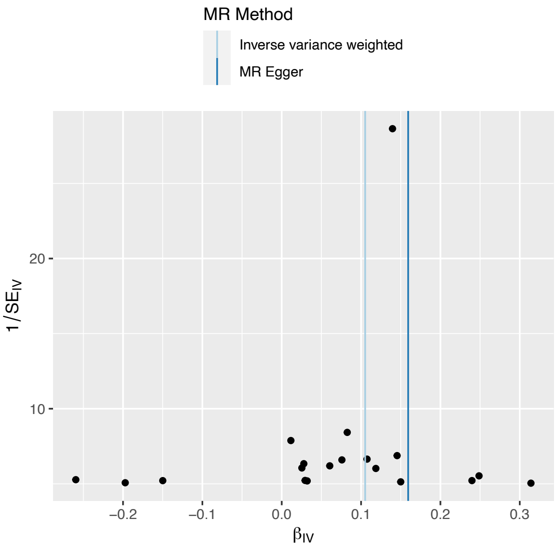

#### H. DB.in.FA on global mean TH

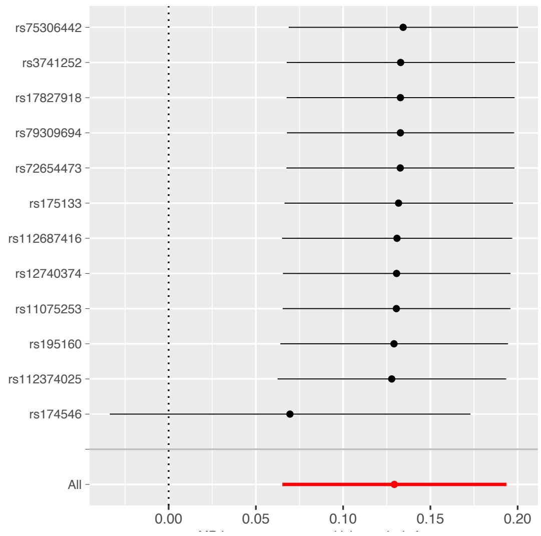

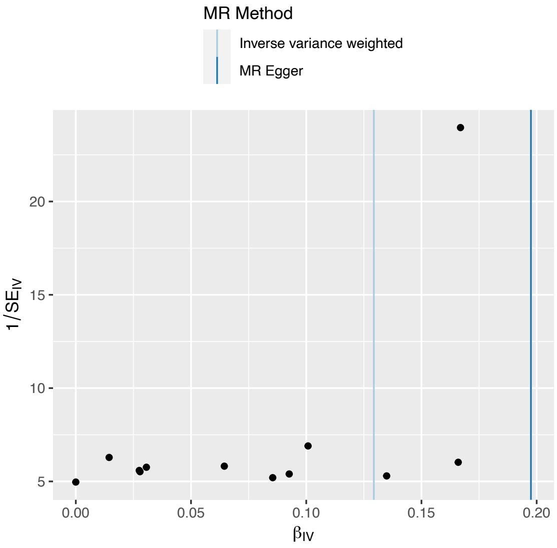

#### I. SmkInit#2 on global mean TH

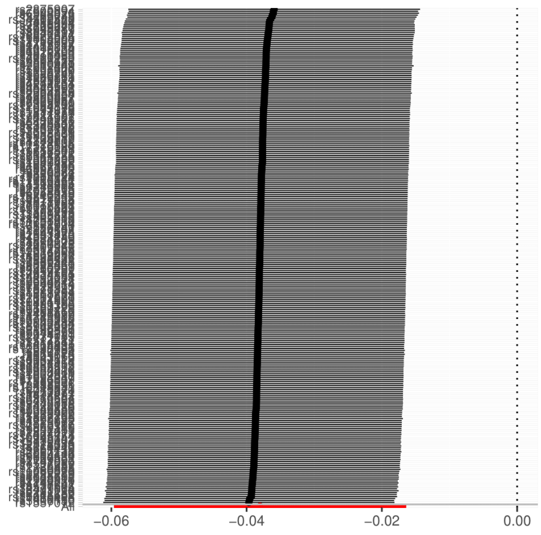

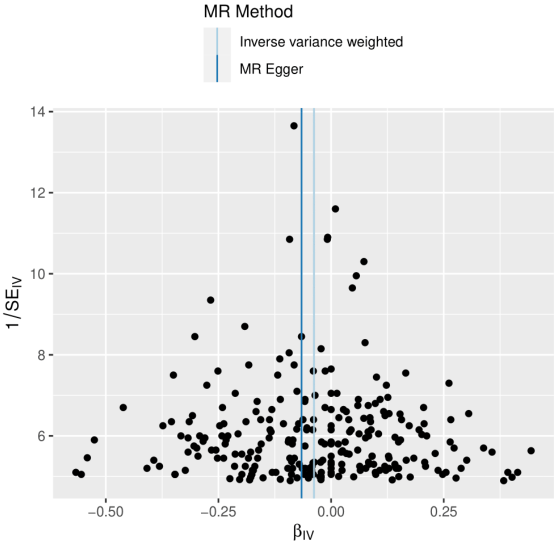

#### J. CH2.DB.ratio on global mean TH

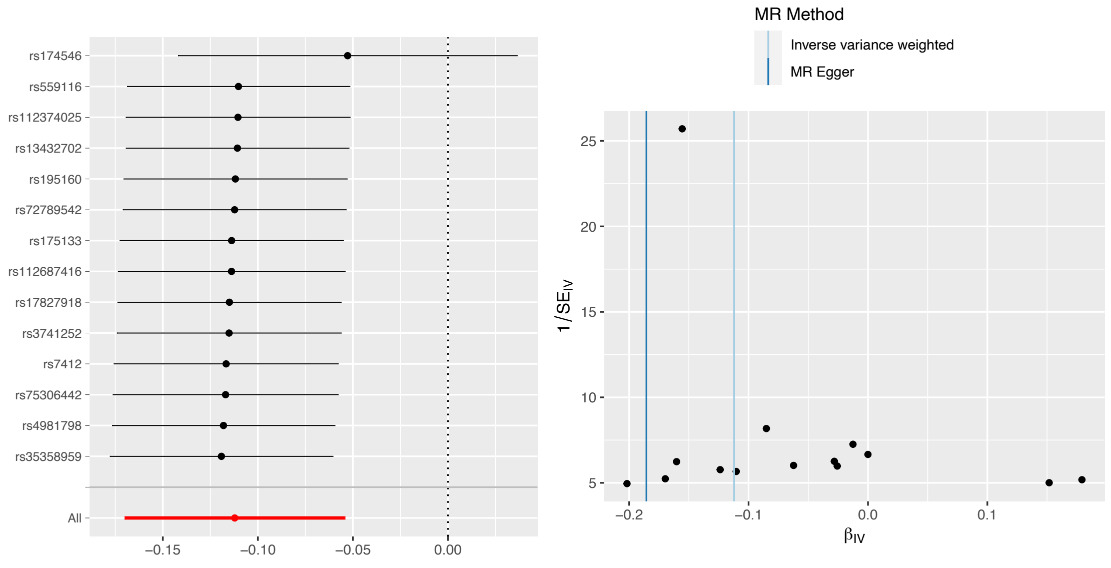

#### K. CH2.in.FA on global mean TH

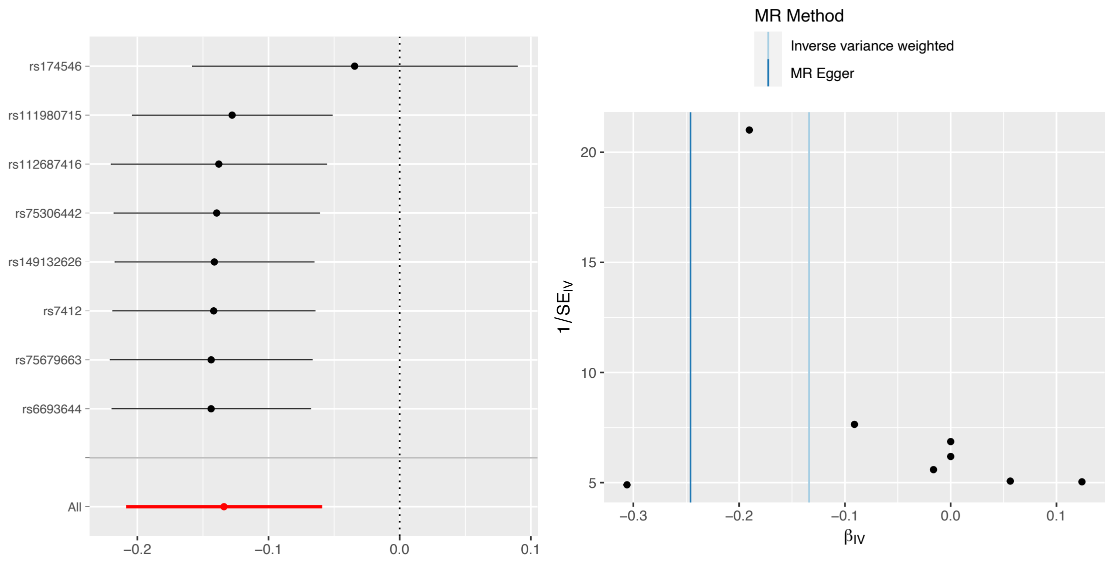

#### L. Crea on global mean TH

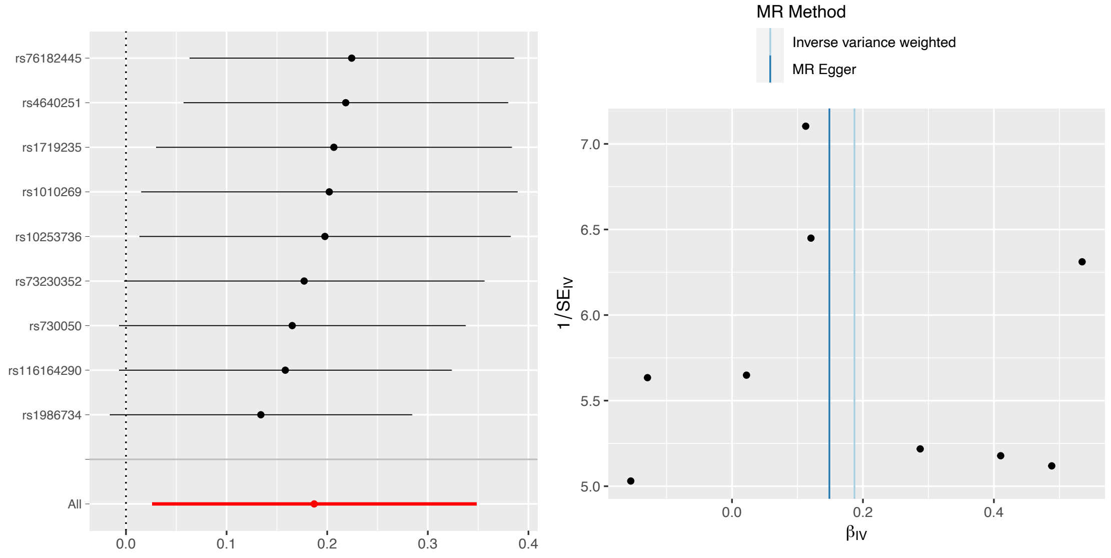

#### M. OtPUFA on global mean TH

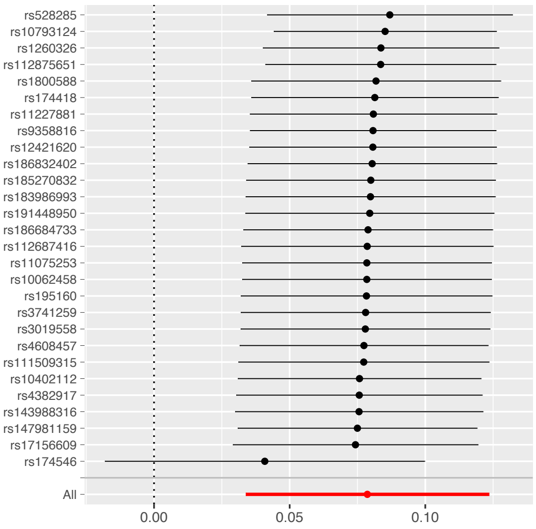

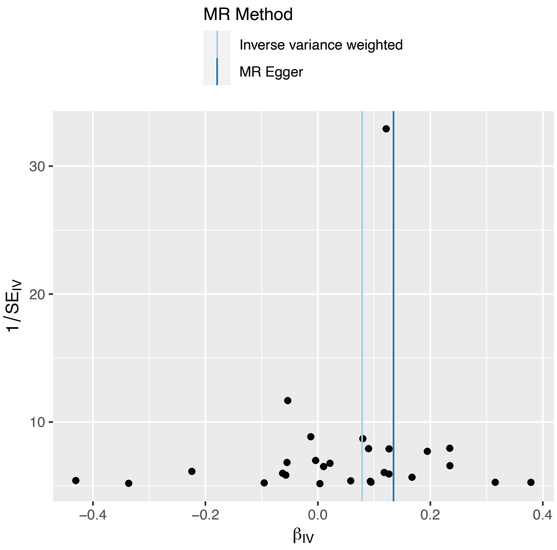

#### N. BIP on global mean TH

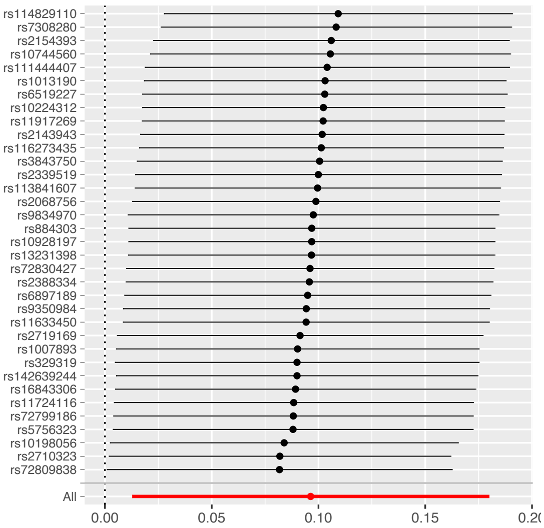

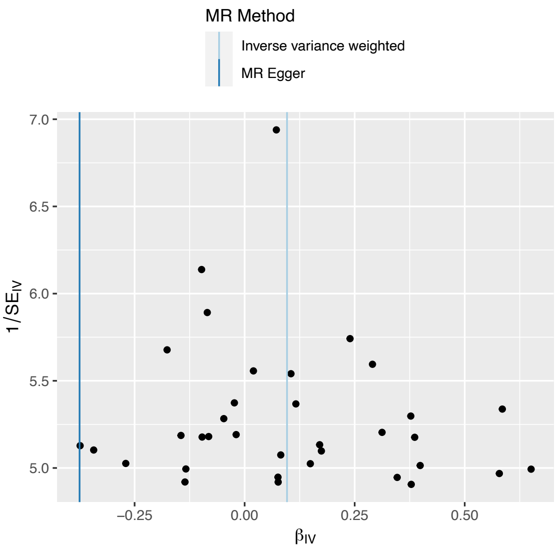

#### O. BMI on global mean TH

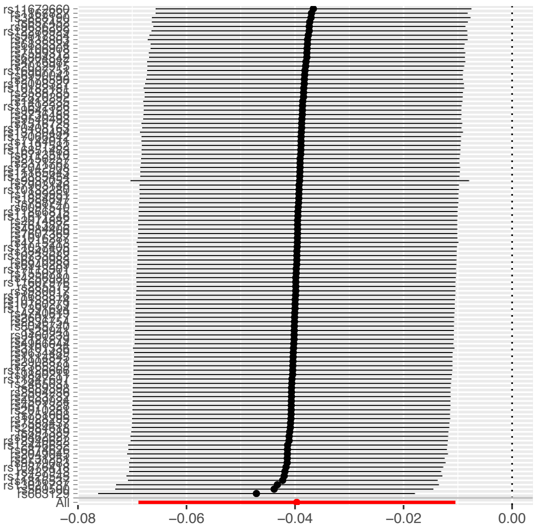

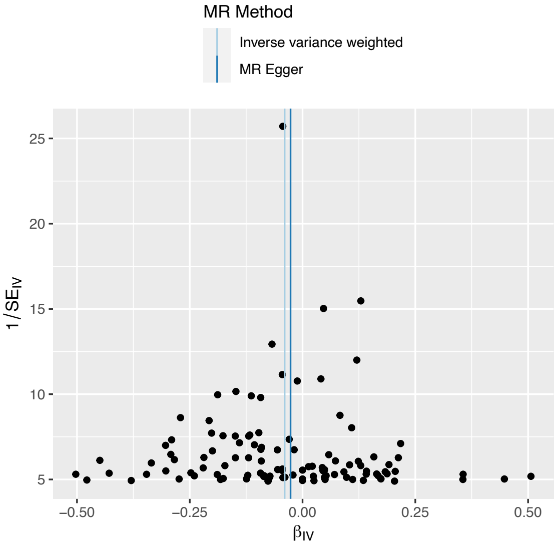

**Figure 6. Regional plots for surface area (SA) with top causality results, as determined from the general GSMR analyses (forward and reverse).**

Global corrected (left) and non-global-corrected (right) MR results between SA regional measures and the significant results, as determined by the forward and reverse analyses between total SA and the phenotypes (see Supplementary Data ST2, ST3). In panels A and C, the color intensity represents the strength of the causal association via beta coefficients (red: strong positive, blue: strong negative). Panels B and D show the -log10 p-value after FDR-correction: the higher this value (i.e., the lighter the blue), the more statistically significant the result. Visualization with the use of the "ggseg"[42] R package in a plot of the Desikan-Killiany atlas, right hemisphere (upper) and left hemisphere (lower). Lateral and medial sections of the brain are also depicted. Statistical tests were two-sided, and p-values were FDR-correction adjusted for multiple testing. Regarding the sample size of GWASs, TSA= 51,665; CP=257,828; EA= 3 million, Height=253,288, CDG= 438,997; HDL, nonHDL , TC, logTG =1,320,016;  BIP#2= 413,466; AgeSmk, SmkInit = 1.2 million;  T2D2020=23,326; T2D2017=659,316; ALZ=200,853; SmkCes#2, SmkInit#2, DrnkWk#2= 3.4 million; ADHD= 53,293.

A. Beta coefficients of MR results of regional SA on cognitive performance (CP).

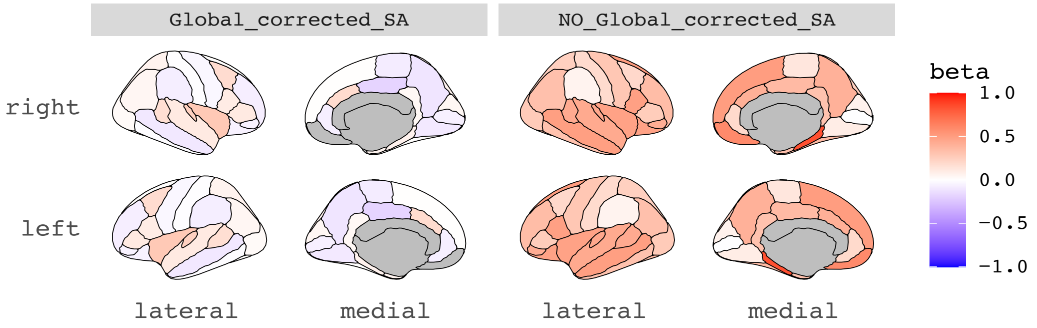

B. -log10 ($p_{fdr}$ values) of MR results of regional SA on cognitive performance (CP).

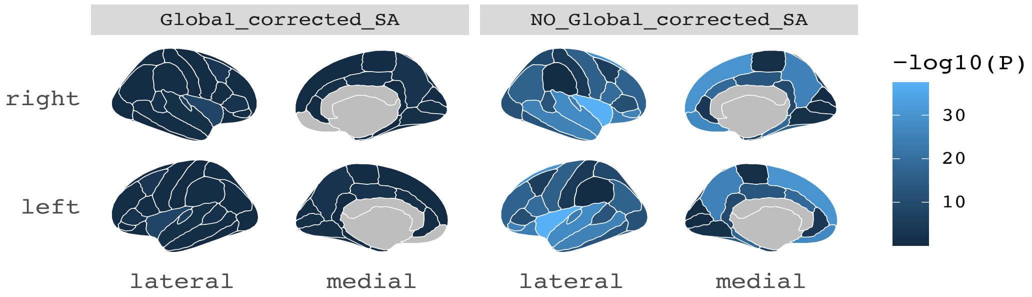

C. Beta coefficients of MR results of cognitive performance (CP) on regional SA.

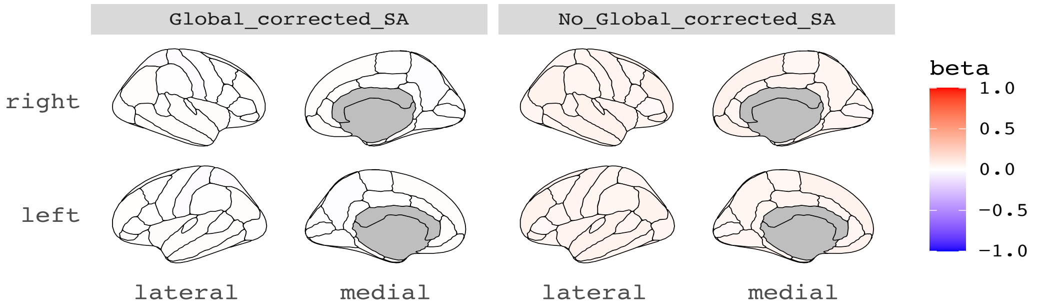

D. -log10 ($p_{fdr}$ values) of MR results of cognitive performance (CP) on regional SA.

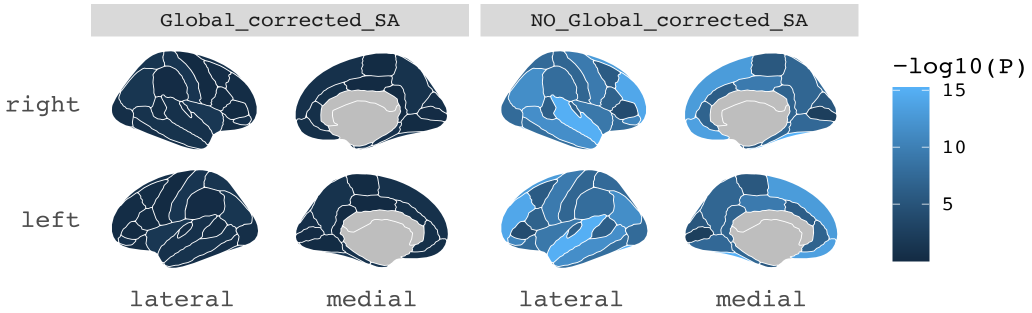

A. Beta coefficients of MR results of regional SA on educational attainment (EA).

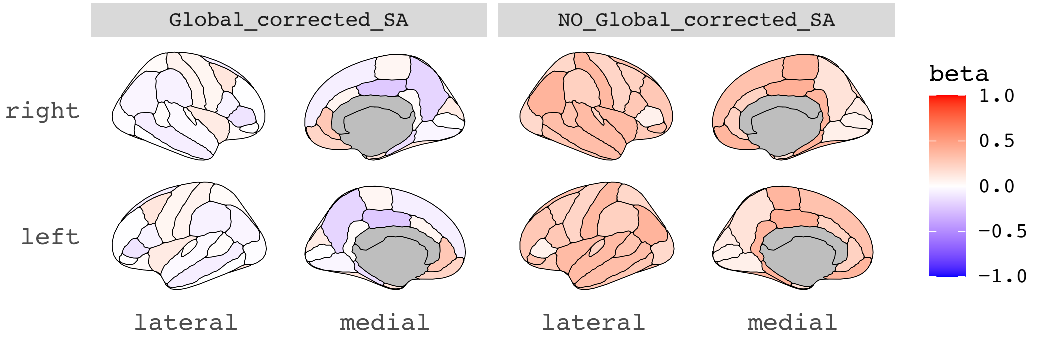

B. -log10 ($p_{.fdr}$ values) of MR results of regional SA on educational attainment (EA).

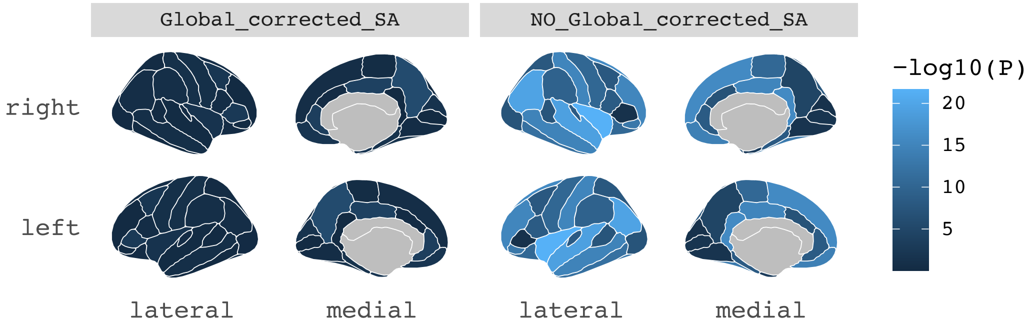

C. Beta coefficients of MR results of educational attainment (EA) on regional SA.

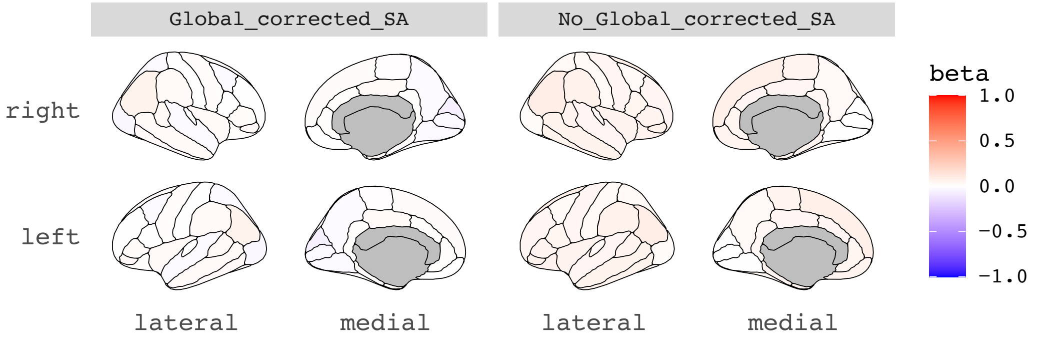

D. -log10 ($p_{.fdr}$ values) of MR results of educational attainment (EA) on regional SA.

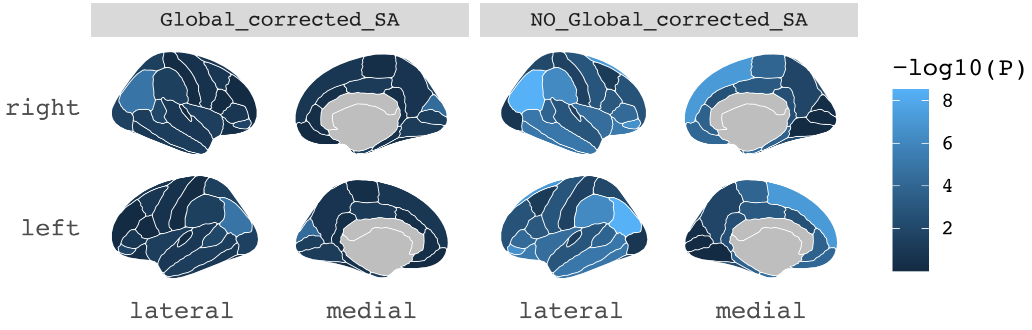

A. Beta coefficients of MR results of regional SA on height.

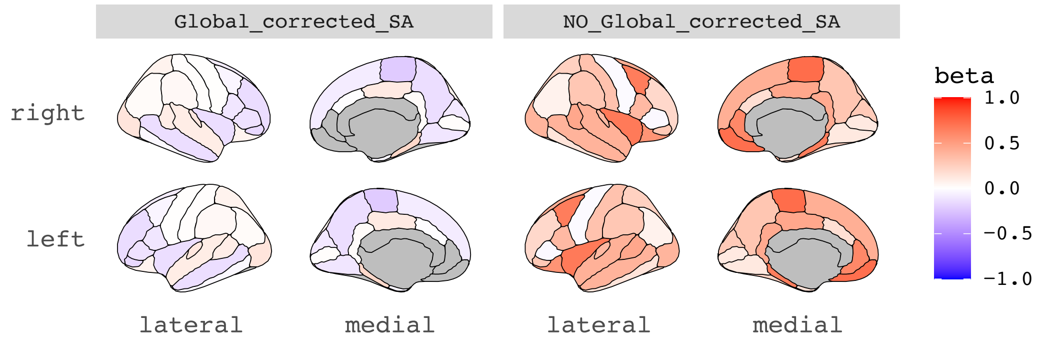

B. -log10 ($p_{fdr}$ values) of MR results of regional SA on height.

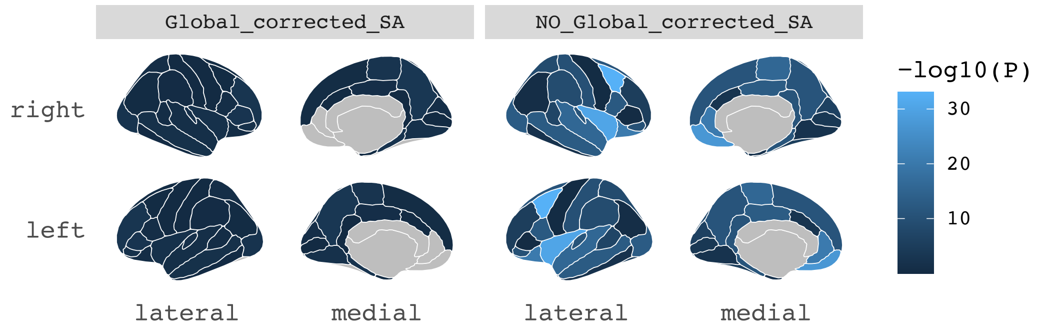

C. Beta coefficients of MR results of height on regional SA.

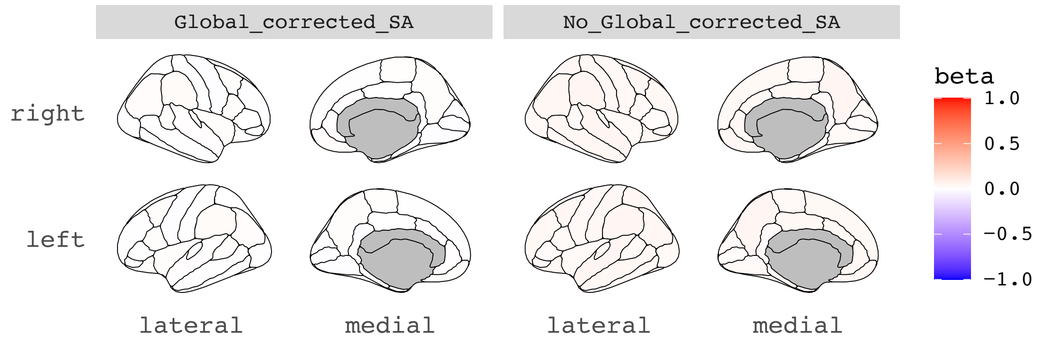

D. -log10 ($p_{fdr}$ values) of MR results of height on regional SA.

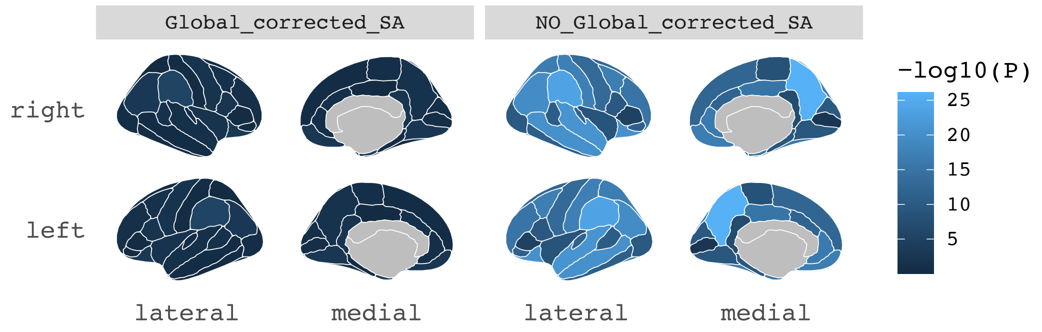

A. Beta coefficients of MR results of regional SA on log-transformed triglycerides (logTG).

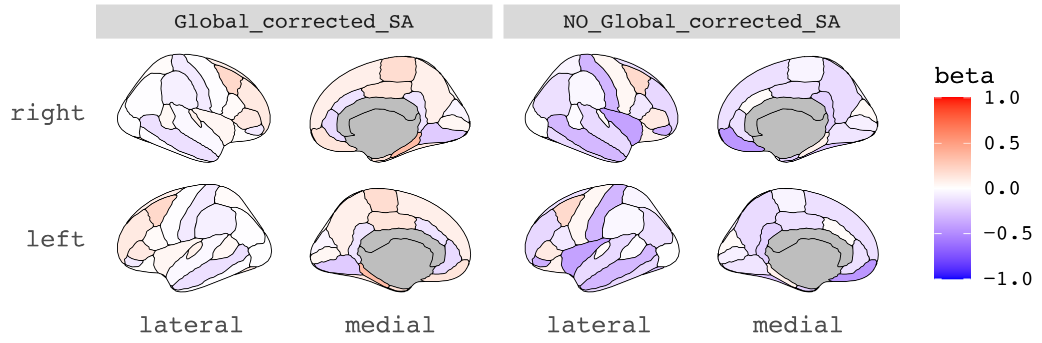

B. -log10 ($p_{fdr}$ values) of MR results of regional SA on log-transformed triglycerides (logTG).

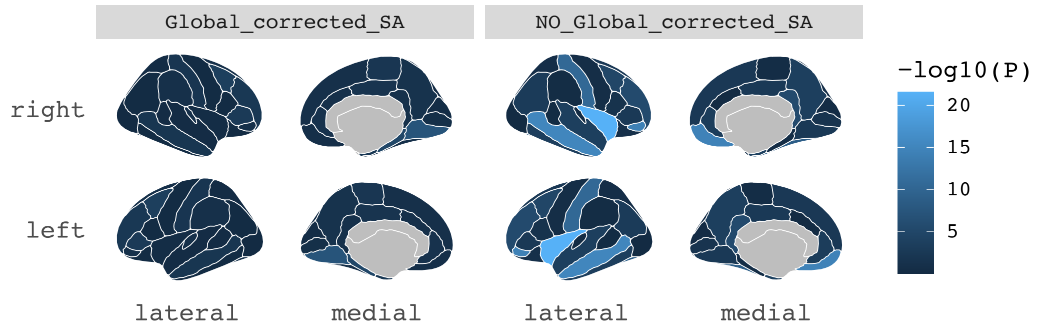

C. Beta coefficients of MR results of log-transformed triglycerides (logTG) on regional SA.

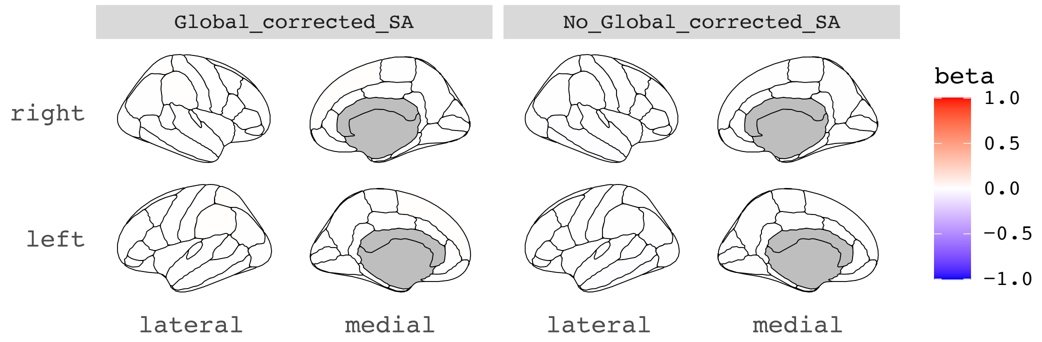

D. -log10 ($p_{fdr}$ values) of MR results of regional log-transformed triglycerides (logTG) on regional SA.

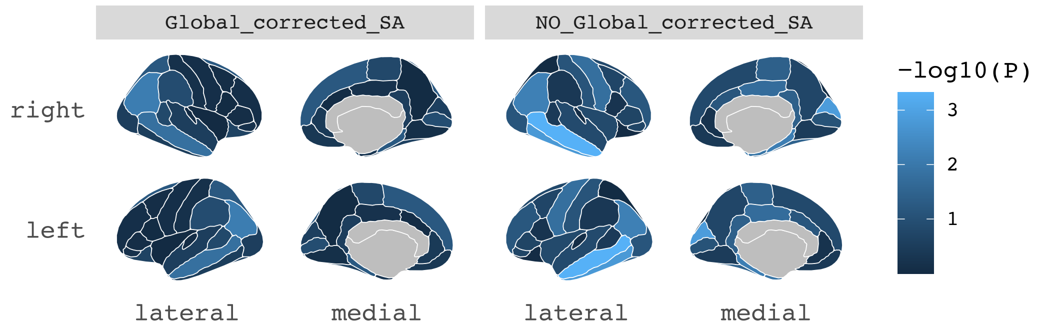

A. Beta coefficients of MR results of regional SA on smoking initiation (2022) (SmkInit#2).

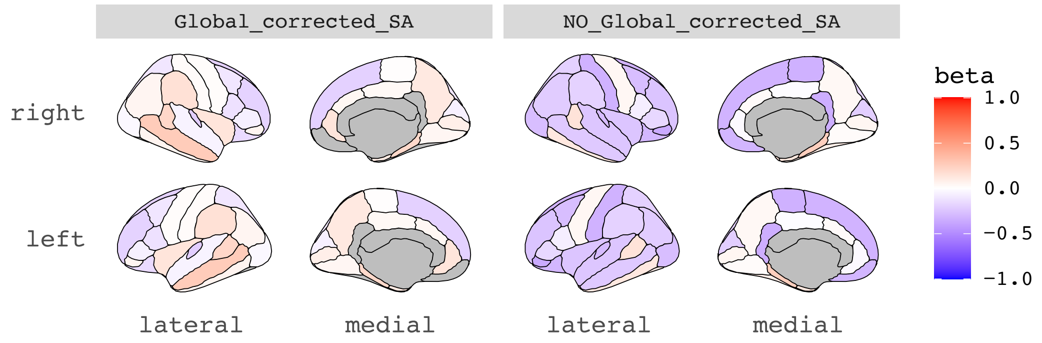

B. -log10 ($p_{fdr}$ values) of MR results of regional SA on smoking initiation (2022) (SmkInit#2).

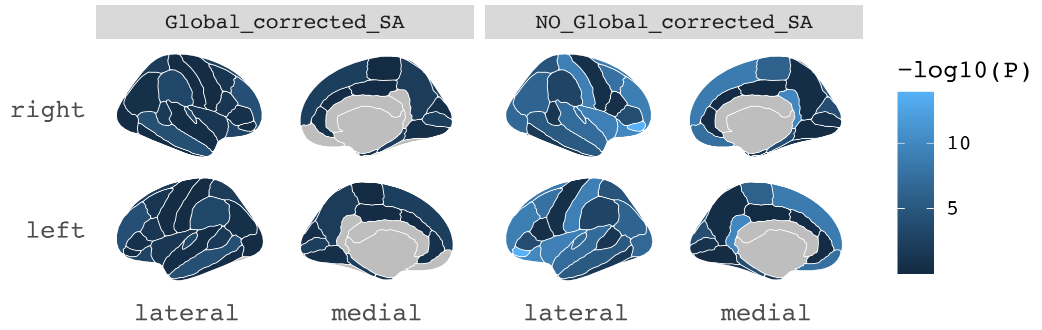

C. Beta coefficients of MR results of smoking initiation (2022) (SmkInit#2) on regional SA.

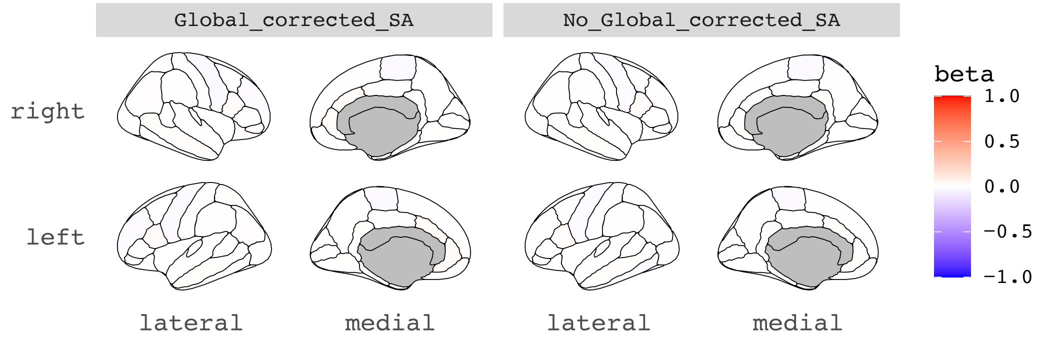

D. -log10 ($p_{fdr}$ values) of MR results of smoking initiation (2022) (SmkInit#2) on regional SA.

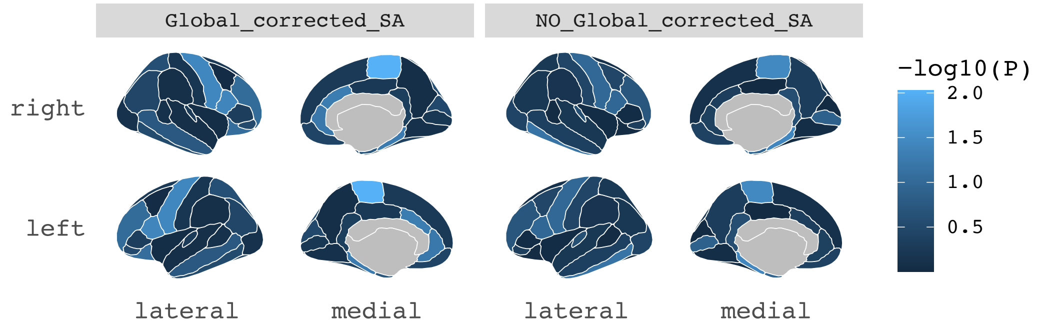

A. Beta coefficients of MR results of regional SA on smoking initiation (2019) (SmkInit).

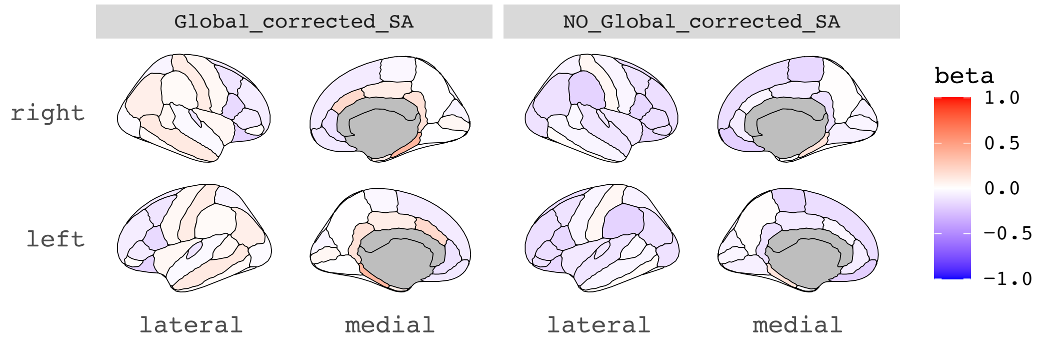

B. -log10 (p.fdr values) of MR results of regional SA on smoking initiation (2019) (SmkInit).

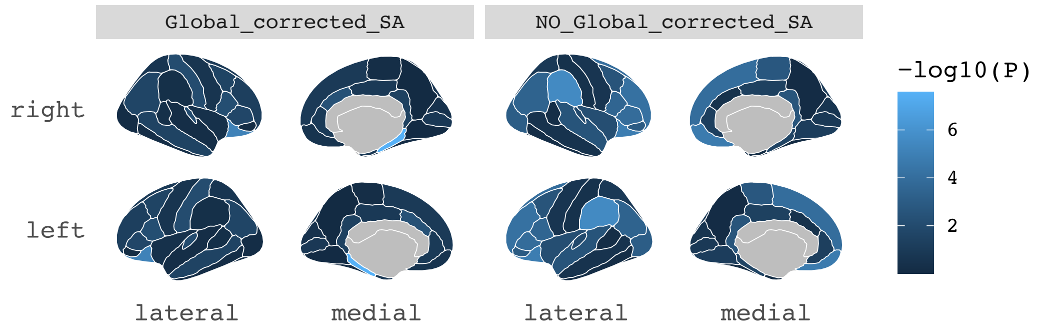

C. Beta coefficients of MR results of smoking initiation (2019) (SmkInit) on regional SA.

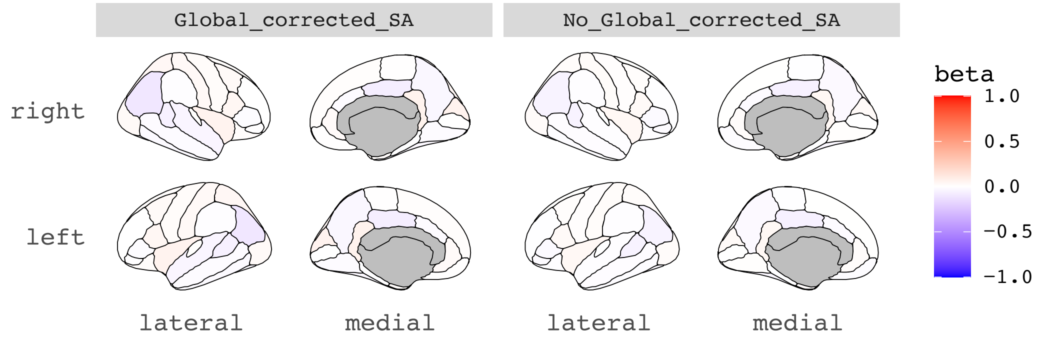

D. -log10 (p.fdr values) of MR results of regional smoking initiation (2019) (SmkInit) on regional SA.

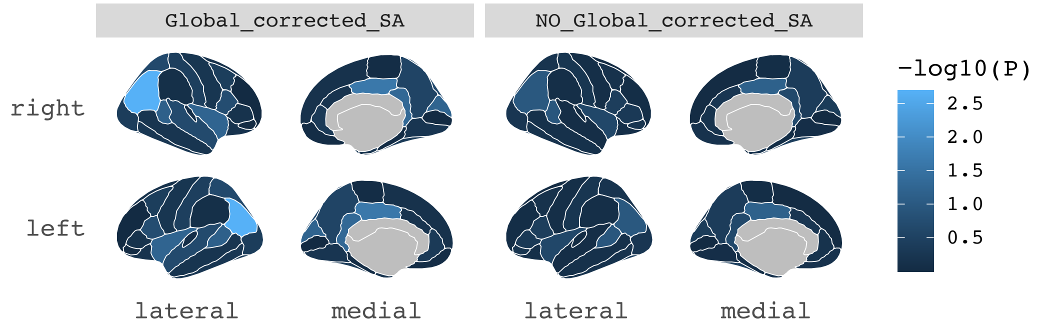

A. Beta coefficients of MR results of regional SA on cross disorders (CDG).

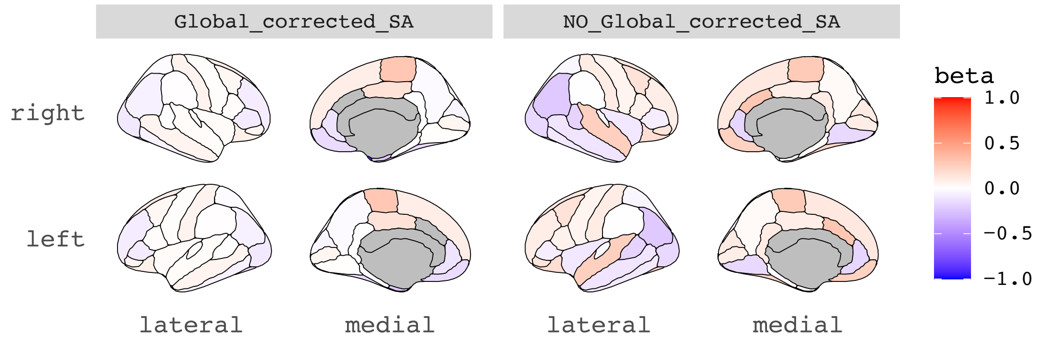

B. -log10 ($p_{.fdr}$ values) of MR results of regional SA on cross disorders (CDG).

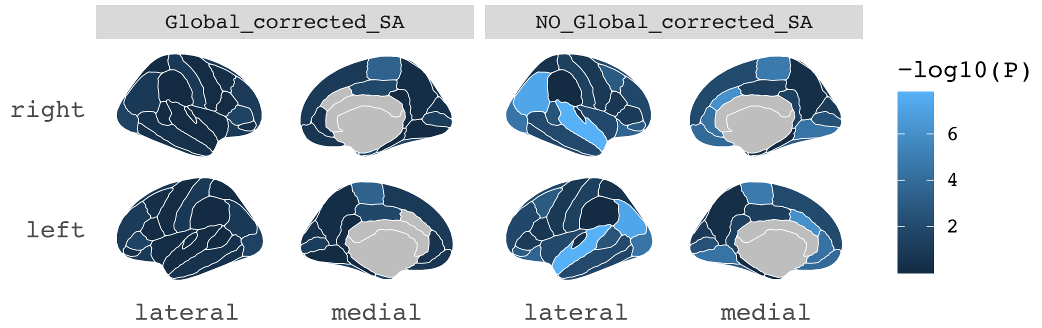

C. Beta coefficients of MR results of cross disorders (CDG) on regional SA.

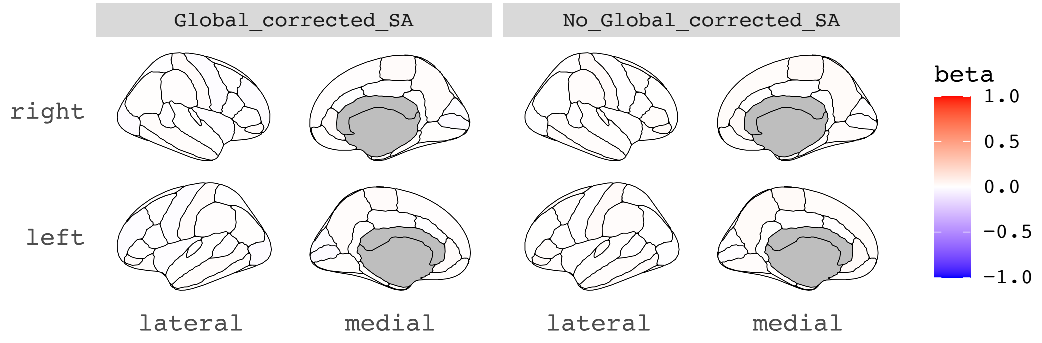

D. -log10 ($p_{.fdr}$ values) of MR results of cross disorders (CDG) on regional SA.

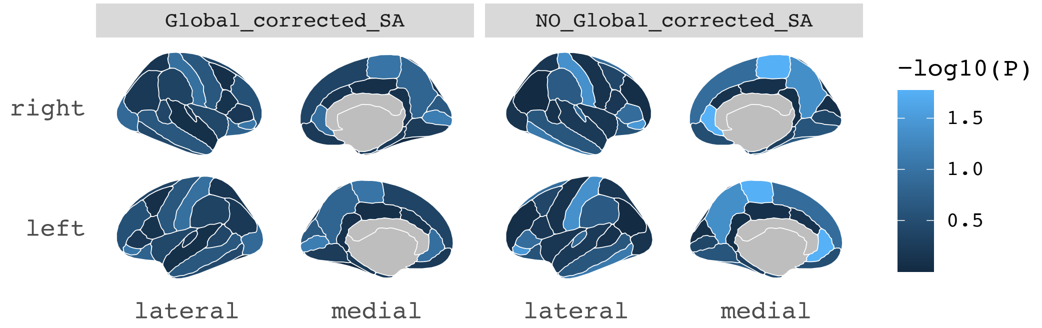

A. Beta coefficients of MR results of regional SA on ADHD2019.

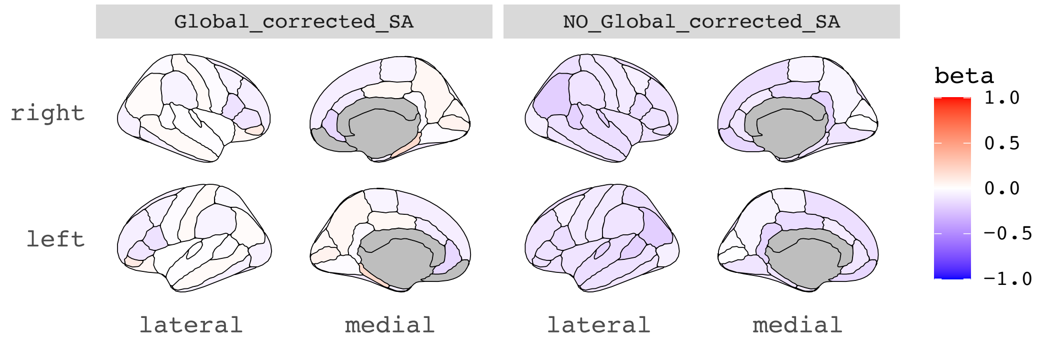

B. -log10 ($p_{fdr}$ values) of MR results of regional SA on ADHD2019.

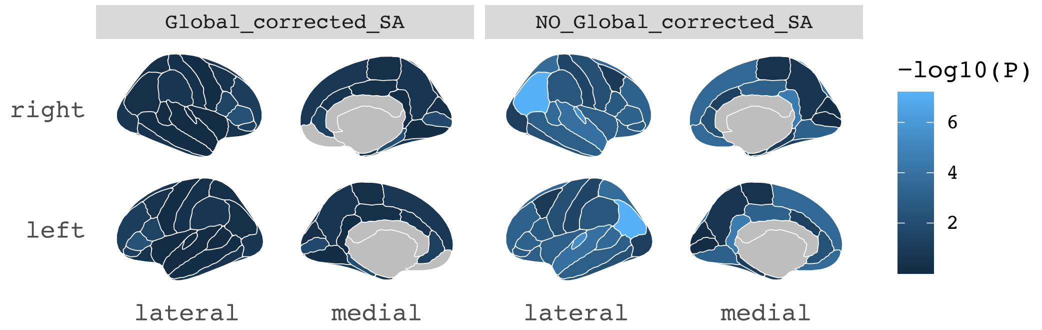

C. Beta coefficients of MR results of ADHD2019 on regional SA.

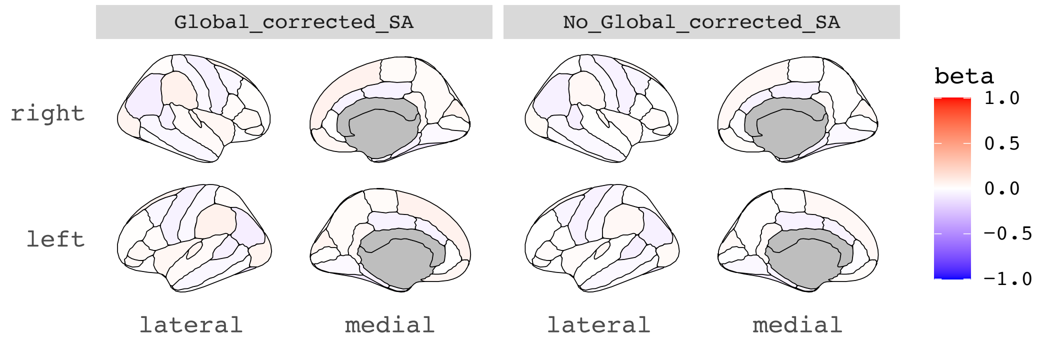

D. -log10 ($p_{fdr}$ values) of MR results of ADHD2019 on regional SA.

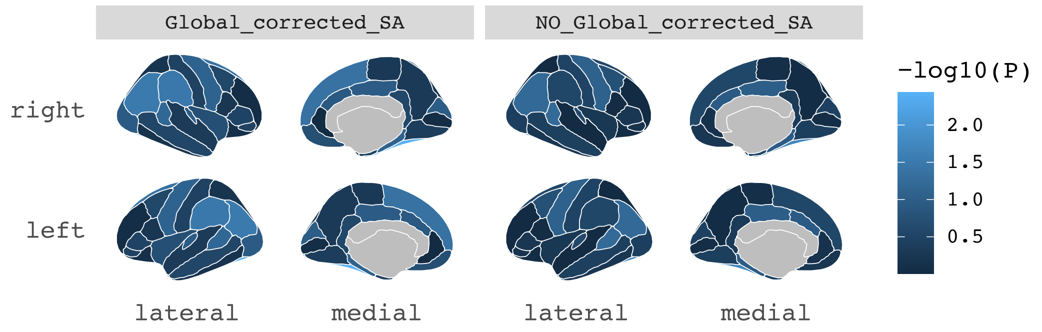

A. Beta coefficients of MR results of regional SA on age of smoking initiation (2022) (AgeSmk#2).

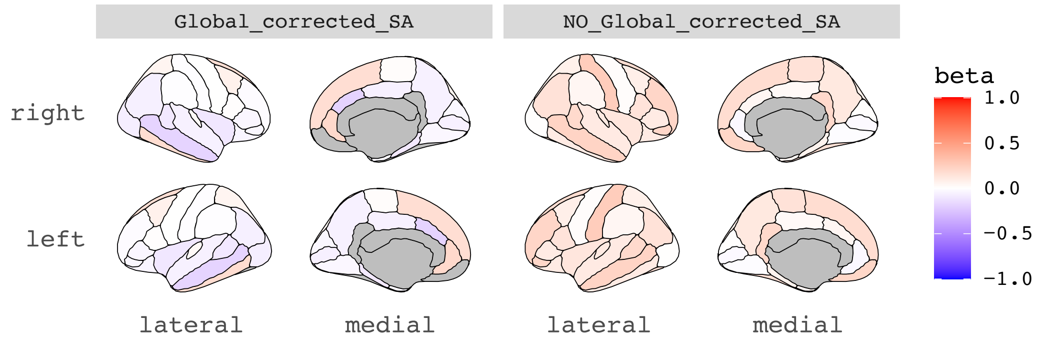

B. -log10 ($p_{fdr}$ values) of MR results of regional SA on age of smoking initiation (2022) (AgeSmk#2).

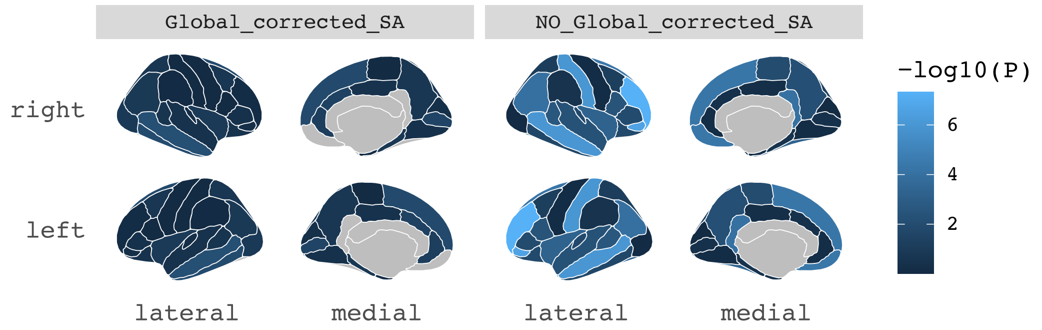

C. Beta coefficients of MR results of age of smoking initiation (2022) (AgeSmk#2) on regional SA.

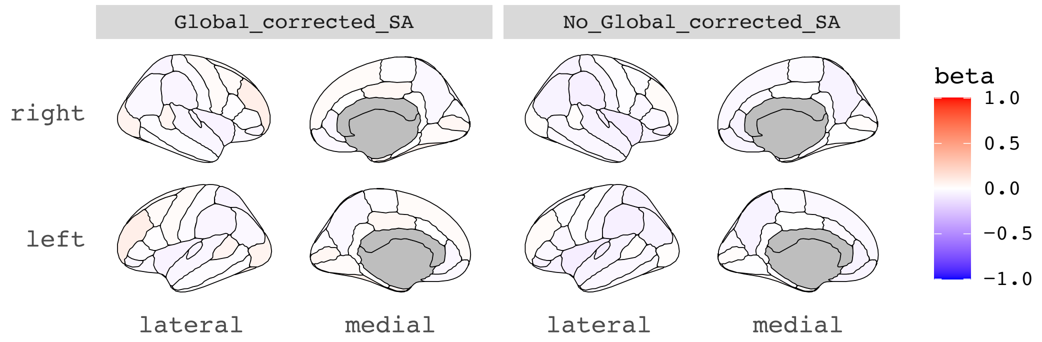

D. -log10 ($p_{fdr}$ values) of MR results of age of smoking initiation (2022) (AgeSmk#2) on regional SA.

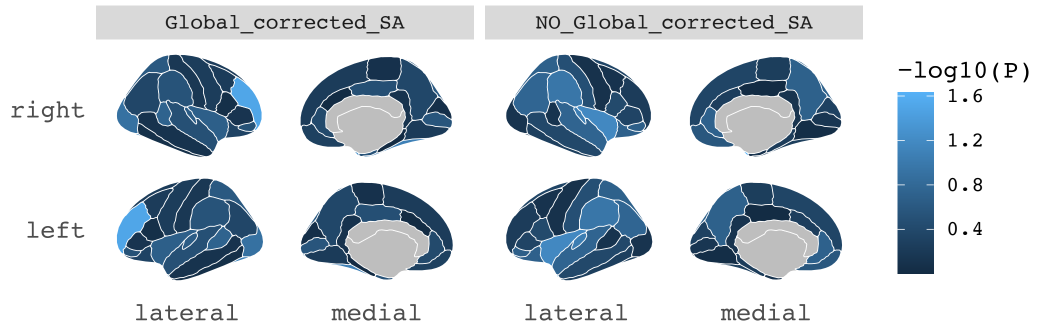

A. Beta coefficients of MR results of regional SA on high-density lipoprotein (HDL).

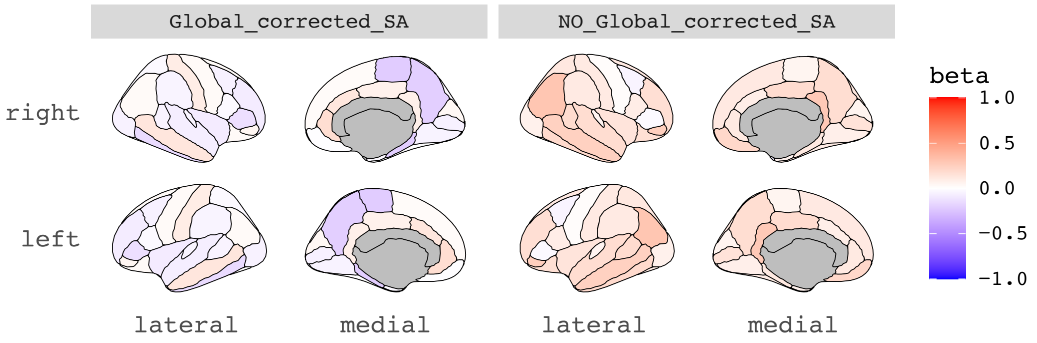

B. -log10 ($p_{fdr}$ values) of MR results of regional SA on high-density lipoprotein (HDL).

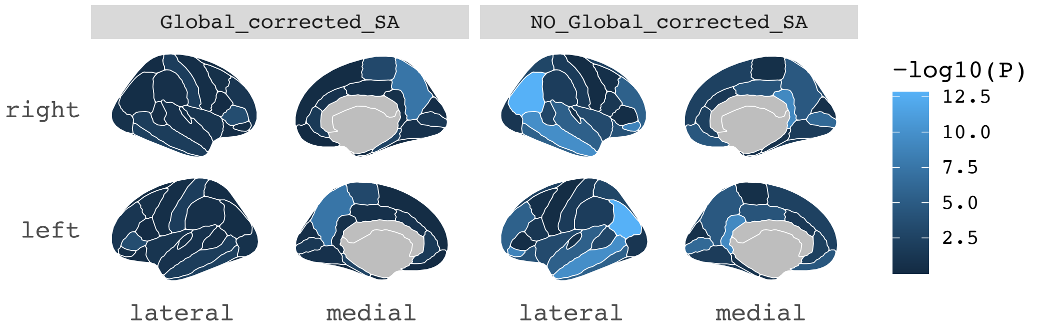

C. Beta coefficients of MR results of high-density lipoprotein (HDL) on regional SA.

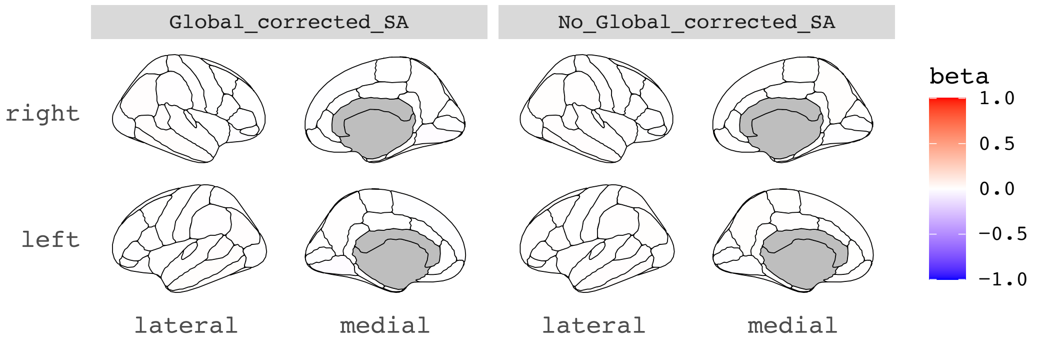

D. -log10 ($p_{fdr}$ values) of MR results of high-density lipoprotein (HDL) on regional SA.

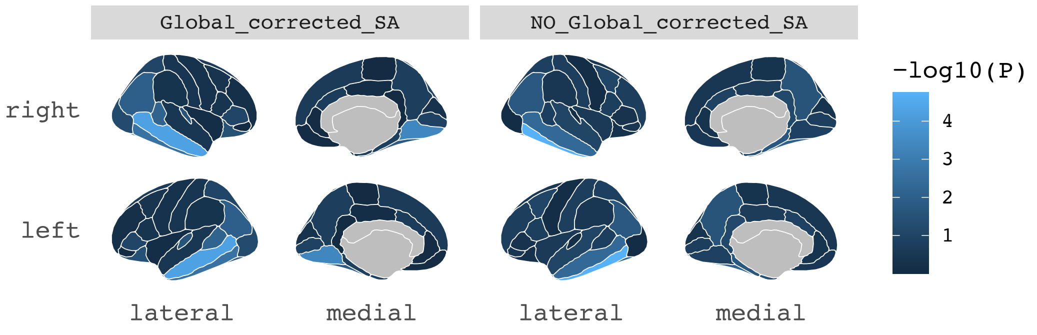

A. Beta coefficients of MR results of regional SA on smoking cessation (2022) (SmkCes#2).

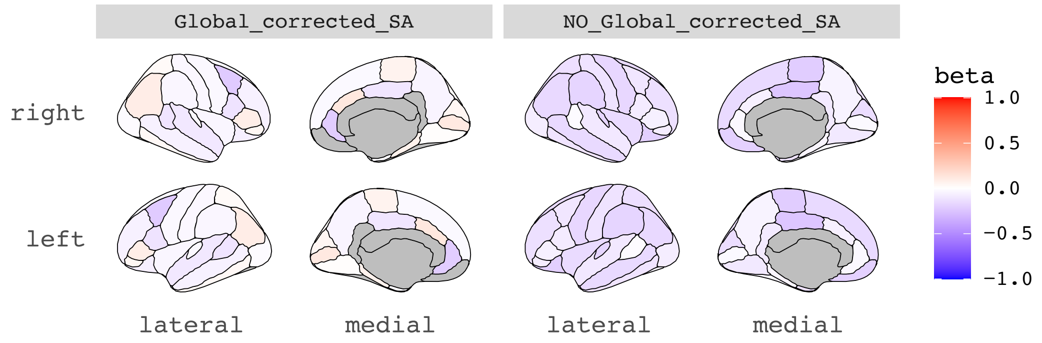

B. -log10 (p.$_{fdr}$ values) of MR results of regional SA on smoking cessation (2022) (SmkCes#2).

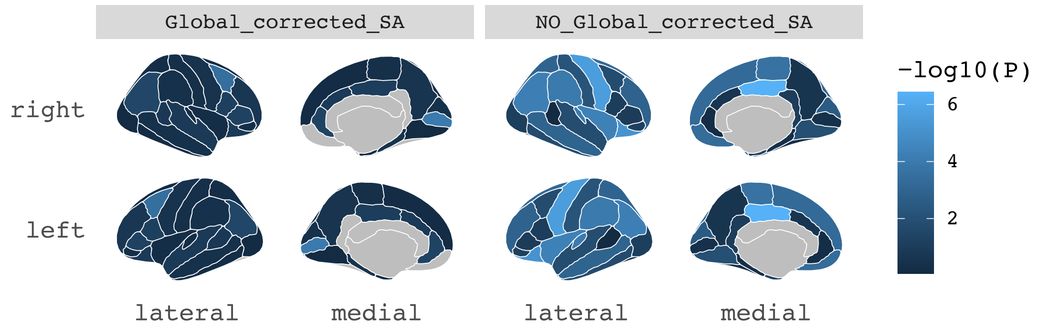

C. Beta coefficients of MR results of smoking cessation (2022) (SmkCes#2) on regional SA.

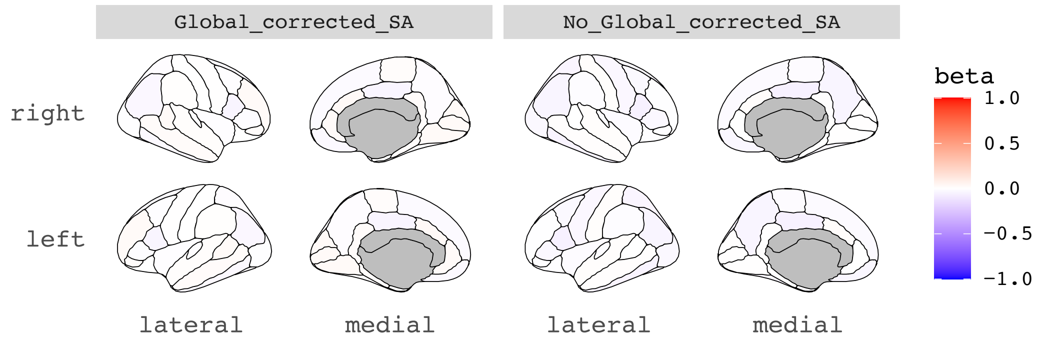

D. -log10 (p.$_{fdr}$ values) of MR results of smoking cessation (2022) (SmkCes#2) on regional SA.

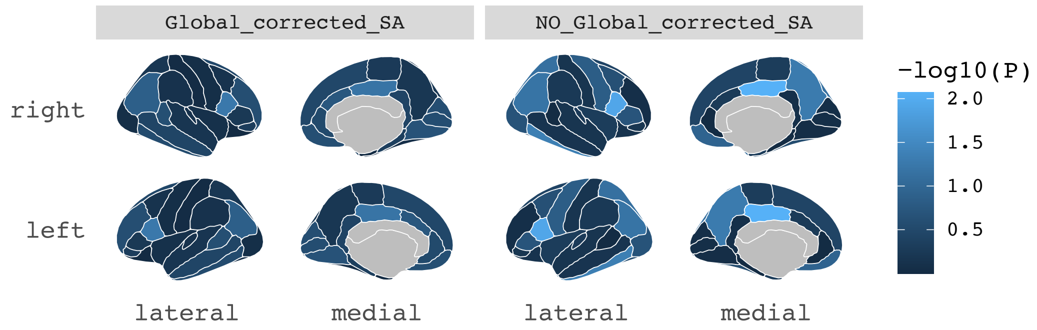

A. Beta coefficients of MR results of regional SA on non-high-density lipoprotein (nonHDL).

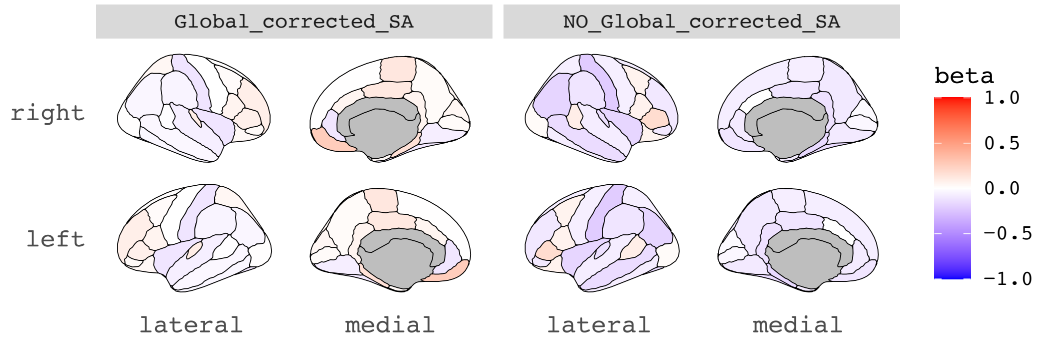

B. -log10 ($p_{.fdr}$ values) of MR results of regional SA on non-high-density lipoprotein (nonHDL).

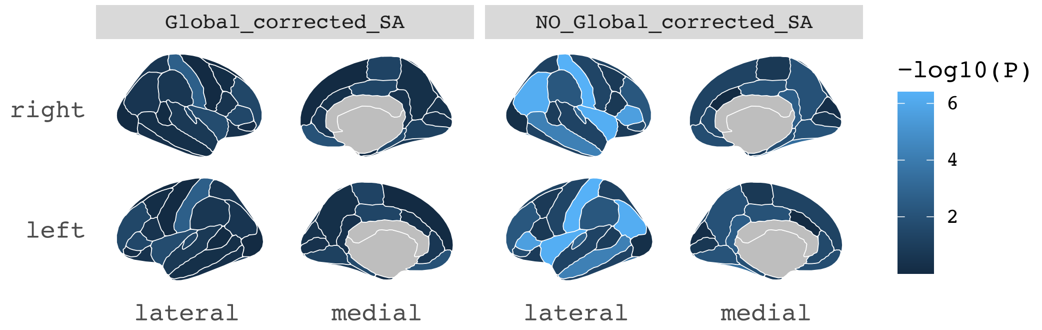

C. Beta coefficients of MR results of non-high-density lipoprotein (nonHDL) on regional SA.

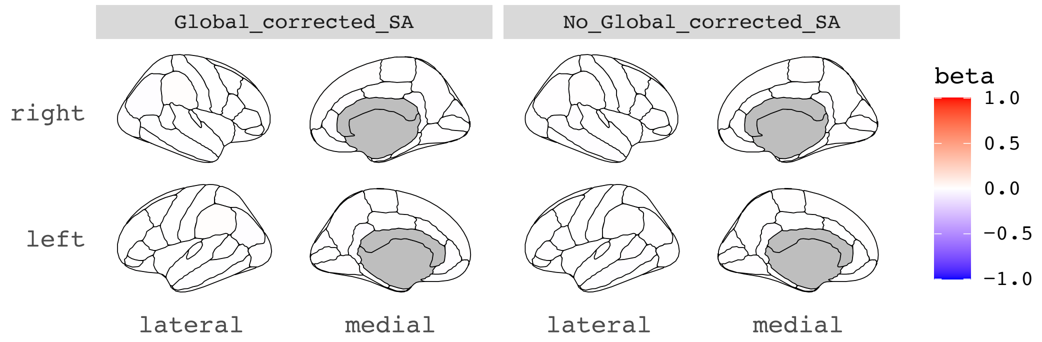

D. -log10 ($p_{.fdr}$ values) of MR results of non-high-density lipoprotein (nonHDL) on regional SA.

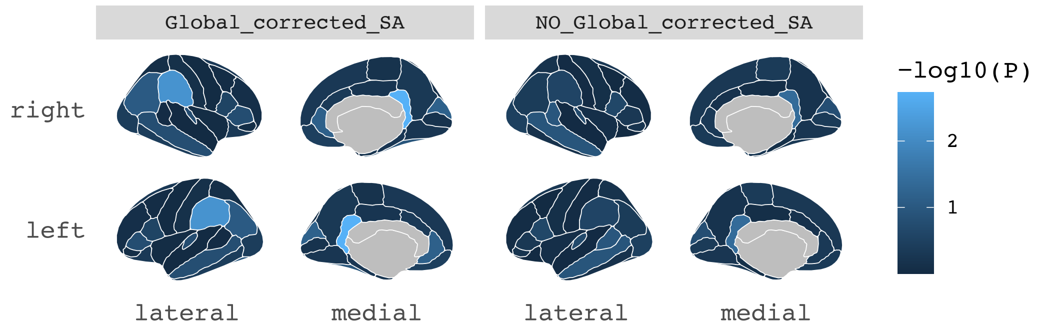

A. Beta coefficients of MR results of regional SA on bipolar disorder (BP - noUKB).

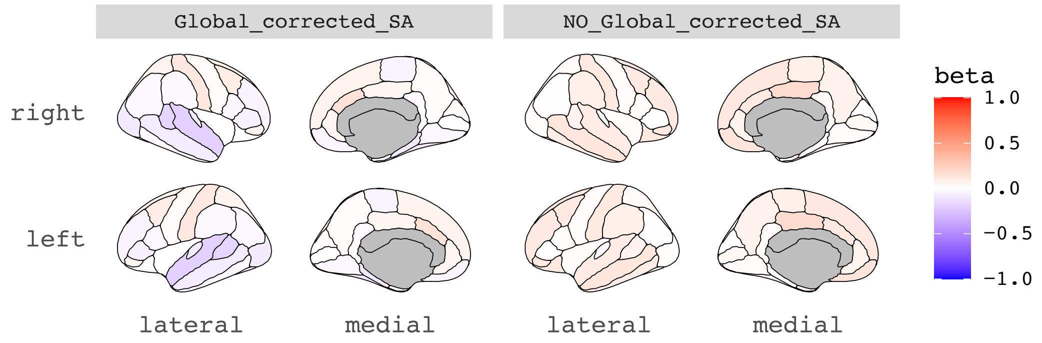

B. -log10 ($p_{fdr}$ values) of MR results of regional SA on bipolar disorder (BP - noUKB).

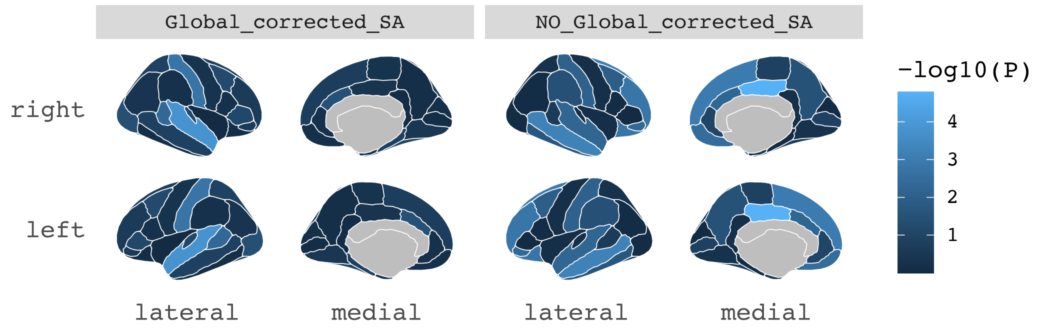

C. Beta coefficients of MR results of bipolar disorder (BP - noUKB) on regional SA.

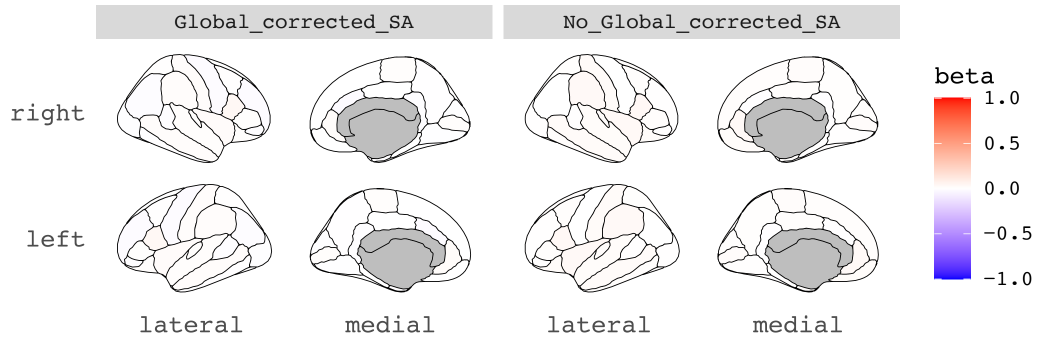

D. -log10 ($p_{fdr}$ values) of MR results of (BP - noUKB) on regional SA.

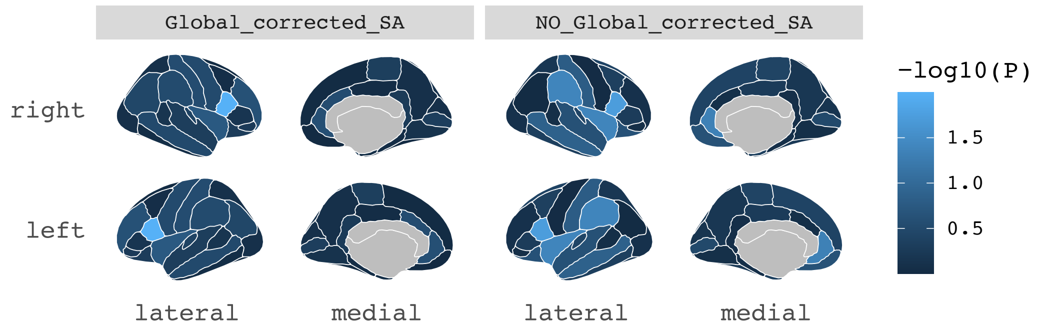

A. Beta coefficients of MR results of regional SA on total cholesterol (TC).

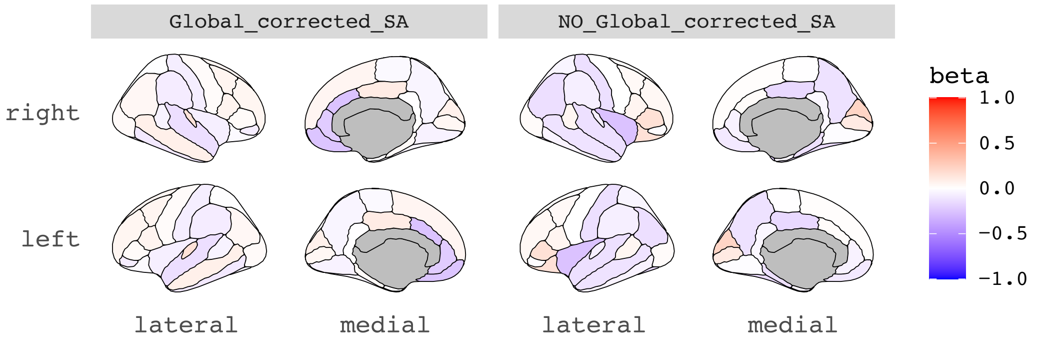

B. -log10 (p.$_{fdr}$ values) of MR results of regional SA on total cholesterol (TC).

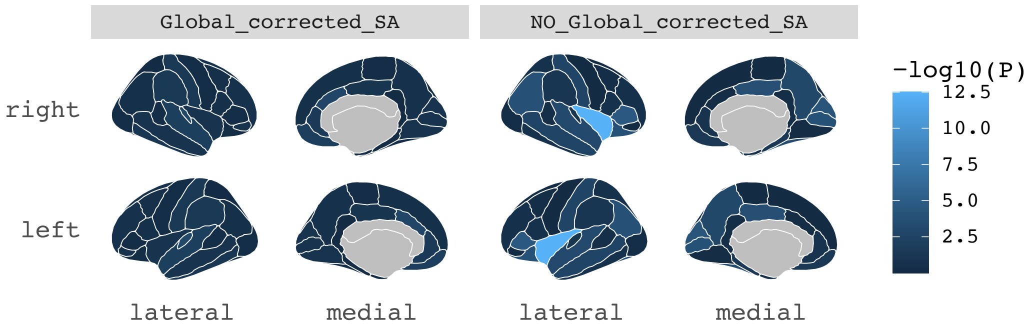

C. Beta coefficients of MR results of total cholesterol (TC) on regional SA.

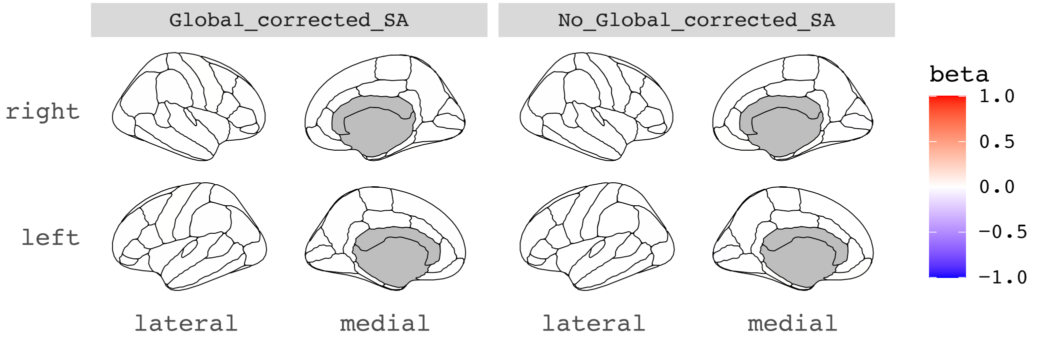

D. -log10 (p.$_{fdr}$ values) of MR results of total cholesterol (TC) on regional SA.

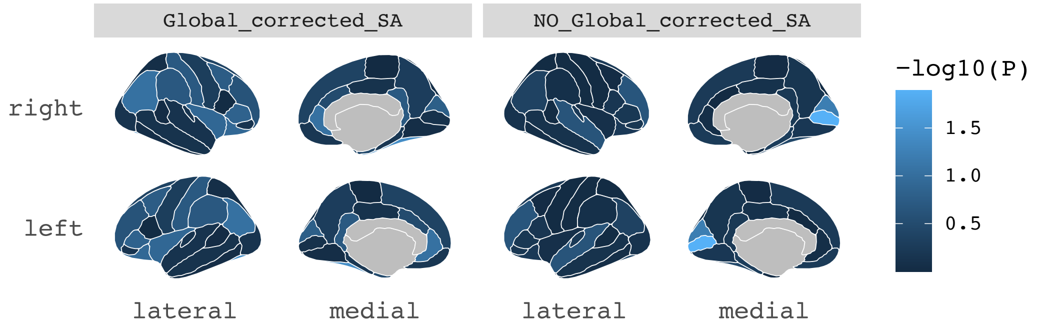

A. Beta coefficients of MR results of regional SA on age of smoking initiation (2019) (AgeSmk).

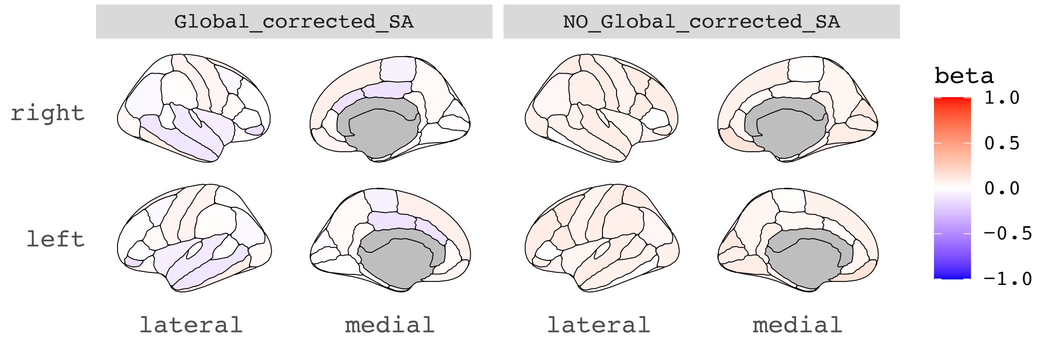

B. -log10 ($p_{.fdr}$ values) of MR results of regional SA on age of smoking initiation (2019) (AgeSmk).

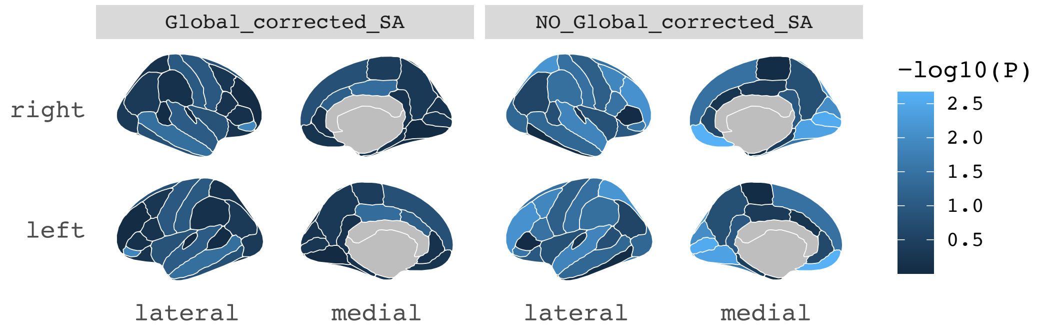

**Note**: The phenotype "age of smoking initiation (2019)" did not have enough independent SNPs to be extracted as instruments and therefore we could not perform reverse MR analyses on SA regions. Hence, panels C and D are missing.

A. Beta coefficients of MR results of regional SA on Alzheimer's disease (ALZ).

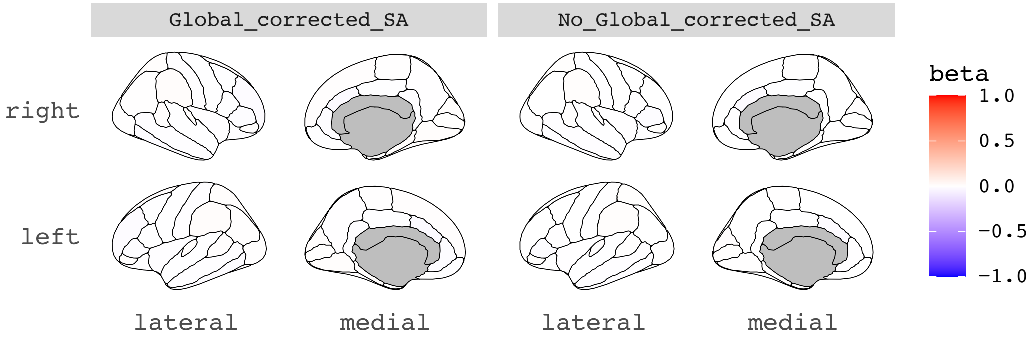

B. -log10 ($p_{fdr}$ values) of MR results of regional SA on Alzheimer's disease (ALZ).

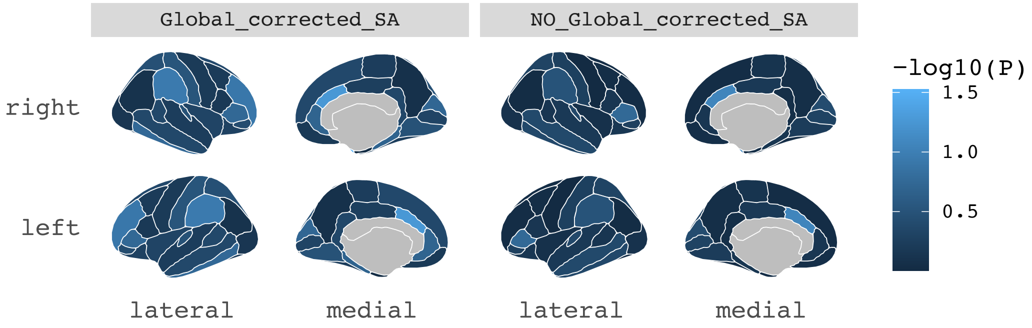

C. Beta coefficients of MR results of Alzheimer's disease (ALZ) on regional SA.

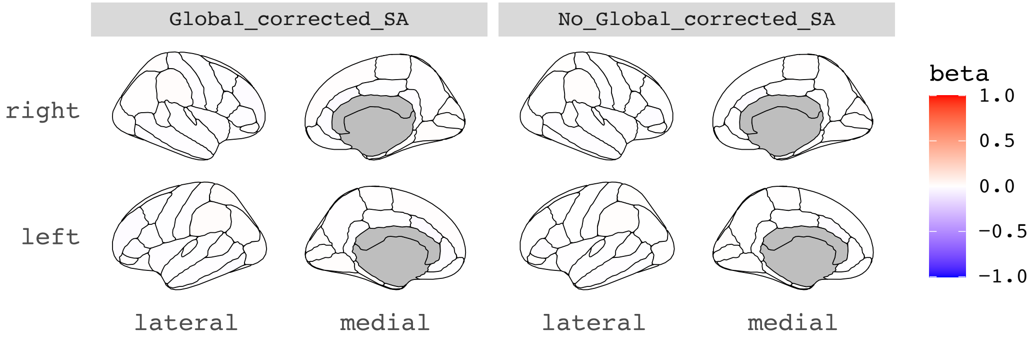

D. -log10 ($p_{fdr}$ values) of MR results of Alzheimer's disease (ALZ) on regional SA.

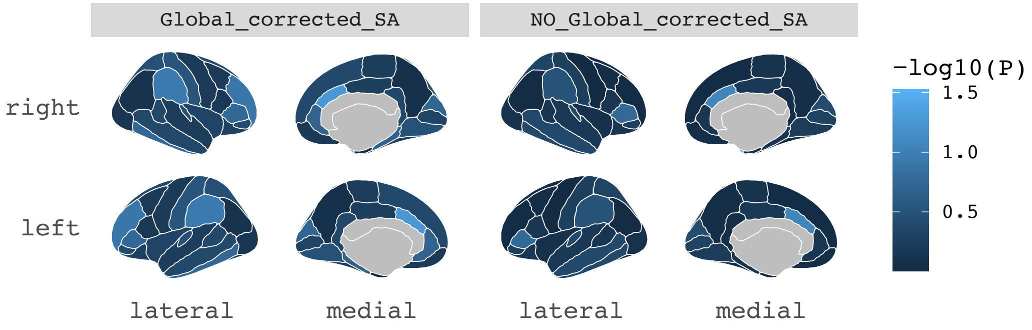

A. Beta coefficients of MR results of regional SA on Type II Diabetes (T2D2020).

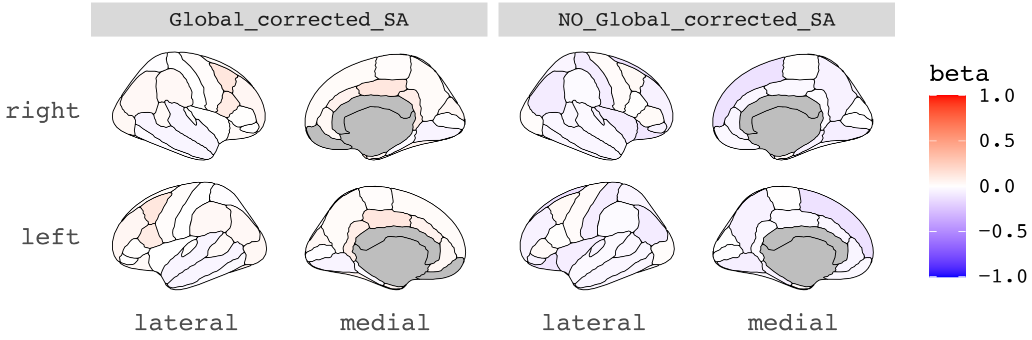

B. -log10 ($p_{fdr}$ values) of MR results of regional SA on Type II Diabetes (T2D2020).

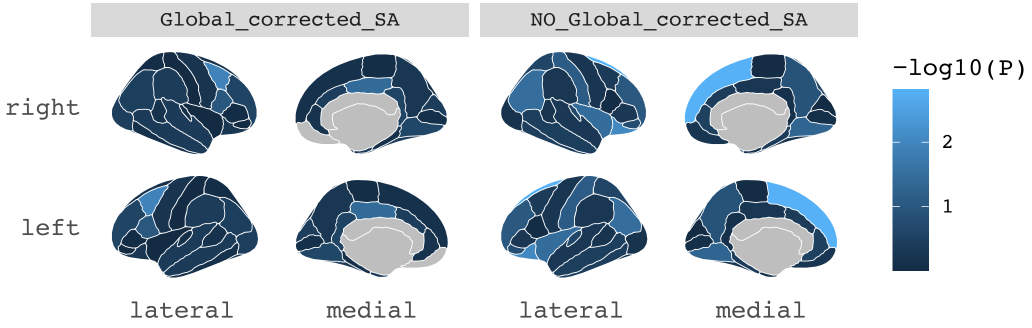

C. Beta coefficients of MR results of Type II Diabetes (T2D2020) on regional SA.

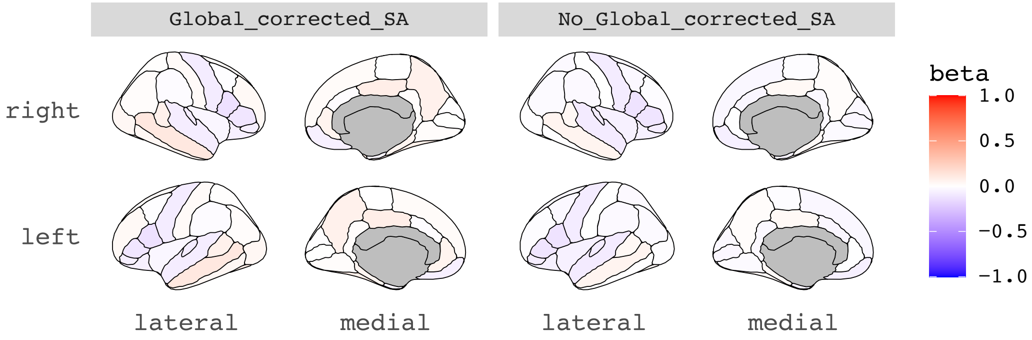

D. -log10 ($p_{fdr}$ values) of MR results of Type II Diabetes (T2D2020) on regional SA.

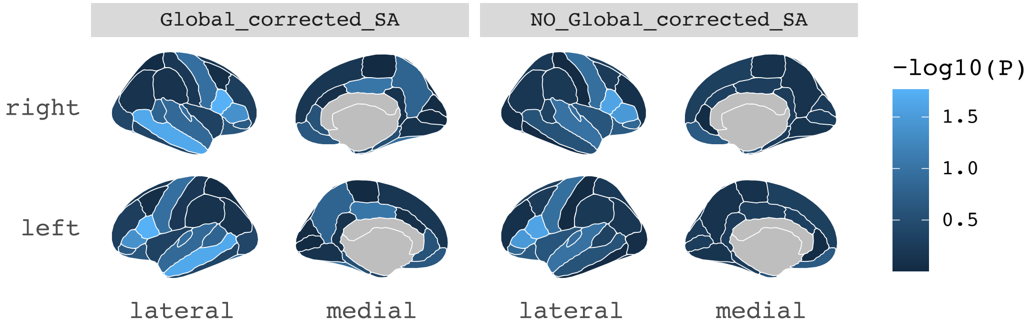

A. Beta coefficients of MR results of regional SA on Type II Diabetes (T2D2017).

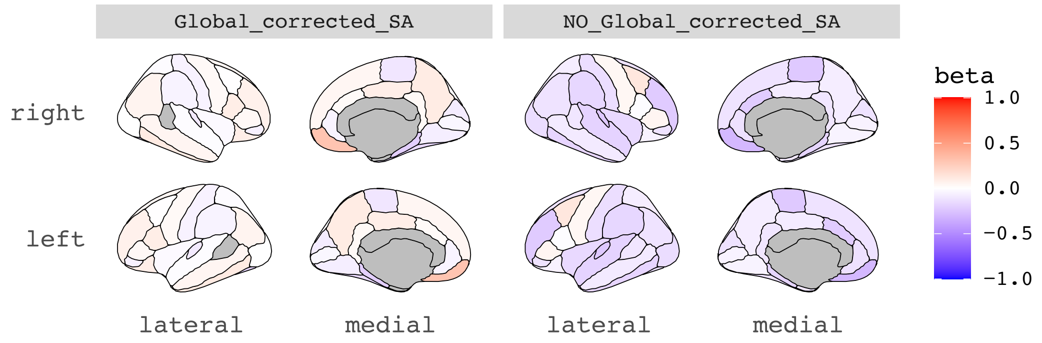

B. -log10 ($p_{fdr}$ values) of MR results of regional SA on Type II Diabetes (T2D2017).

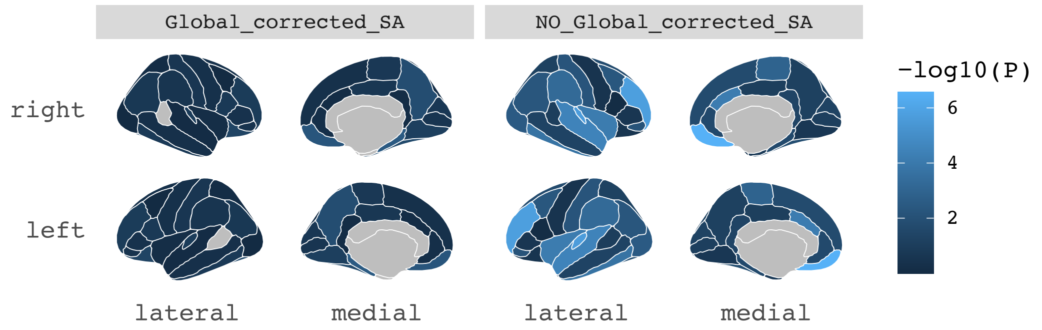

C. Beta coefficients of MR results of Type II Diabetes (T2D2017) on regional SA.

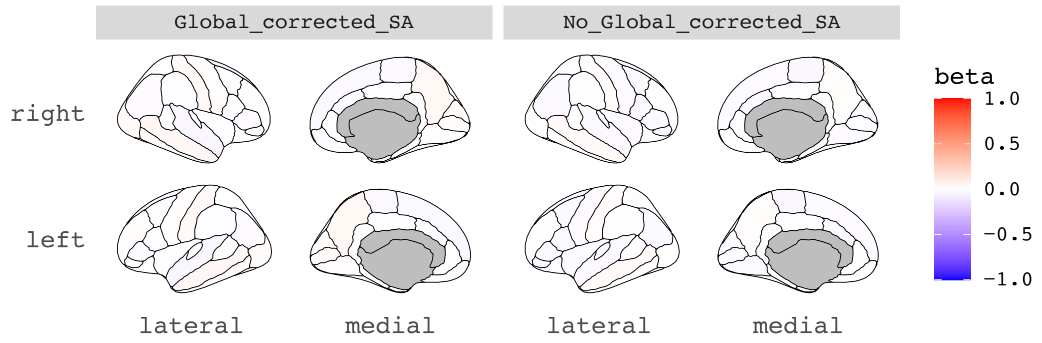

D. -log10 ($p_{fdr}$ values) of MR results of Type II Diabetes (T2D2017) on regional SA.

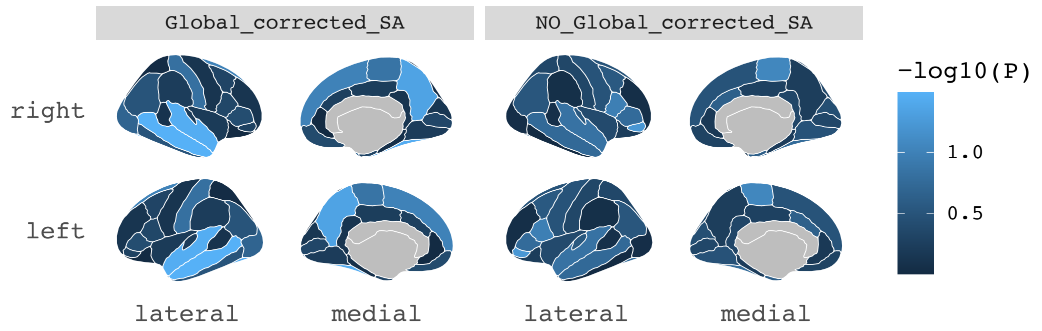

**SFigure 7. Regional plots for cortical thickness (TH) with top causality results, as determined from the general GSMR analyses (forward and reverse).**

Global corrected (left) and non-global-corrected (right) MR results between TH regional measures and the significant results, as determined by the forward and reverse analyses between the global mean TH and the phenotypes (see Supplementary Data ST4, ST5). In panels A and C, the color intensity represents the strength of the causal association via beta coefficients (red: strong positive, blue: strong negative). Panels B and D show the -log10 p-value after FDR-correction: the higher this value (i.e., the lighter the blue), the more statistically significant the result. Visualization with the use of the "ggseg"[42] R package in a plot of the Desikan-Killiany atlas, right hemisphere (upper) and left hemisphere (lower). Lateral and medial sections of the brain are also depicted. Statistical tests were two-sided, and p-values were FDR-correction adjusted for multiple testing. Regarding the sample size of GWASs, MTH= 51,665; AgeSmk, SmkInit = 1.2 million; SmkCes#2, SmkInit#2, DrnkWk#2= 3.4 million; SCZ3=161,405; SCZ2=105,308; BDSCZ=41,653; BIP=31,710; BMI=322,154; Bis.DB.ratio, CH2.in.FA, Bis.FA.ratio, otPUFA, DB.in.FA, Crea, CH2.DB.ratio= 24,925.

A. Beta coefficients of MR results of regional TH on schizophrenia (SCZ3).

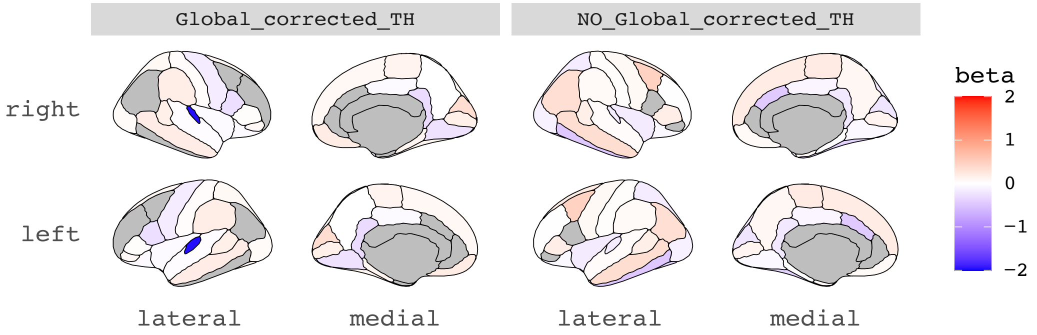

B.-log10 ($p_{fdr}$ values) of MR results of regional TH on schizophrenia (SCZ3).

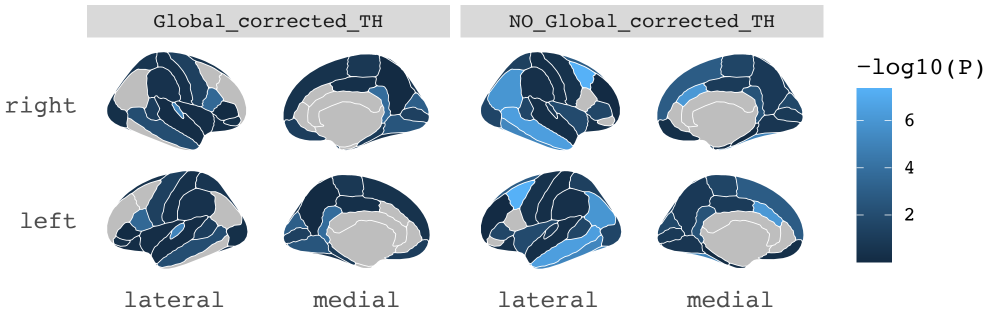

C. Beta coefficients of MR results of schizophrenia (SCZ3) on regional TH.

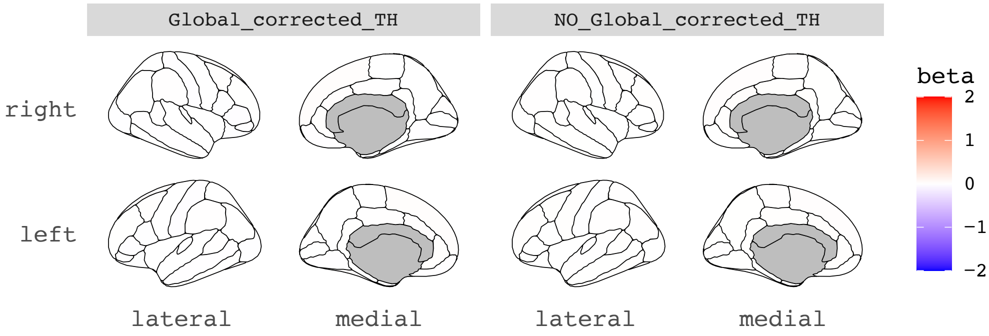

D. -log10 ($p_{fdr}$ values) of MR results of schizophrenia (SCZ3) on regional TH.

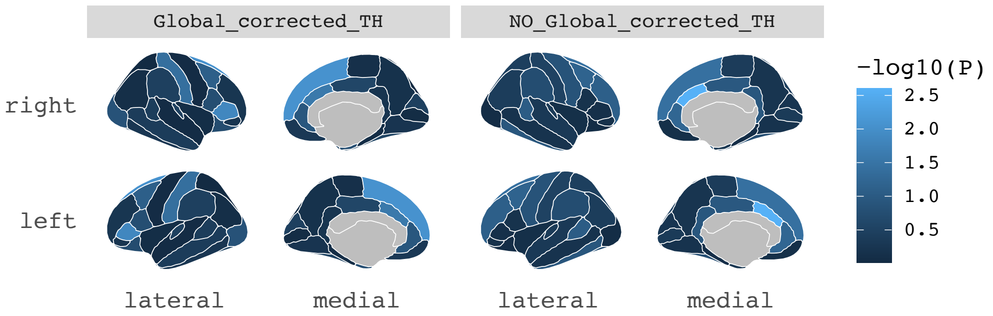

A. Beta coefficients of MR results of regional TH on schizophrenia (SCZ2).

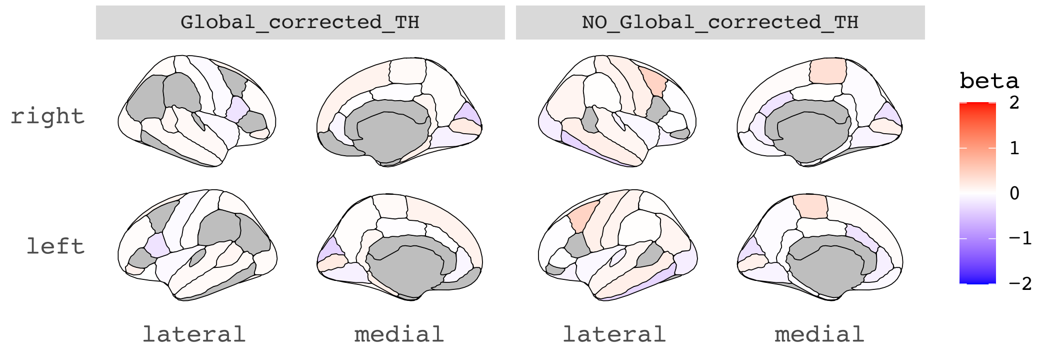

B. -log10 ($p_{.fdr}$ values) of MR results of regional TH on schizophrenia (SCZ2).

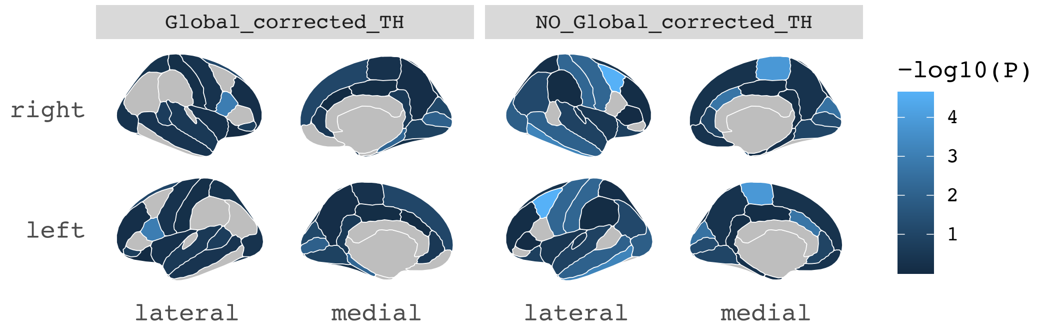

C. Beta coefficients of MR results of schizophrenia (SCZ2) on regional TH.

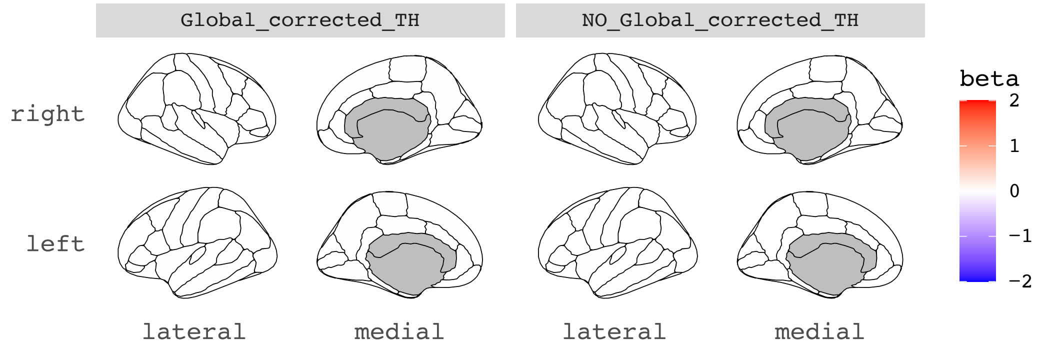

D. -log10 ($p_{.fdr}$ values) of MR results of schizophrenia (SCZ2) on regional TH.

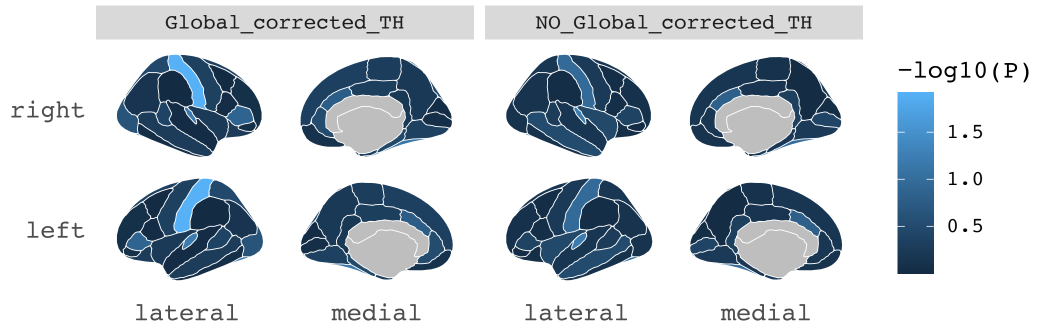

A. Beta coefficients of MR results of regional TH on drinks per week (2022) (DrnkWk#2).

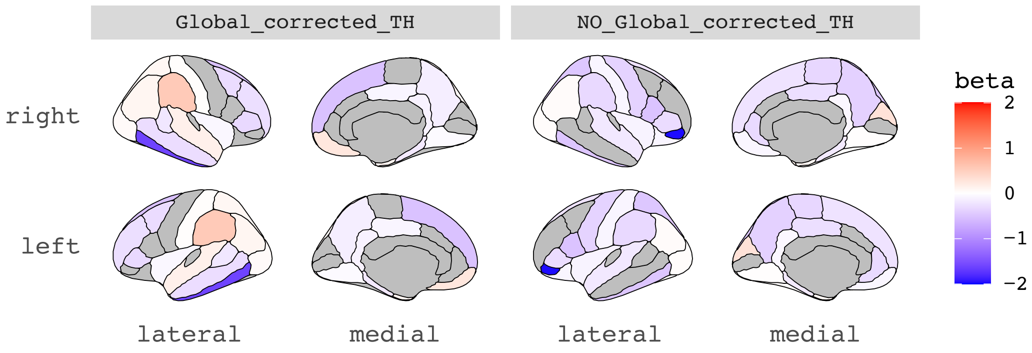

B. -log10 ($p_{.fdr}$ values) of MR results of regional TH on drinks per week (2022) (DrnkWk#2).

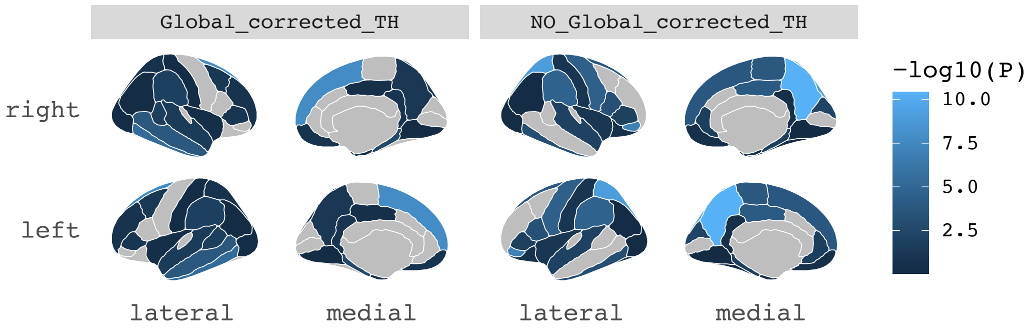

C. Beta coefficients of MR results of drinks per week (2022) (DrnkWk#2) on regional TH.

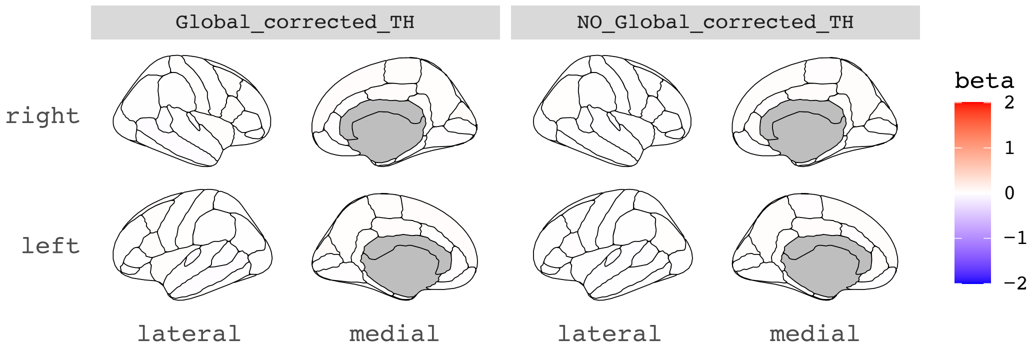

D. -log10 ($p_{.fdr}$ values) of MR results of drinks per week (2022) (DrnkWk#2) on regional TH.

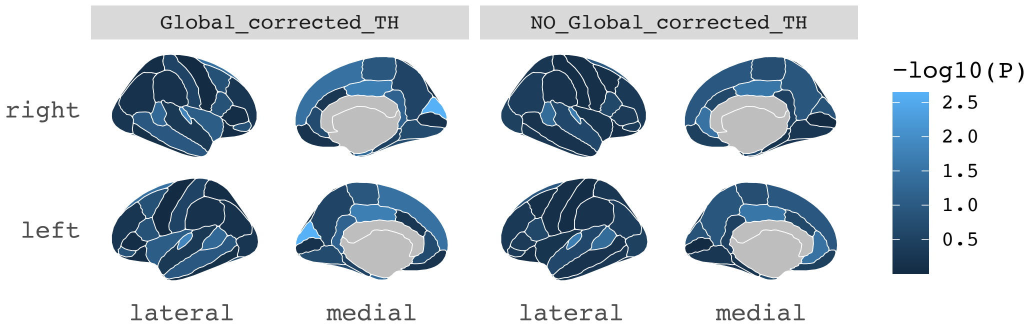

A. Beta coefficients of MR results of regional TH on smoking initiation (2022) (SmkInit#2).

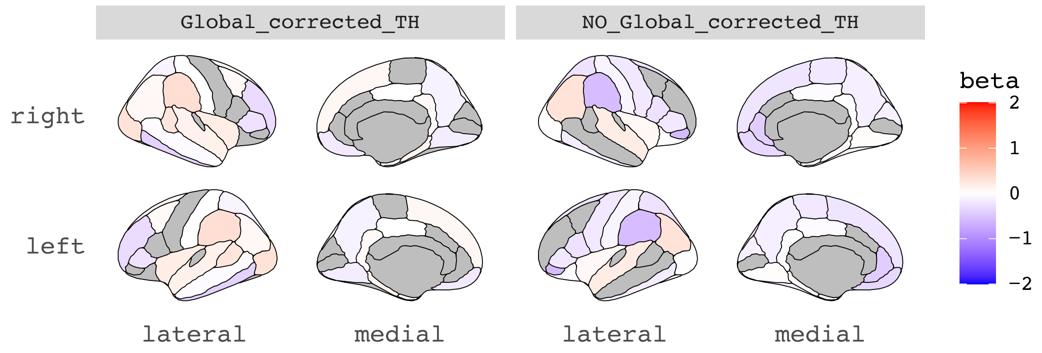

B. -log10 (p.$_{fdr}$ values) of MR results of regional TH on smoking initiation (2022) (SmkInit#2).

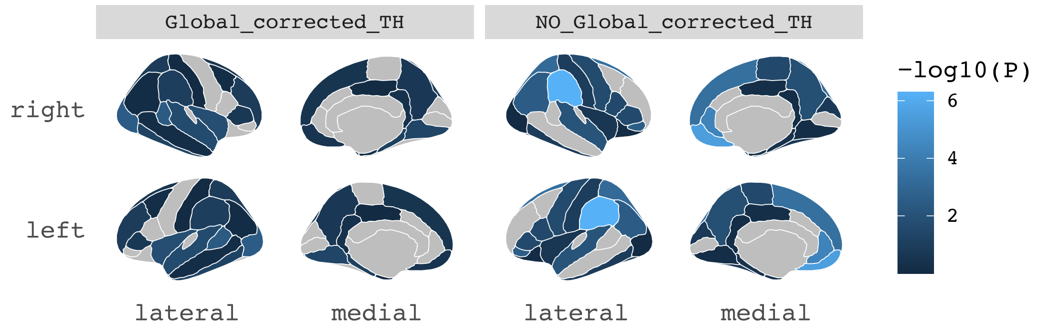

C. Beta coefficients of MR results of smoking initiation (2022) (SmkInit#2) on regional TH.

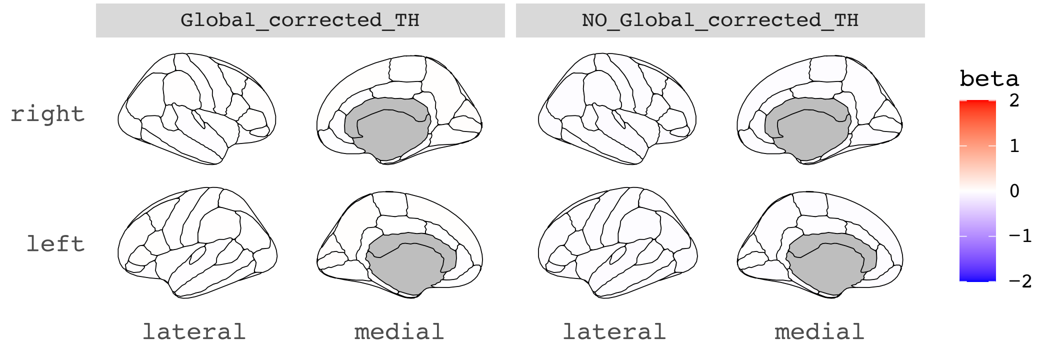

D. -log10 (p.$_{fdr}$ values) of MR results of smoking initiation (2022) (SmkInit#2) on regional TH.

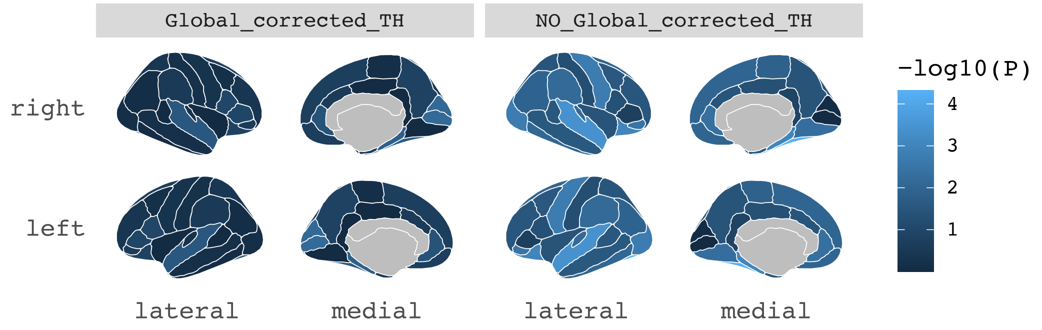

A. Beta coefficients of MR results of regional TH on meta-analysis bipolar disorder-schizophrenia (BDSCZ).

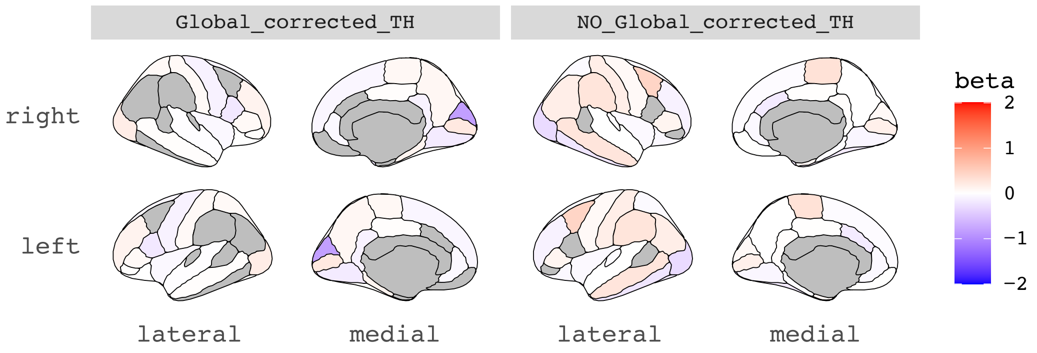

B. -log10 ($p_{.fdr}$ values) of MR results of regional TH on meta-analysis bipolar disorder-schizophrenia (BDSCZ).

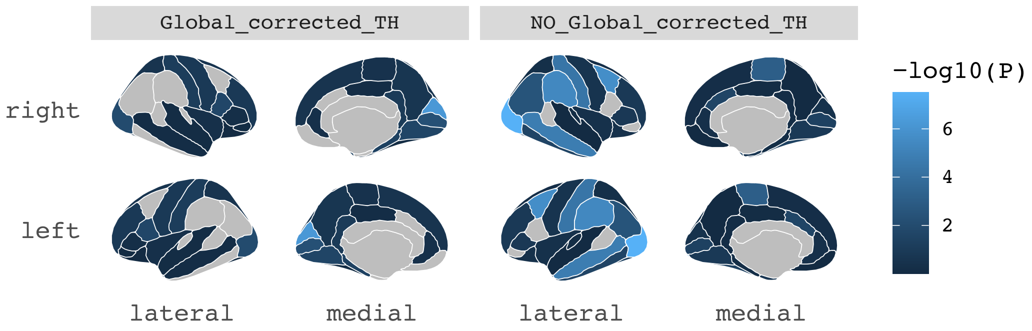

C. Beta coefficients of MR results of meta-analysis bipolar disorder-schizophrenia (BDSCZ) on regional TH.

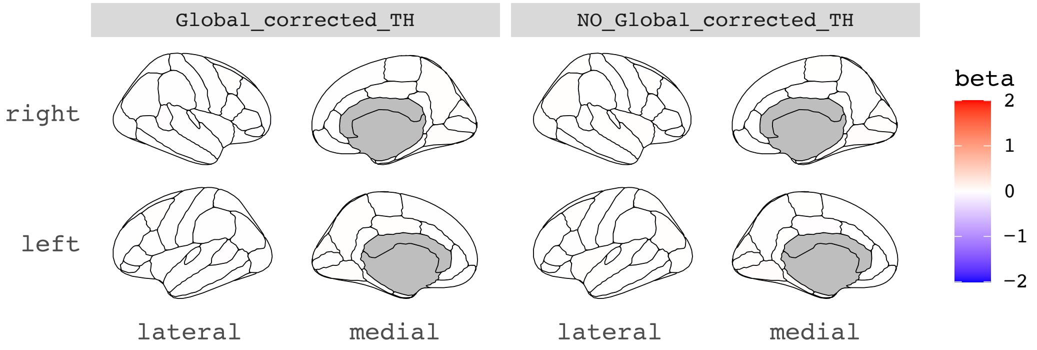

D. -log10 ($p_{.fdr}$ values) of MR results of meta-analysis bipolar disorder-schizophrenia (BDSCZ) on regional TH.

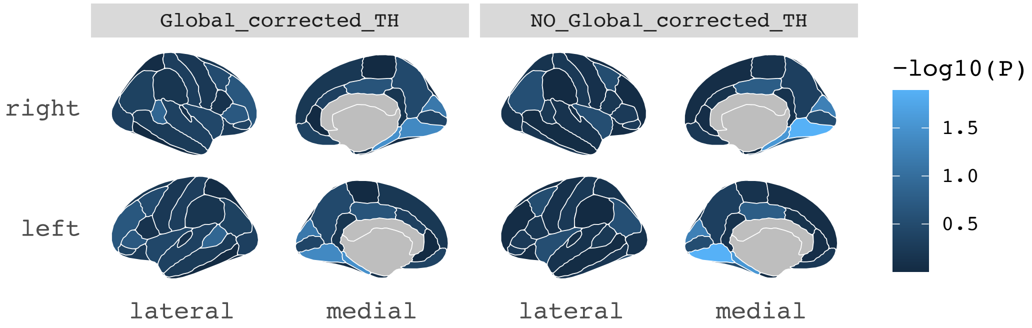

A. Beta coefficients of MR results of regional TH on Bis.DB.ratio.

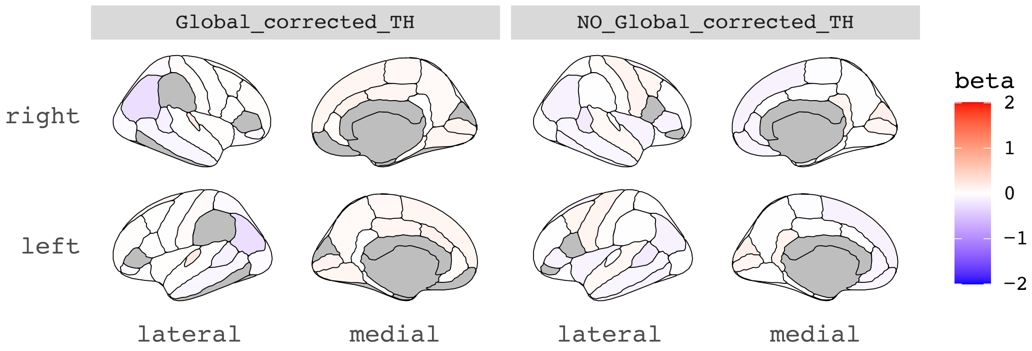

B. -log10 ($p_{fdr}$ values) of MR results of regional TH on Bis.DB.ratio.

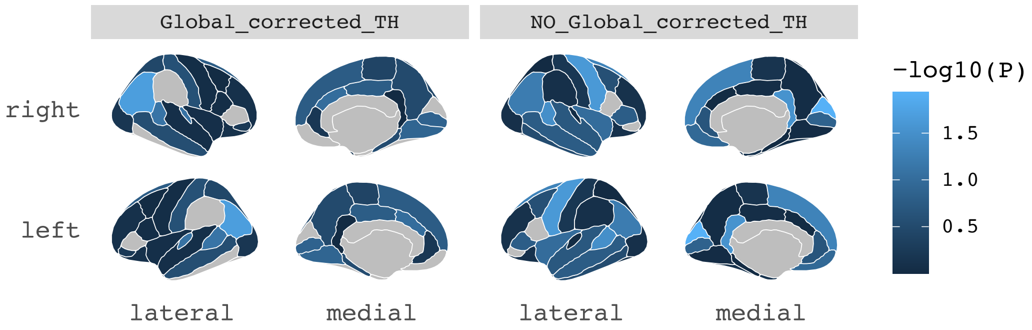

C. Beta coefficients of MR results of Bis.DB.ratio on regional TH.

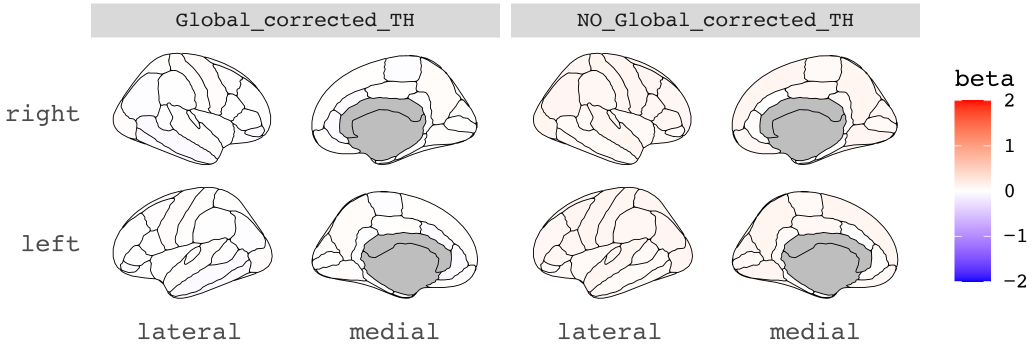

D. -log10 ($p_{fdr}$ values) of MR results of Bis.DB.ratio on regional TH.

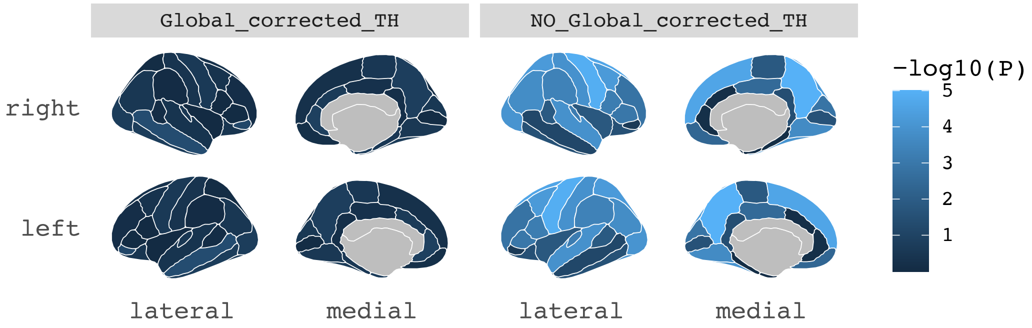

A. Beta coefficients of MR results of regional TH on Bis.FA.ratio.

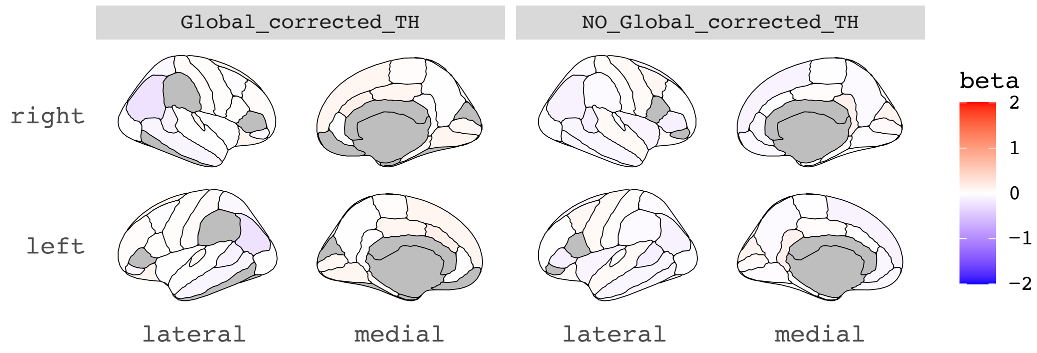

B. -log10 ($p_{fdr}$ values) of MR results of regional TH on Bis.FA.ratio.

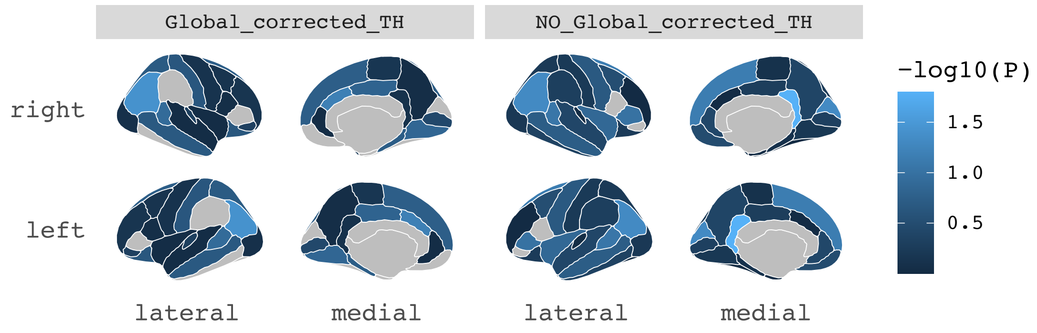

C. Beta coefficients of MR results of Bis.FA.ratio on regional TH.

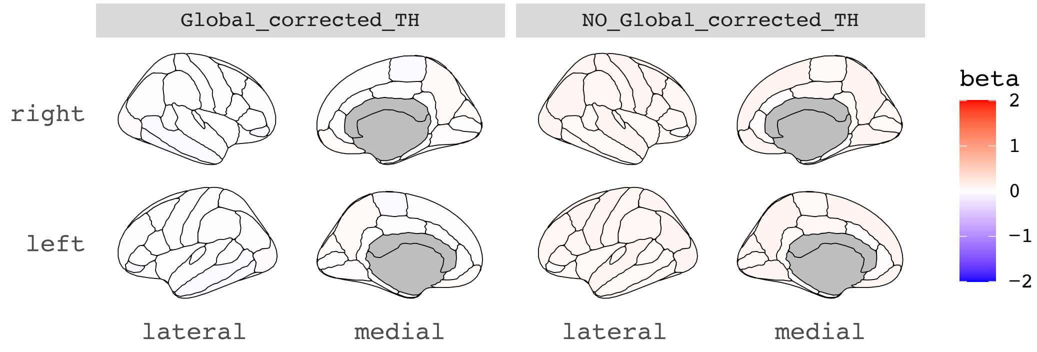

D. -log10 ($p_{fdr}$ values) of MR results of Bis.FA.ratio on regional TH.

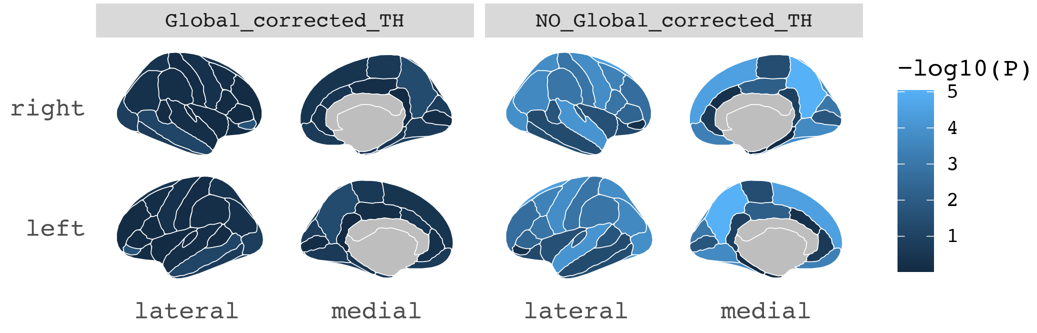

A. Beta coefficients of MR results of regional TH on DB.in.FA.

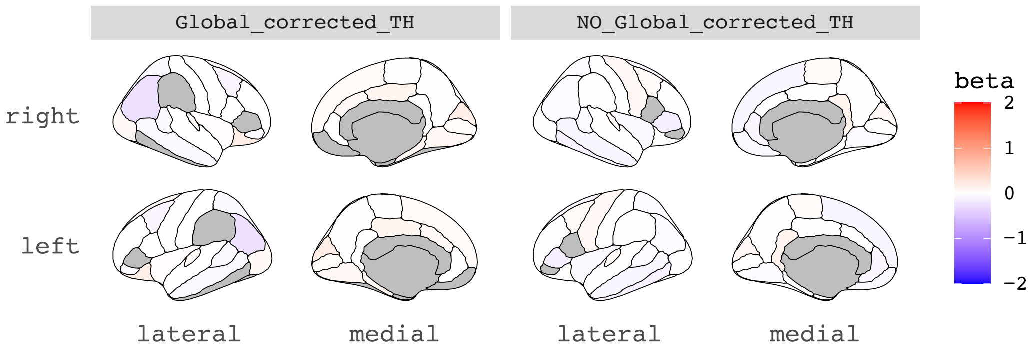

B. -log10 (p.$_{fdr}$ values) of MR results of regional TH on DB.in.FA.

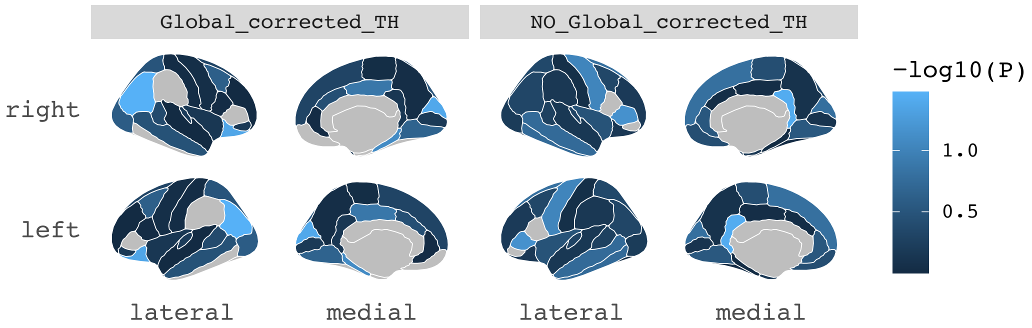

C. Beta coefficients of MR results of DB.in.FA on regional TH.

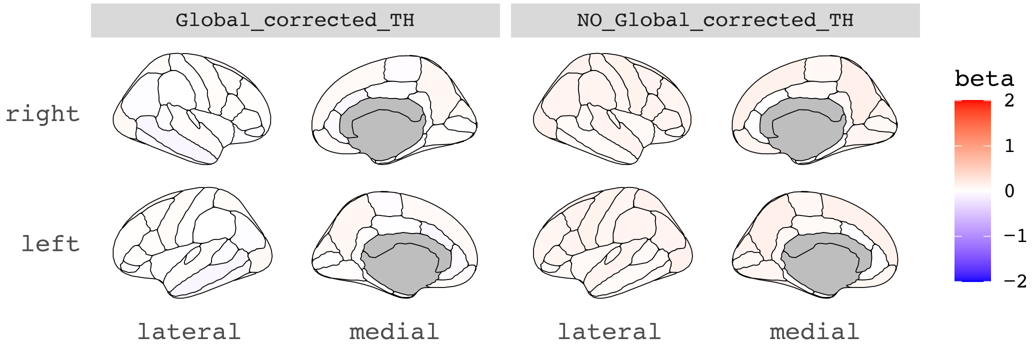

D. -log10 (p.$_{fdr}$ values) of MR results of DB.in.FA on regional TH.

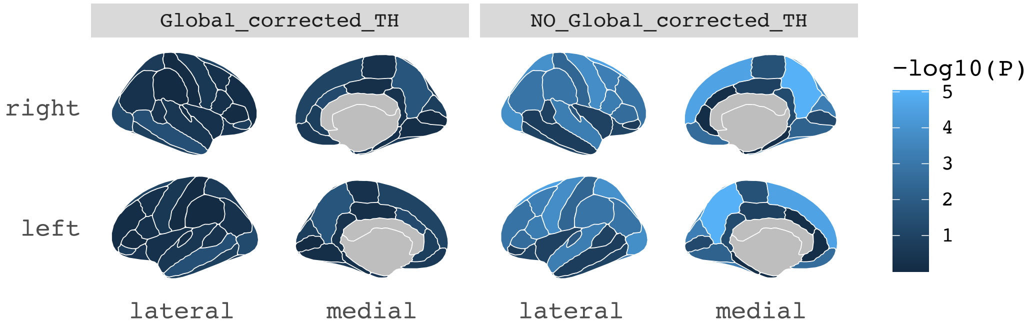

A. Beta coefficients of MR results of regional TH on CH2.DB.ratio

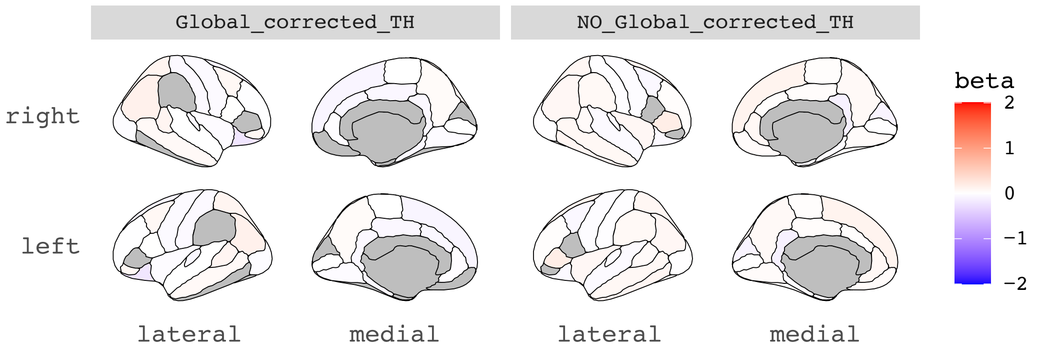

B. -log10 ($p_{fdr}$ values) of MR results of regional TH on CH2.DB.ratio

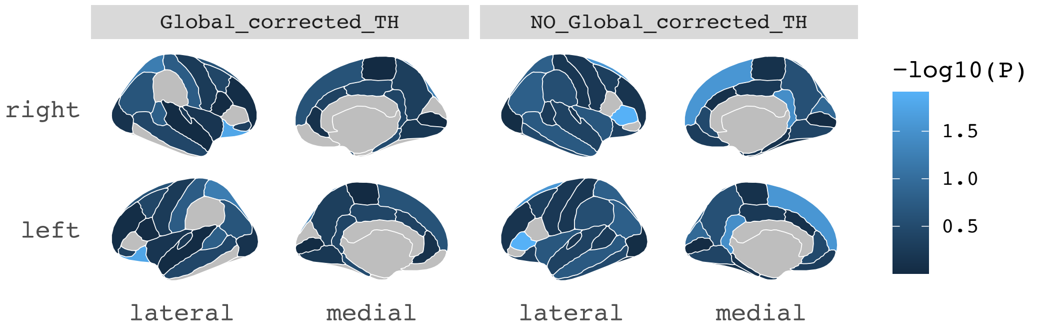

C. Beta coefficients of MR results of CH2.DB.ratio on regional TH.

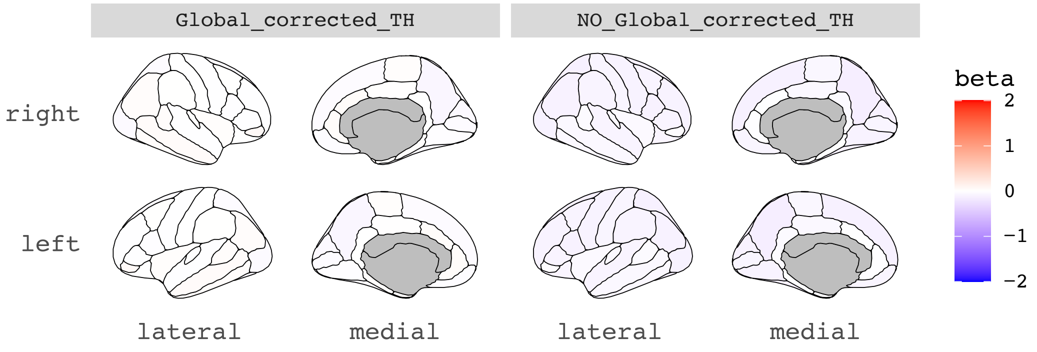

D. -log10 ($p_{fdr}$ values) of MR results of CH2.DB.ratio on regional TH.

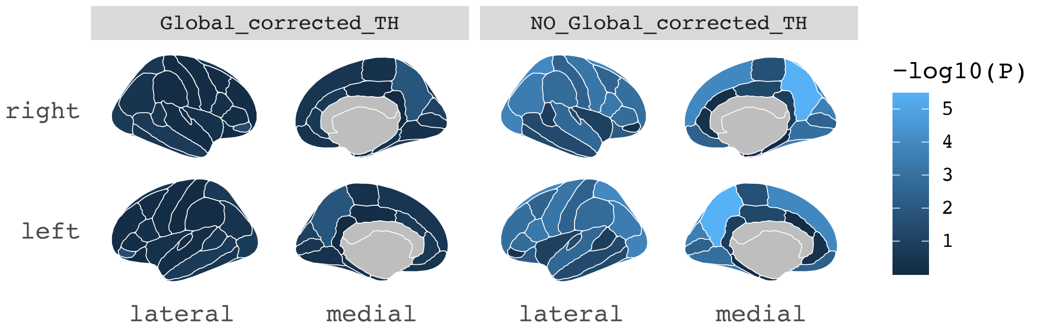

A. Beta coefficients of MR results of regional TH on CH2.in.FA.

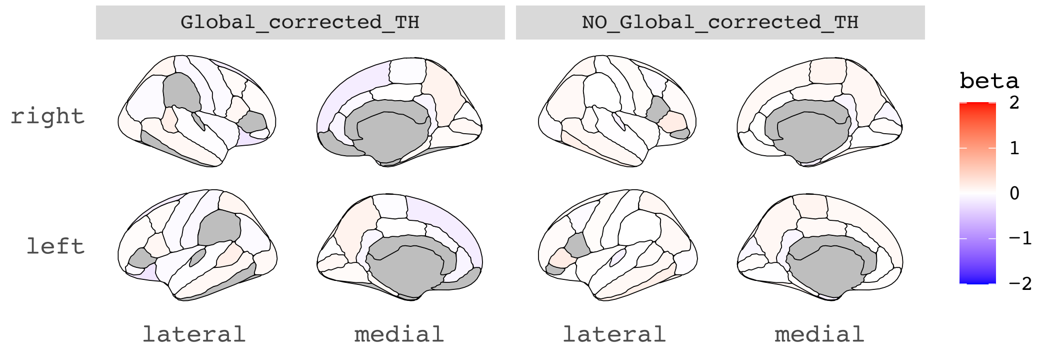

B. -log10 ($p_{.fdr}$ values) of MR results of regional TH on CH2.in.FA.

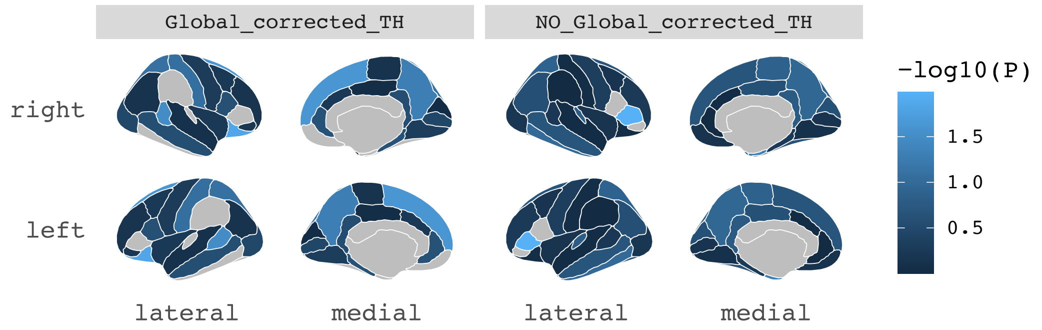

C. Beta coefficients of MR results of CH2.in.FA on regional TH.

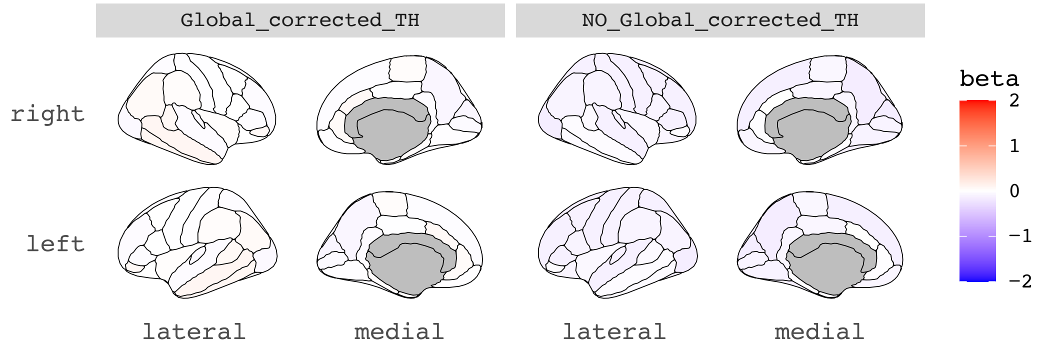

D. -log10 ($p_{.fdr}$ values) of MR results of CH2.in.FA on regional TH.

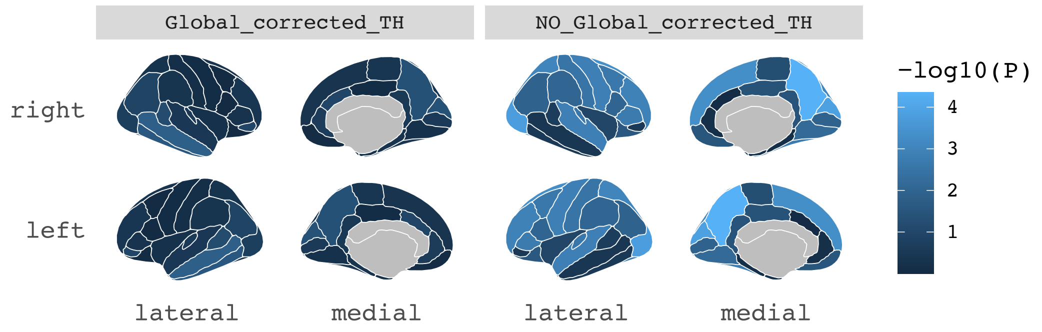

A. Beta coefficients of MR results of regional TH on otPUFA.

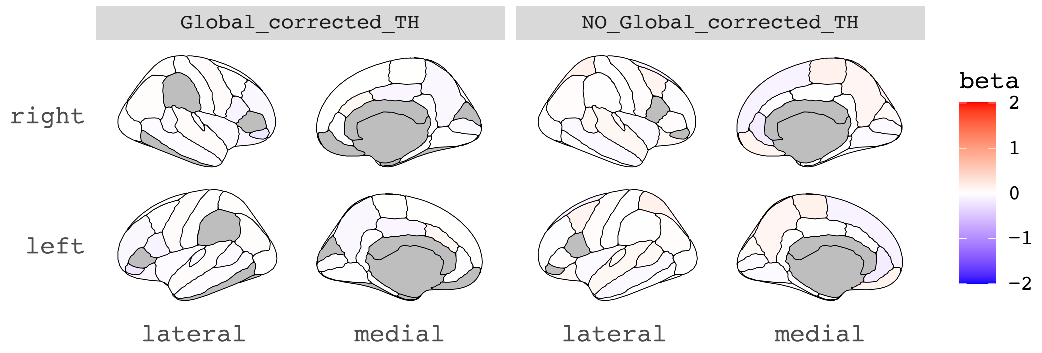

B. -log10 (p.$_{fdr}$ values) of MR results of regional TH on otPUFA.

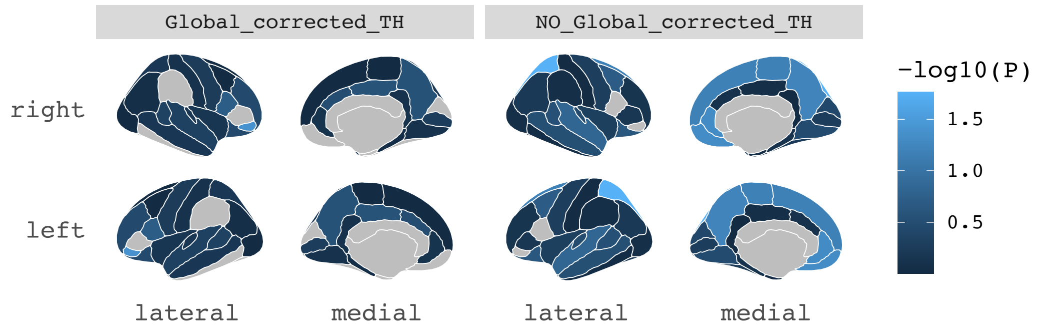

C. Beta coefficients of MR results of otPUFA on regional TH.

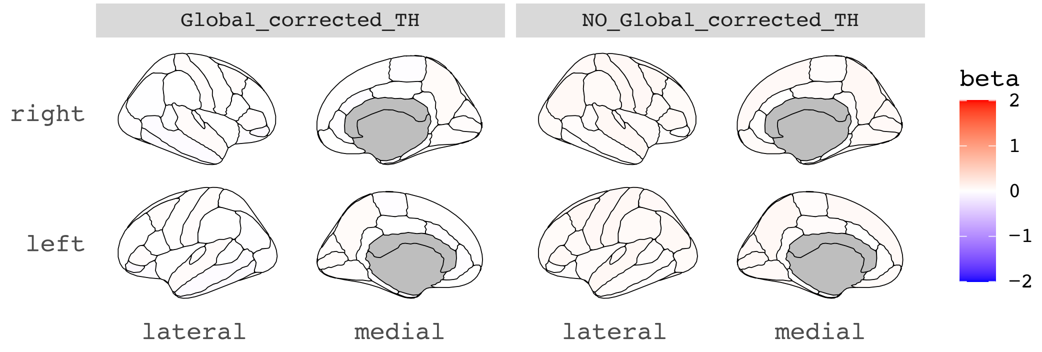

D. -log10 (p.$_{fdr}$ values) of MR results of otPUFA on regional TH.

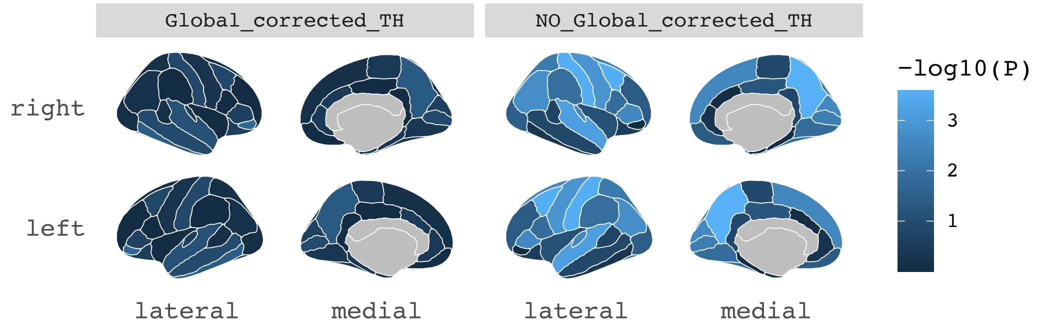

A. Beta coefficients of MR results of regional TH on creatinine (Crea).

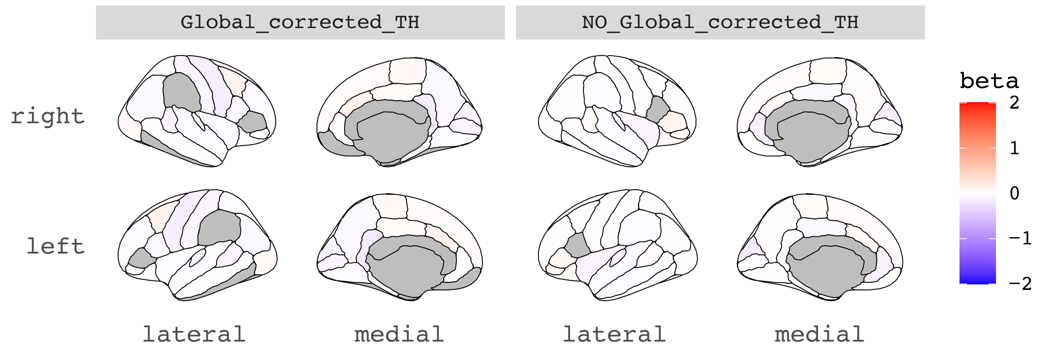

B. -log10 ($p_{.fdr}$ values) of MR results of regional TH on creatinine (Crea).

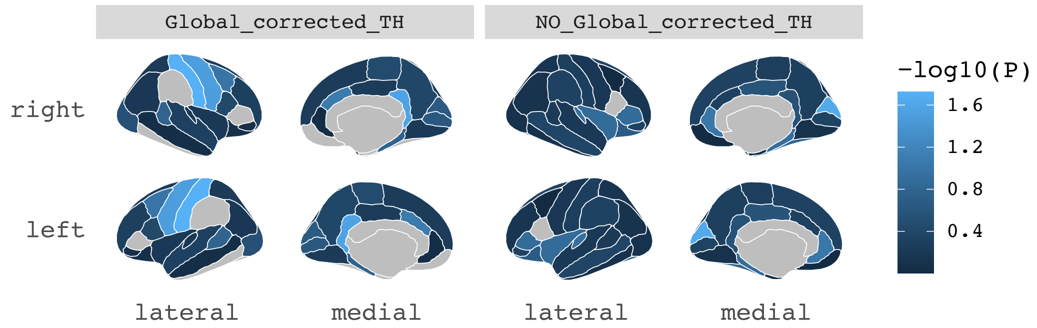

C. Beta coefficients of MR results of creatinine (Crea) on regional TH.

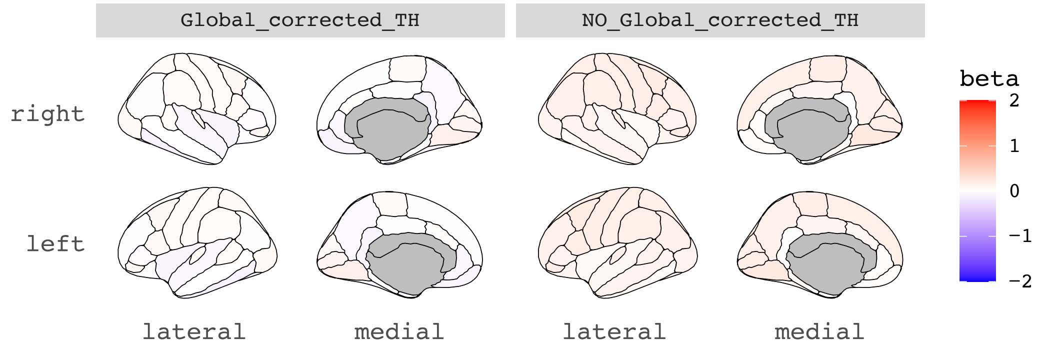

D. -log10 ($p_{.fdr}$ values) of MR results of creatinine (Crea) on regional TH.

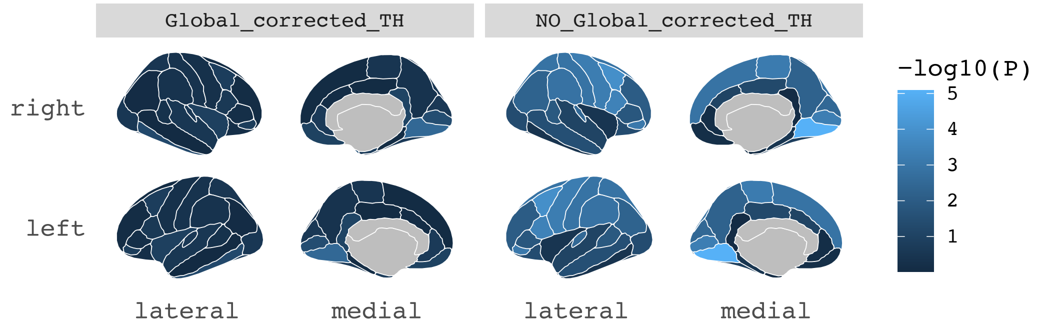

A. Beta coefficients of MR results of regional TH on bipolar disorder (BIP).

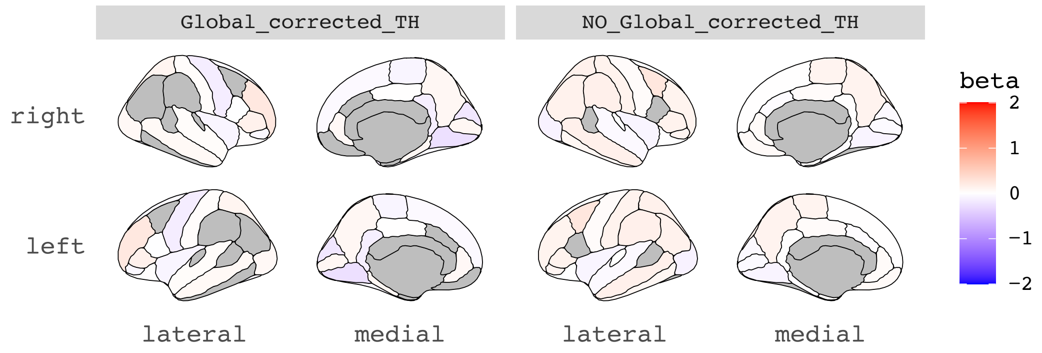

B. -log10 ($p_{fdr}$ values) of MR results of regional TH on bipolar disorder (BIP).

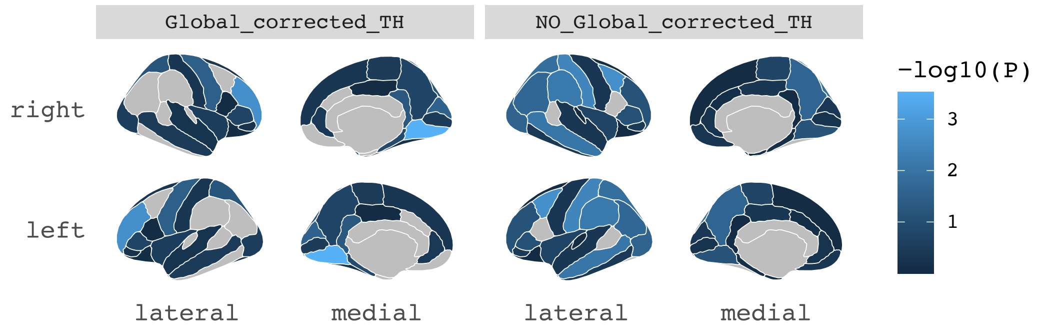

C. Beta coefficients of MR results of bipolar disorder (BIP) on regional TH.

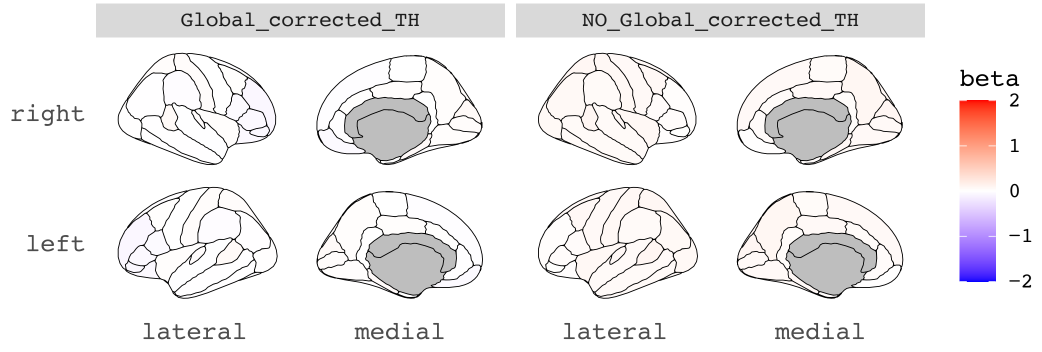

D. -log10 ($p_{fdr}$ values) of MR results of bipolar disorder (BIP) on regional TH.

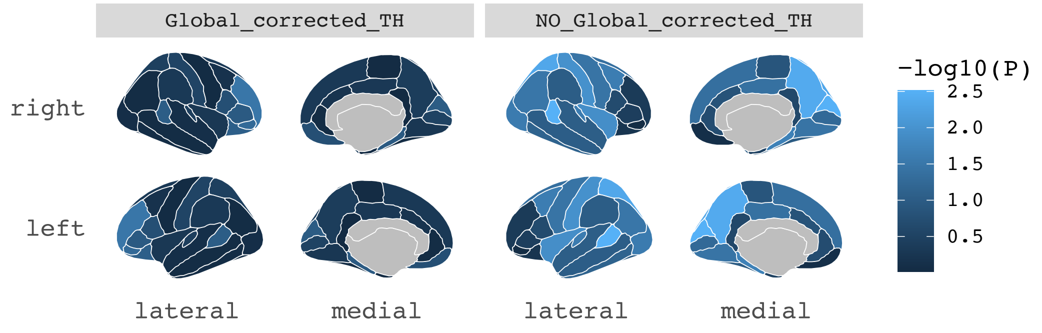

A. Beta coefficients of MR results of regional TH on body mass index (BMI).

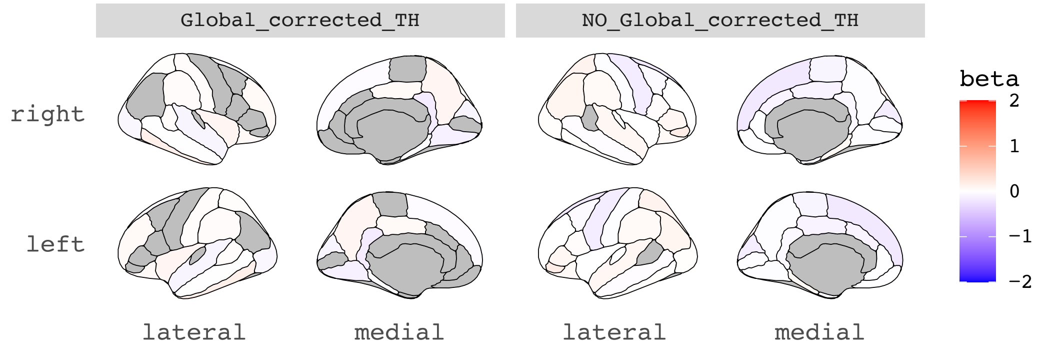

B. -log10 ($p_{fdr}$ values) of MR results of regional TH on body mass index (BMI).

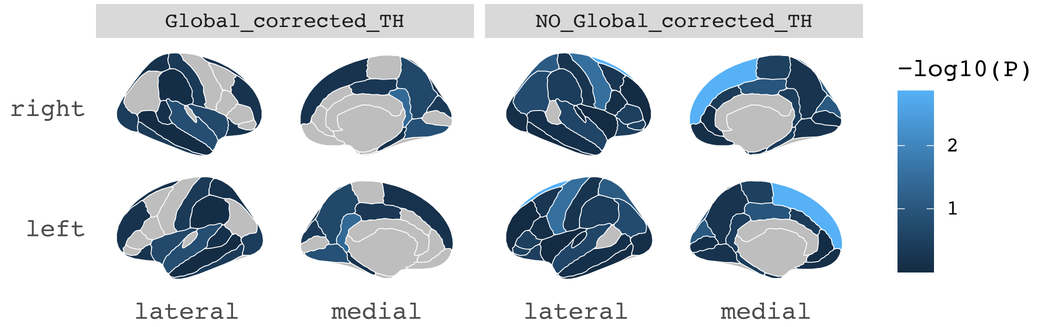

C. Beta coefficients of MR results of body mass index (BMI) on regional TH.

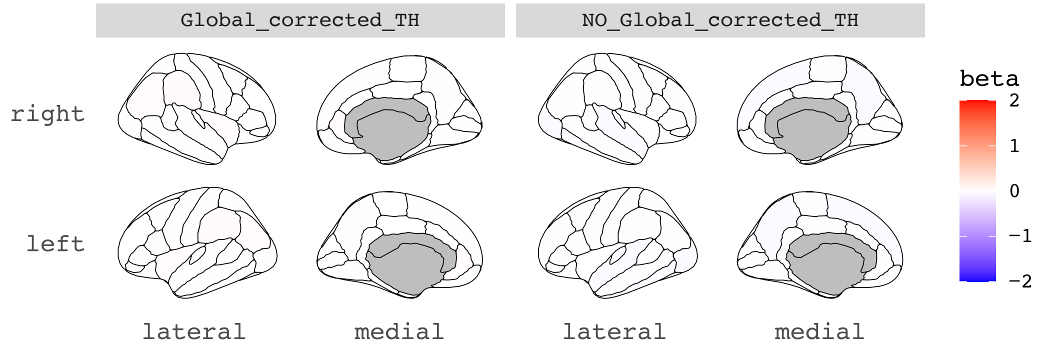

D. -log10 ($p_{fdr}$ values) of MR results of body mass index (BMI) on regional TH.

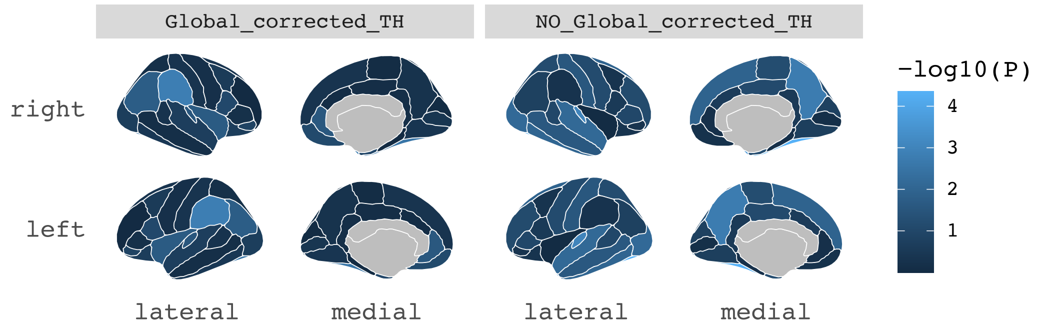
